# Evaluating Thrombectomy Devices and Combination Therapies in Acute ischemic Stroke: A systematic review & Network Meta-Analysis of 201 studies

**DOI:** 10.1101/2024.10.24.24316030

**Authors:** Annu Gulia, Manyata Srivastava, Manabesh Nath, Shubham Misra, Kamalesh Kumar Patel, Ashish Datt Upadhyay, Deepti Vibha, Shashvat Desai, Pradeep Kumar

## Abstract

**Background and Objective:** Mechanical Thrombectomy (MT) has become the standard treatment for acute ischemic stroke (AIS) caused by large vessel occlusion (LVO). However, the relative efficacy and safety of different MT devices remain uncertain. To evaluate and compare the safety and efficacy of different Thrombectomy devices and combination therapies in acute ischemic stroke, utilizing a network meta-analysis.

**Methods:** Patients receiving different MT devices (MERCI, TREVO, Solitaire, Penumbra, or a combination of MT devices) compared to Standard Care or Intravenous Thrombolysis (IVT), or Intra-arterial Thrombolysis (IAT) for AIS treatment. Safety outcomes were symptomatic intracranial hemorrhage (sICH) and all-cause mortality at 90 days. Efficacy outcome were good functional recovery at 90 days (defined as a modified Rankin Scale score of 0–2) and successful recanalization (measured by a TICI score of 2b-3).

**Results:** We included 201 studies, comprising 43 RCTs and 159 cohort studies with 71,154 AIS patients. TREVO device demonstrated the highest efficacy for functional recovery (OR=3.63, 95% CrI: 2.45–5.43), followed by MT + IVT (OR=2.87, CrI: 2.30–3.59). TREVO also achieved the highest rate of successful recanalization (OR 3.35, CrI: 1.36–8.19). MERCI, Solitaire and Aspiration devices were linked to a higher risk of sICH. For all-cause mortality at 90 days, TREVO notably reduced the odds (OR=0.56, CrI: 0.37–0.86), whereas aspiration devices showed no significant difference from standard treatments.

**Conclusion:** Our findings demonstrate contemporary stent-retriever device technology as the most effective option for improving functional recovery, recanalization success, and reducing mortality in AIS patients. These results highlight the critical need for selecting the most effective and safest thrombectomy device to optimize outcomes in acute stroke care.

## Introduction

Acute ischemic stroke (AIS) resulting from large vessel occlusion (LVO) represents a significant global health challenge, contributing to high morbidity and mortality rates.^1^ The introduction of mechanical thrombectomy (MT) has markedly transformed the management of AIS, providing substantial improvements in patient outcomes when performed promptly.^2^ Consequently, MT has become the standard treatment for LVO-induced AIS. The rapid advancement and proliferation of various MT devices have created a complex landscape of treatment options, each characterized by distinct designs, mechanisms of action, and clinical outcomes.^3^ Among the devices available, MERCI, TREVO, Solitaire, and Penumbra are commonly used in clinical practice, each with its unique operational approach and efficacy profile.^4,5^

The MERCI (Mechanical Embolus Removal in Cerebral Ischemia) device, one of the first approved for thrombectomy, utilizes a corkscrew-shaped wire to capture and retrieve the thrombus.^6^ Despite its pivotal role, MERCI has largely been surpassed by newer devices due to lower recanalization rates and higher complication rates.^7^ The Penumbra system, in contrast, employs direct aspiration through a catheter to remove the clot. This device has shown high effectiveness, particularly in cases where the clot is soft and amenable to aspiration, and has been associated with favourable outcomes in numerous studies.^8,9^ Newer-generation stent retrievers (SR) like TREVO and Solitaire have become prevalent in many stroke centres.^10–12^ These devices deploy a self-expanding stent to ensnare the clot within its mesh, facilitating retrieval as the stent is withdrawn.^13^ Clinical studies have demonstrated that TREVO and Solitaire offer superior recanalization rates and safety profiles compared to older devices such as MERCI, resulting in improved overall clinical outcomes.^14–16^

The swift evolution of MT technology has led to a diverse array of devices with varying operational mechanisms and clinical efficacy.^17^ Despite this progress, the optimal device choice for different clinical scenarios remains debated, as direct comparative studies are limited. To address this gap, we undertook a network meta-analysis to provide a comprehensive comparison of the efficacy and safety profiles of these thrombectomy devices.

## Methods

### Protocol Registration

This systematic review and network meta-analysis (NMA) was conducted in accordance with the Preferred Reporting Items for Systematic Reviews and Network Meta-Analyses (PRISMA-NMA) guidelines ^18^ and the study protocol was registered on PROSPERO.

### Inclusion Criteria

**Population (P):** Patients with Acute Ischemic Stroke (AIS).

**Intervention (I):** Mechanical Thrombectomy (MT) devices, including MERCI, TREVO, Solitaire, Penumbra, or a combination of MT devices.

**Comparator (C):** Standard Care, Intravenous Thrombolysis (IVT), or Intra-arterial Thrombolysis (IAT) with or without any MT devices.

### Outcomes (O)

Safety outcomes: symptomatic intracranial hemorrhage (sICH) and all-cause mortality at 90 days.

Efficacy outcome: good functional recovery at 90 days (defined as a modified Rankin Scale score of 0–2) and successful recanalization (measured by a TICI score of 2b-3).

**Study Design (S):** Randomized controlled trials (RCTs) or cohort studies (prospective or retrospective)

### Exclusion Criteria

- Had non-comparative designs.
- Were conference proceedings, dissertations, preprints, ongoing studies, or grey literature without relevant outcome data.

### Literature Search

A comprehensive search strategy was employed, combining keywords and medical subject headings (MeSH) related to acute ischemic stroke, mechanical thrombectomy, and specific MT devices. Detailed search strategies are outlined in supplementary materials (**Table eS1**) covering publications up to June 30, 2024. To ensure completeness, references from relevant articles, including meta-analyses, reviews, and RCTs, were also reviewed.

### Study Selection and Data Extraction

Reviewers working in pairs, independently screened abstracts and full-text articles, and extracted data using a pre-specified form according to the inclusion and exclusion criteria. Data extraction included:

**Basic Information:** Study name, countries, number of centers, treatment comparisons, and number of participants.

**Participant Characteristics:** Age, sex ratio, baseline National Institutes of Health Stroke Scale (NIHSS) scores, baseline Alberta Stroke Program Early Computed Tomography (ASPECT) score, details of anterior/posterior circulation occlusion, treatment procedures, and outcomes.

**Outcomes:** All outcomes were reported as dichotomous variables.

Discrepancies during study selection and data extraction were resolved through discussion with the corresponding author to reach a consensus

### Quality Assessment

The quality of the included studies was assessed to ensure the reliability and validity of the findings. For RCTs, the modified Jadad scale^19,20^ was utilized. For cohort studies, the Newcastle-Ottawa Scale (NOS)^21^ was employed.

### Statistical Analysis

We conducted a pairwise meta-analysis of direct comparisons using a Bayesian random-effects model to obtain direct evidence. Treatment effects were estimated using odds ratios (OR) and 95% credible intervals (CrI) for dichotomous outcomes. Credible intervals provide a range of plausible values for the OR based on the observed data. We also calculated the Surface Under the Cumulative Ranking (SUCRA) curve values, including their CrI, to assess the uncertainty in MT device rankings. A SUCRA value closer to 100% indicates a higher rank in effectiveness (or safety) compared to other MT devices assessed in the NMA. Online web based MetaInsight tool (MetaInsight V4.0.2 Beta software) for Bayesian network meta-analyses were employed using random-effects model for all comparisons.^22^

## Results

### Selection Process

**Figure-1** illustrates the PRISMA flow diagram outlining the selection process and the reasons for exclusion from the NMA. Of the 8,581 records initially identified, 6,080 were excluded prior to screening, leaving 2,501 records for review. After excluding 1,889 records, 612 reports were sought for retrieval; however, 247 could not be obtained. Among the 365 reports assessed for eligibility, 164 were excluded, resulting in 201 studies included in the final analysis comprising 43 RCTs and 158 cohort studies.

**Figure 1:**
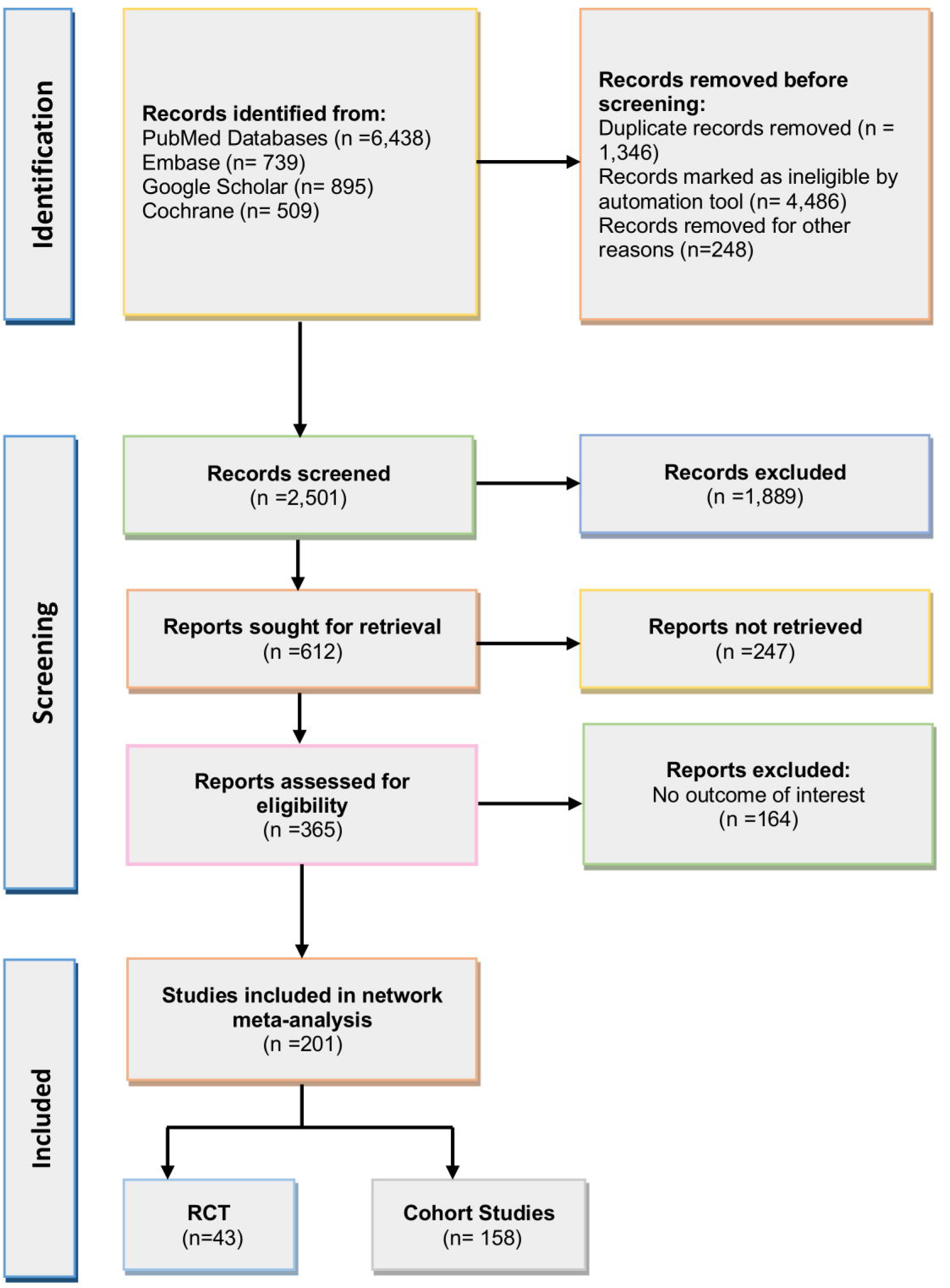
PRISMA Flow Diagram for Study search, selection, and inclusion process.

### Study Characteristics

**eTable-S2** presents the characteristics and outcomes of the included studies. The studies were published between 2008 and 2024. Of the 201 studies, 113 were multicentric, 71 were single-centric, 3 were dual-centric, and 11 did not report their centricity. The sample sizes varied from 9 to 10,548 across different study arms. The criteria for defining sICH included the European Cooperative Acute Stroke Study III (ECASS III), the National Institute of Neurological Disorders and Stroke (NINDS), an increase of 4 points in the National Institutes of Health Stroke Scale (NIHSS) score, and the Safe Implementation of Thrombolysis in Stroke-Monitoring Study (SITS-MOST). Successful recanalization was defined by the modified Treatment in Cerebral Ischemia (mTICI), the Thrombolysis in Cerebral Infarction (TICI), and the expanded Treatment in Cerebral Ischemia (eTICI) scores (2b-3). Studies were conducted across seven distinct countries, including 41 from China, 37 from the United States of America, 21 from Germany, 15 from France, and 7 each from Japan, South Korea, and Netherlands. All the included studies enrolled AIS patients ranging from 3 to 24 hours after the symptom onset.

The included studies that did not specify the exact device used were grouped under a general category of mechanical thrombectomy (MT) devices, with subcategories including MT combined with intra-arterial therapy (MT + IAT), MT combined with intravenous thrombolysis (MT + IVT). Standard treatment (ST), IAT and IVT were separate comparator in limited studies (**Figure-eS1**).

This approach allowed for the inclusion of studies where device specifics were not available, ensuring broader categorization. For studies that did not provide the exact name of the thrombectomy devices but described the general approach, we categorized them based on their mechanism of action. If a study utilized stent retrievers, aspiration devices, or a combination of both, they were grouped accordingly into stent retrievers (SR), aspiration devices (ASP), or a combination of both (SR + ASP). Finally, for studies that explicitly named the thrombectomy devices used, they were categorized based on the device, such as Penumbra, MERCI, Solitaire, or TREVO.

The analysis incorporated 12 interventions across all outcomes, with 13 to 15 multi-arm studies and 126 to 159 two-arm studies per outcome. There were 42 to 46 pairwise comparisons with direct data out of 66 possible comparisons. The total number of studies ranged from 137 for sICH to 172 for mRS, covering 52,114 to 71,154 patients and resulting in 4,221 to 52,086 events within the network. **Table-1** indicates that no study reported zero events for all safety as well as efficacy outcomes. **Figure 2 (a-d)** shows the network geometry plots with the available direct comparisons for all efficacy (good functional recovery and successful recanalization) and safety outcomes for sICH within 24 hours and all-cause mortality at 90 days.

**Figure 2.**
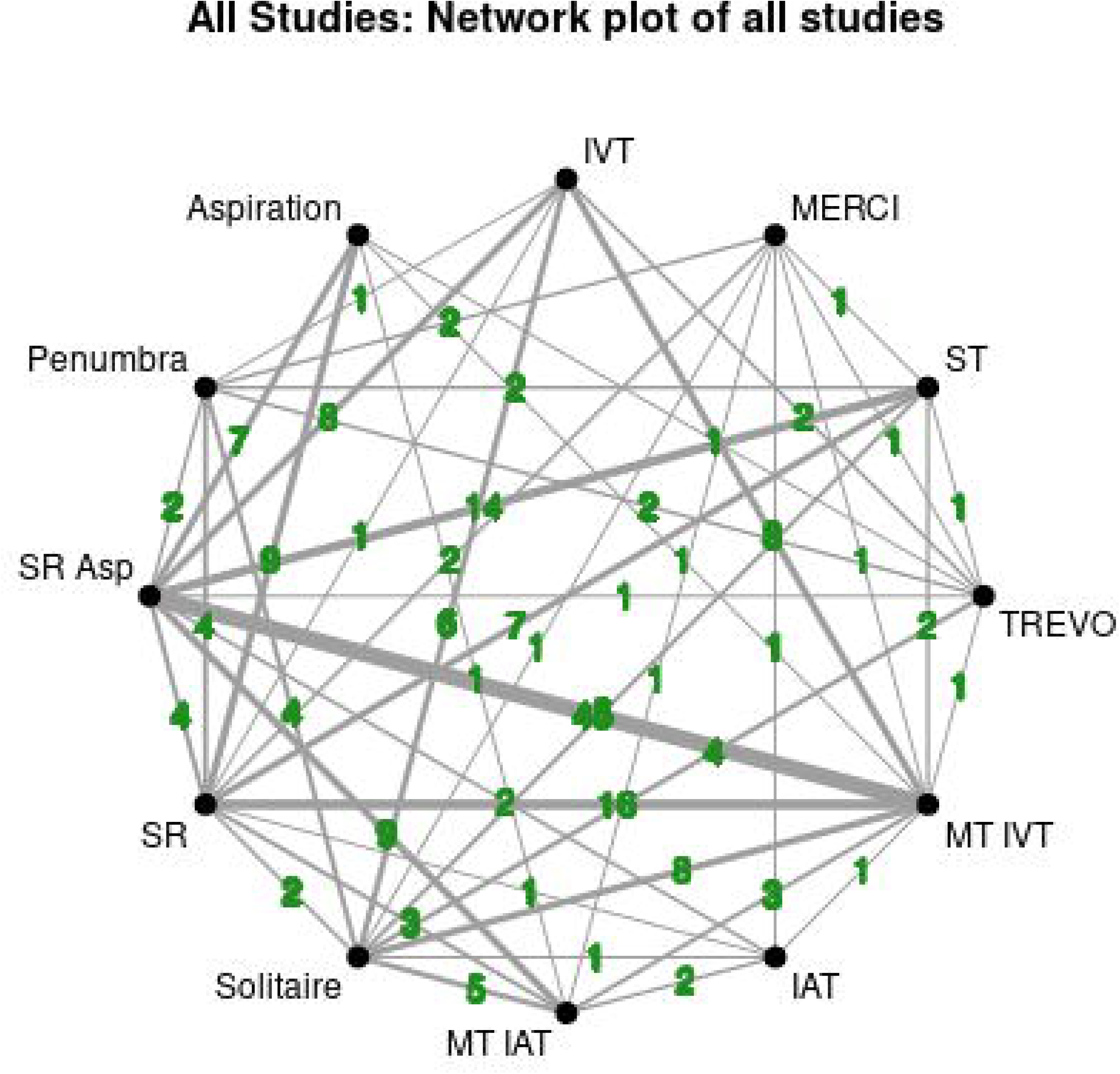
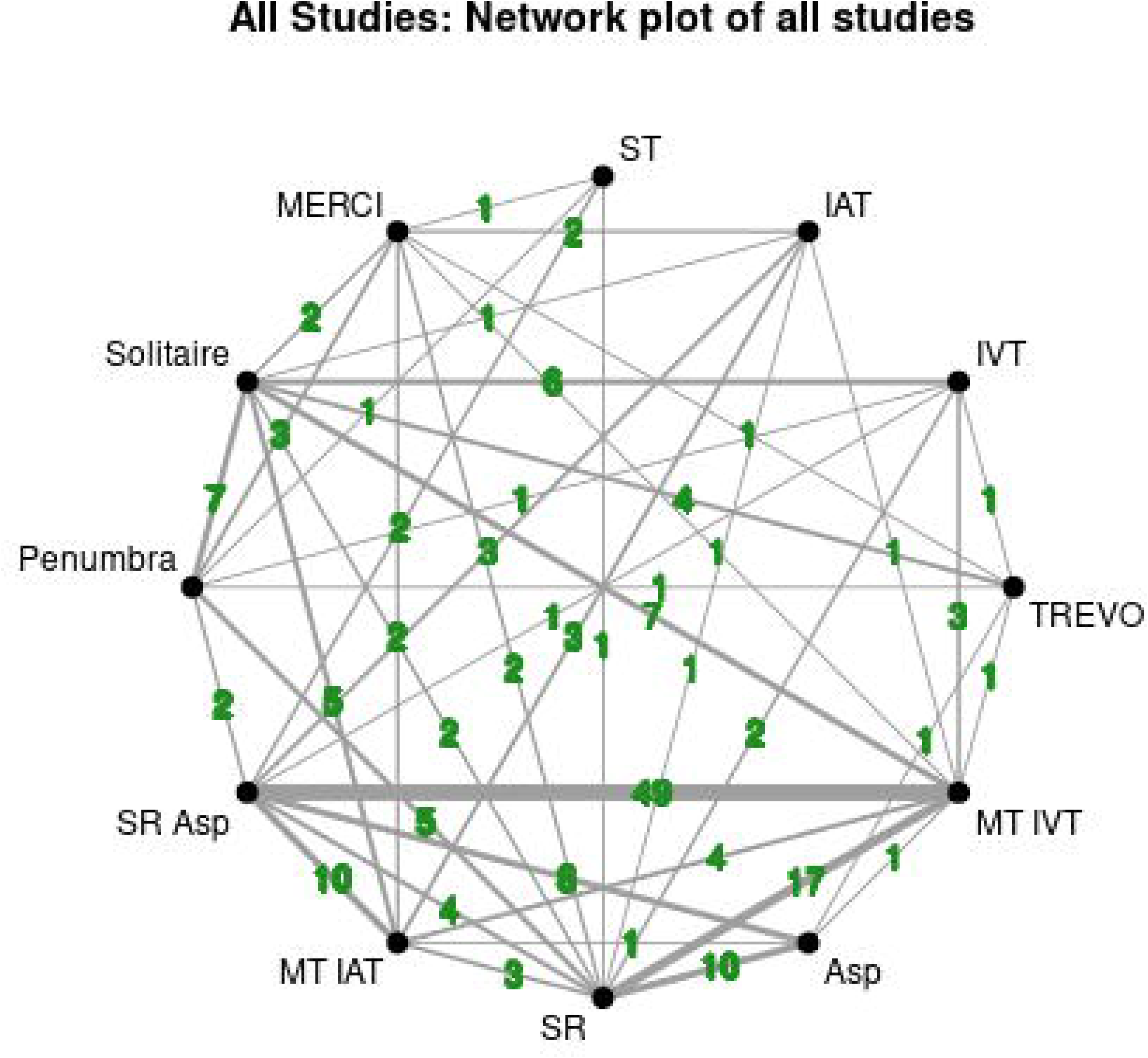
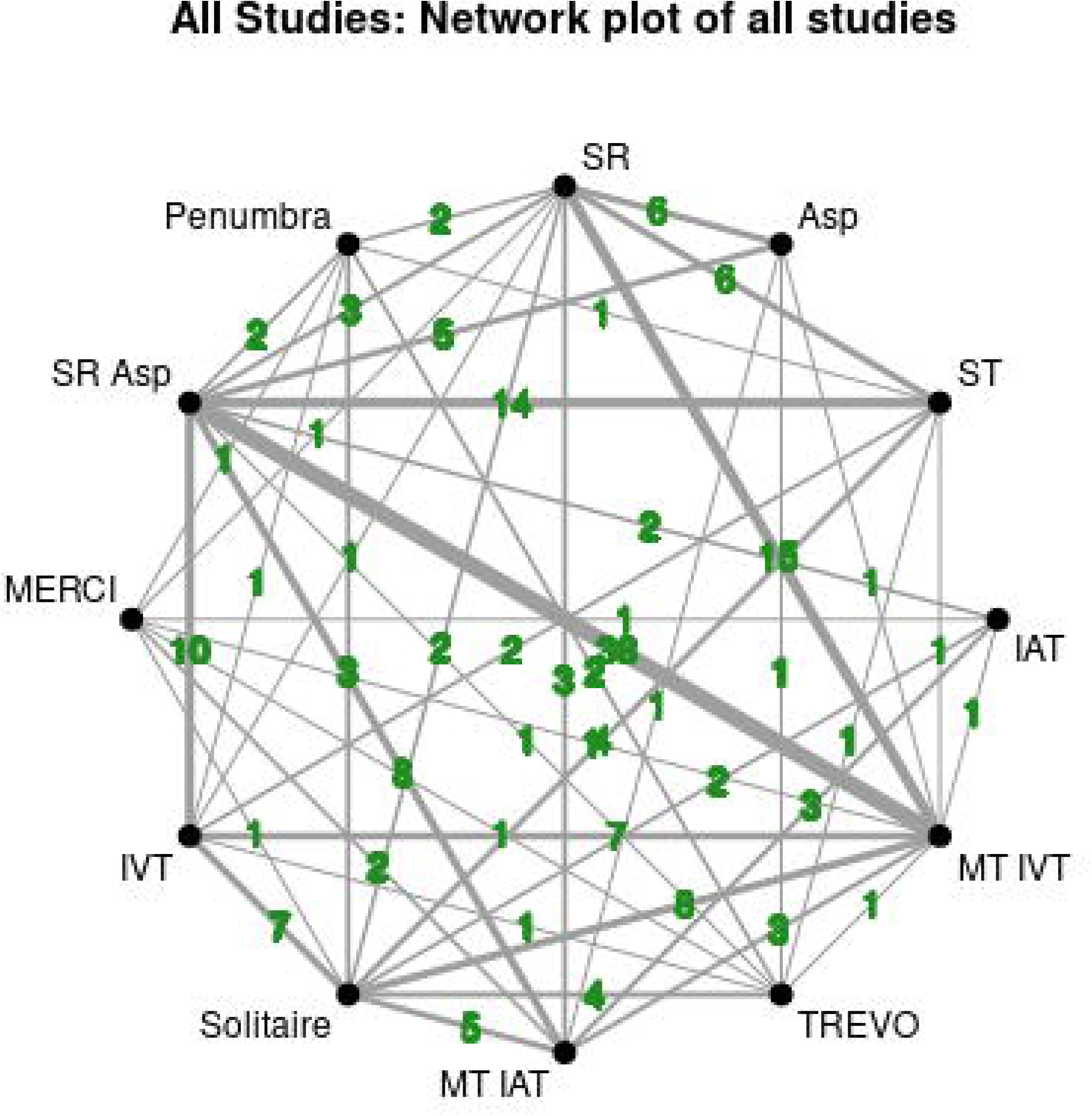
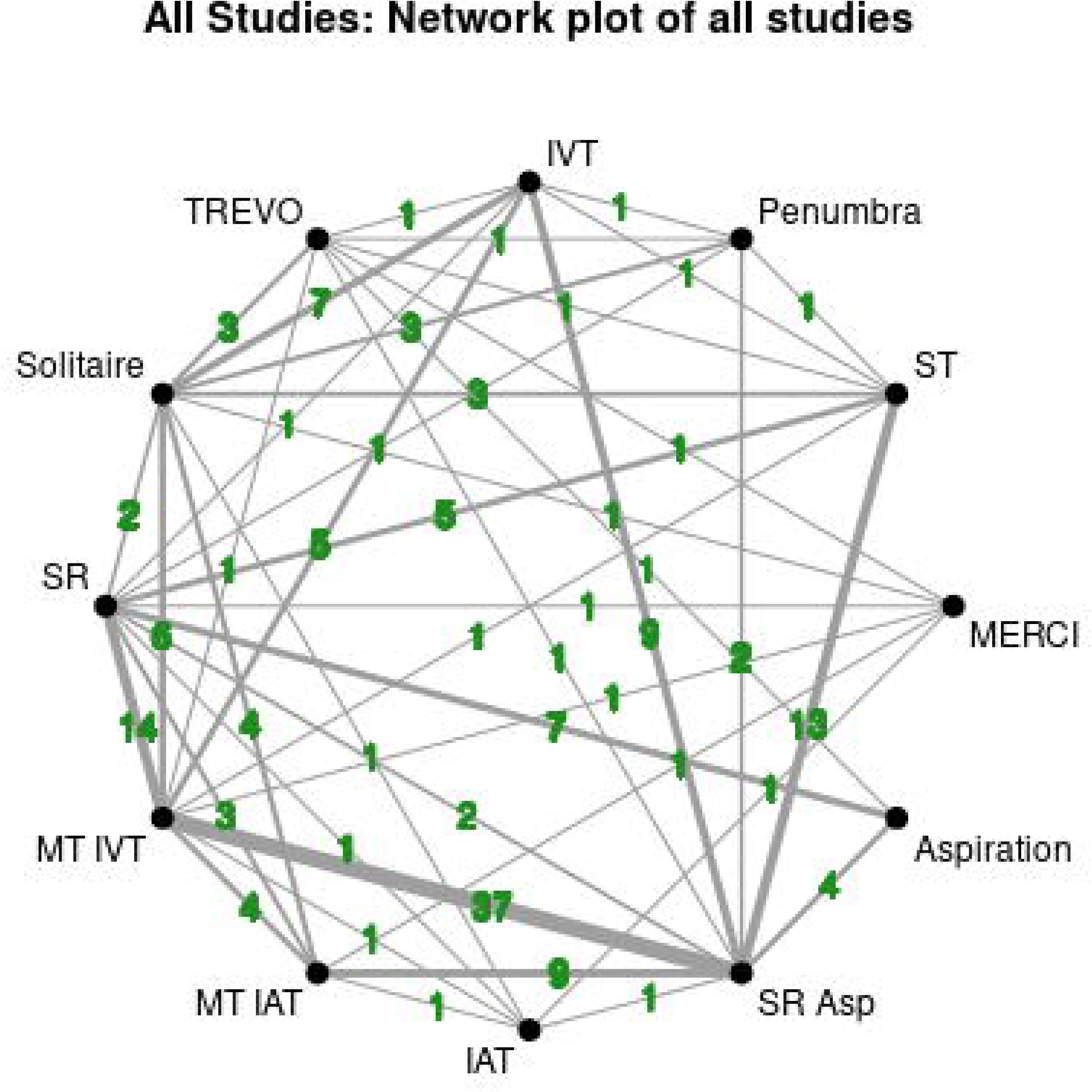
**(a-d):** Network map for the complete evidence network for: (a) good functional recovery at 90 days; (b) successful recanalization; (c) all-cause mortality at 90 days and (d) sICH within 24 hours.

**Table.1:** Characteristics table of all studies.

| Characteristic | Outcome |  |  |  |
| --- | --- | --- | --- | --- |
|  | mRS | Recanalization | Mortality | sICH |
| Number of Studies With At Least One Zero Event | 0 | 0 | 0 | 0 |
| Number of Studies with All Zero Events | 0 | 0 | 0 | 0 |
| Number of Multi-Arms Studies | 13 | 15 | 13 | 11 |
| Number of Interventions | 12 | 12 | 12 | 12 |
| Total Number of Pairwise Comparisons With Direct Data | 46 | 42 | 45 | 43 |
| Total Possible Pairwise Comparisons | 66 | 66 | 66 | 66 |
| Number of Two-arm Studies | 159 | 135 | 141 | 126 |
| Number of Studies | **172** | **150** | **154** | **137** |
| Number of Studies With No Zero Events | 172 | 150 | 154 | 137 |
| Total Number of Events in Network | **25720** | **52086** | **11454** | **4221** |
| Total Number of Patients in Network | **71154** | **66245** | **52114** | **65839** |

### Quality Assessment

All 43 RCTs and 159 cohort studies included in the analysis were of high methodological quality with a low risk of bias (**Table-eS2 and eS4[a-b]**). This determination was made using rigorous assessment tools: RCTs were evaluated using modified Jadad Score, while cohort studies were assessed with the NOS Scale. These evaluations confirm that the study designs and reporting standards were methodologically sound, ensuring the reliability, validity of the findings and generalizability of the conclusions derived from the aggregated data.

## Main Findings

### Efficacy outcomes

#### a. Good functional recovery (mRS ≤2 at 90 days)

TREVO device showed the highest odds ratio (OR=3.63, 95% CrI: 2.45–5.43) for achieving good functional recovery at 90 days as compared to ST, followed closely by intra-arterial thrombolysis (IAT) (OR=2.84, 95% CrI: 1.70–4.74) and the combination of MT + IVT (OR =2.87, 95% CrI: 2.30–3.59). MERCI device demonstrated the lowest improvement (OR 1.63, 95% CrI: 1.04–2.55) **[Figure 3(a)]**.

**Figure 3.**
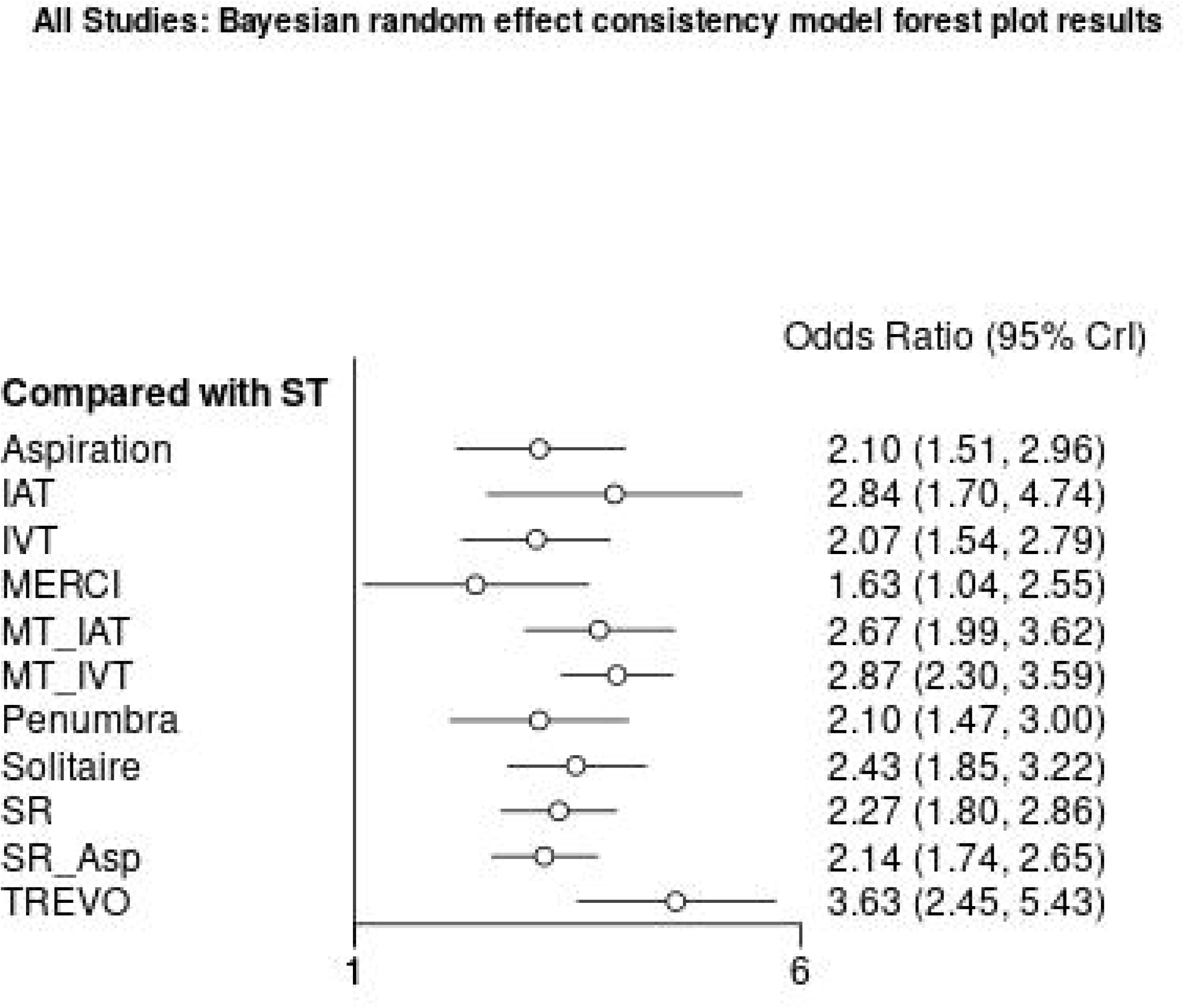
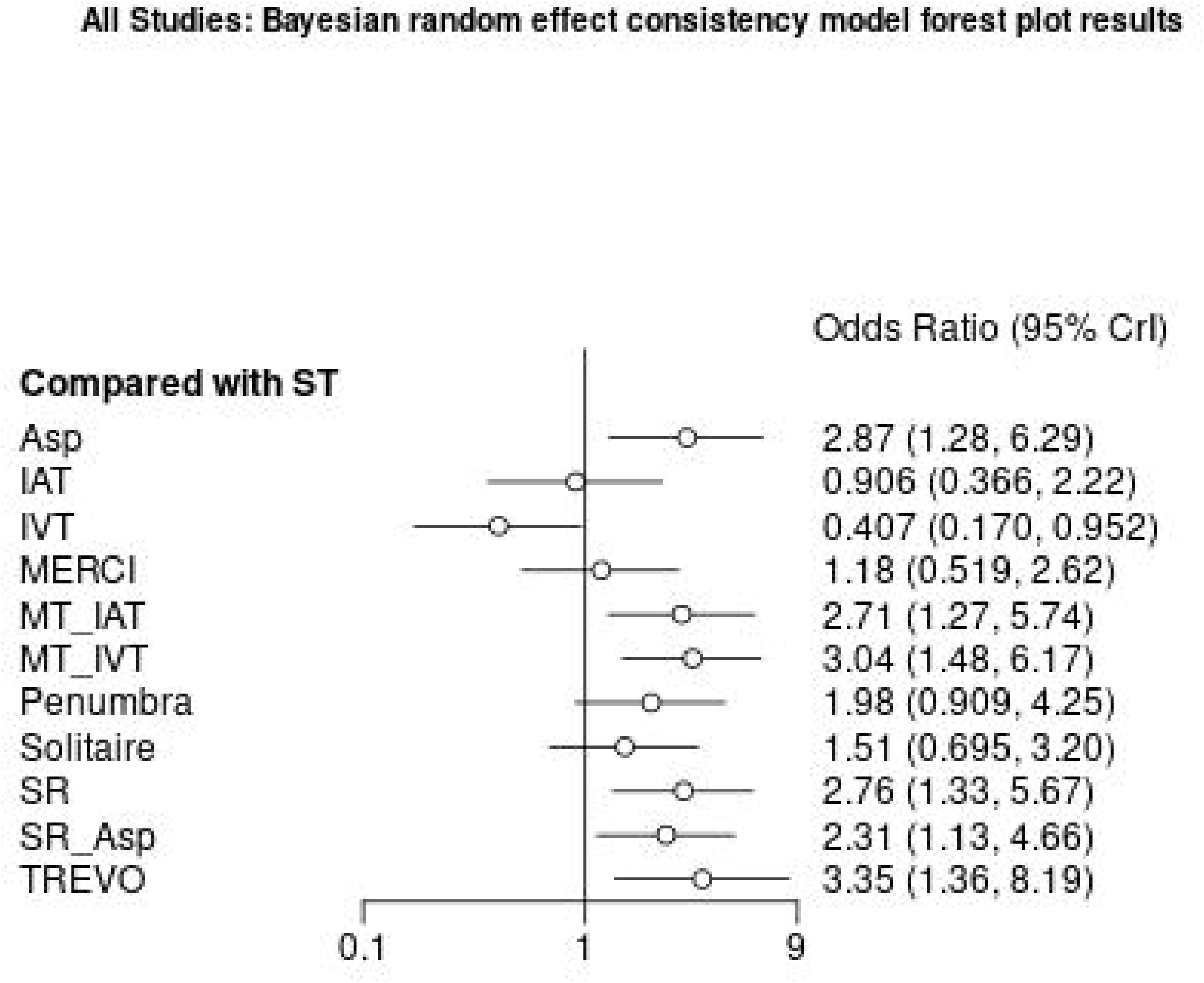
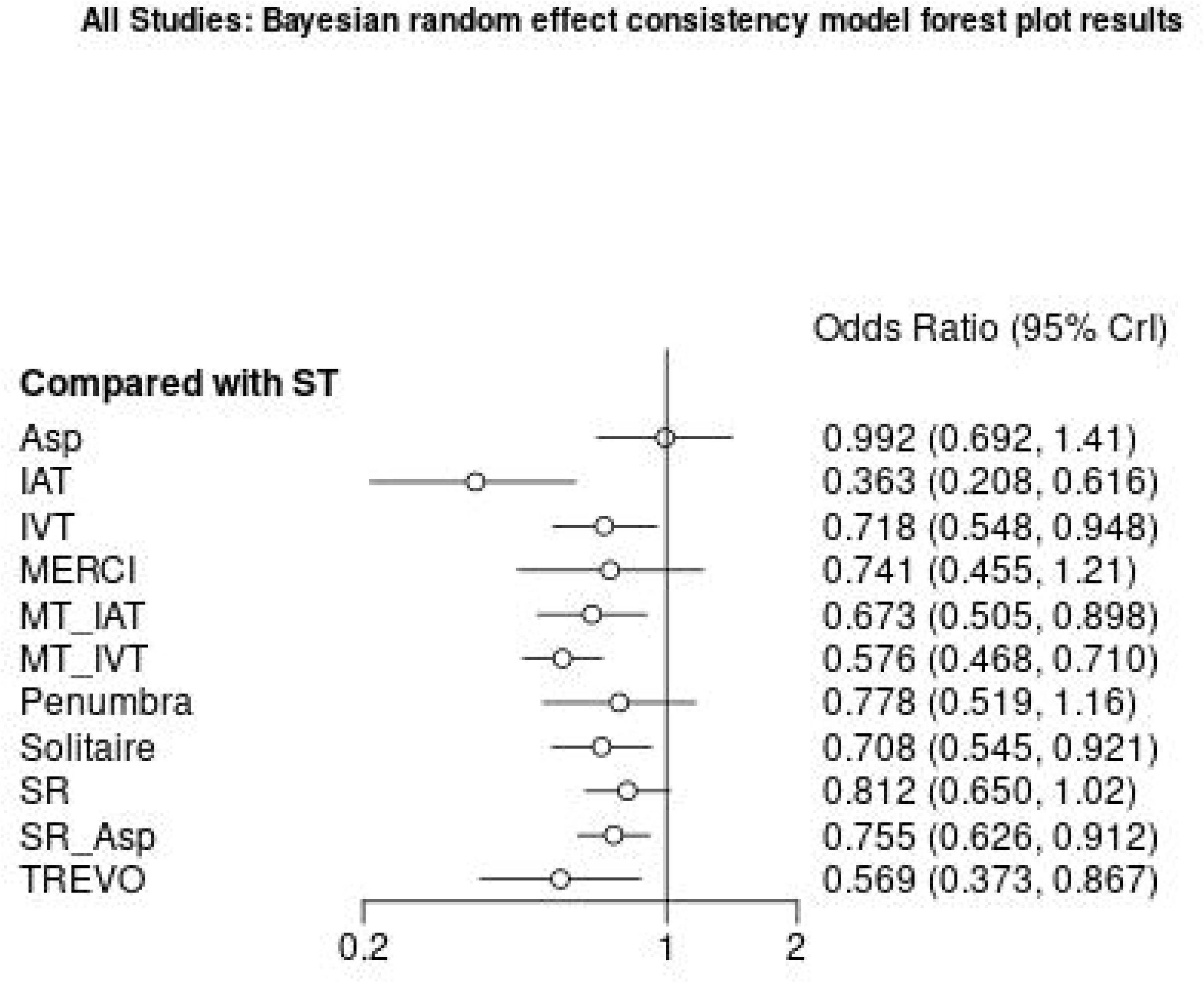
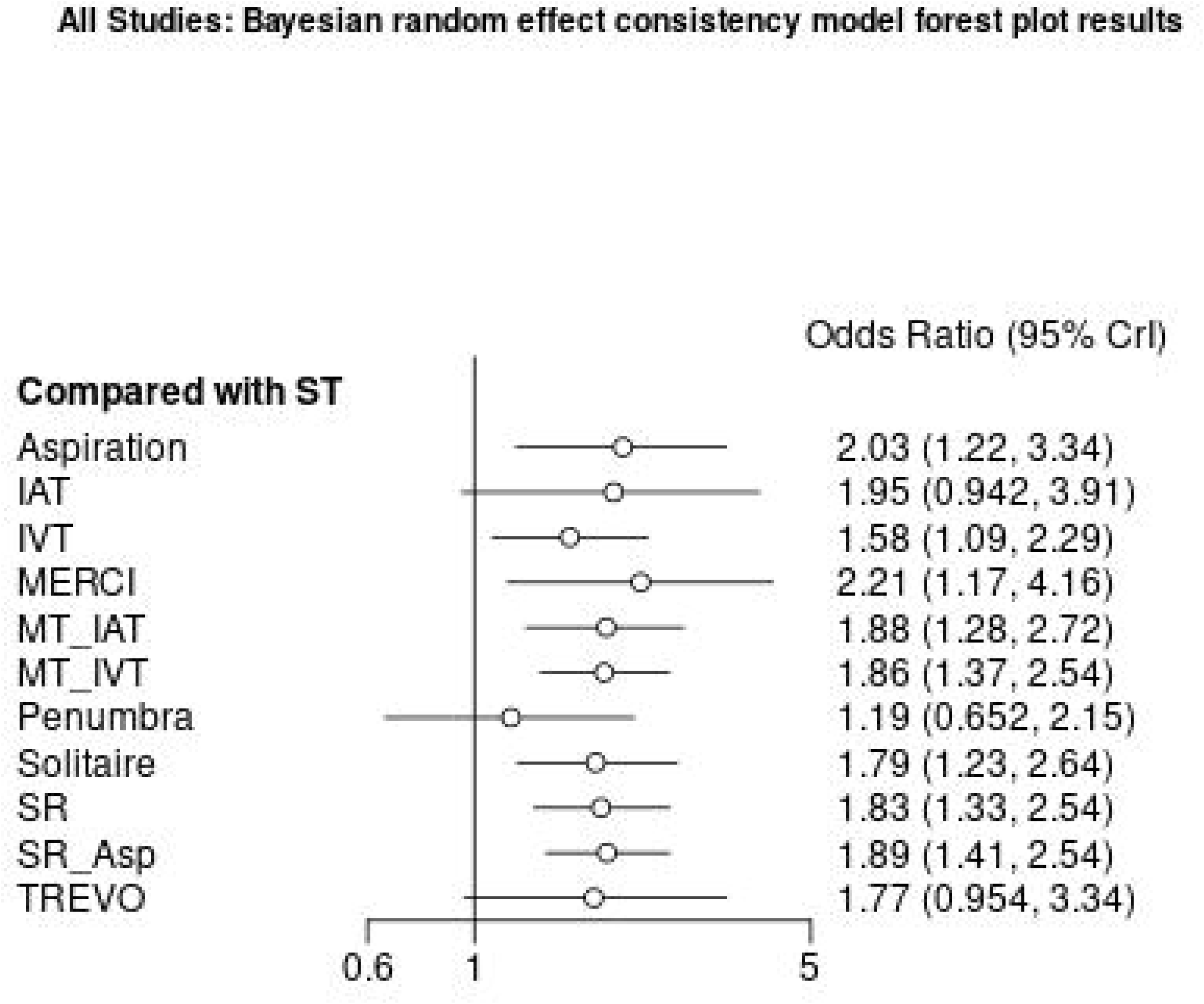
**(a-d)**: Forest plot of relative effects from Bayesian random effect consistency model for: (a) good functional recovery at 90 days; (b) successful recanalization; (c) all-cause mortality at 90 days and (d) sICH within 24 hours.

#### b. Successful recanalization (TICI or TIMI flow ≥2b)

Successful recanalization was observed for TREVO device (OR 3.35, 95% CrI: 1.36–8.19), followed by (MT + IVT) (OR 3.04, 95% CrI: 1.48–6.17) and Asp (OR 2.87, 95% CI: 1.28–6.29) as compared to ST. Conversely, IVT significantly reduced the odds of successful recanalization (OR 0.41, 95% CrI: 0.17–0.95), and IAT also demonstrated lower odds (OR 0.91, 95% CrI: 0.37–2.22) but non-significant [**Figure-3b**].

### Safety outcomes

#### Symptomatic Intracranial Hemorrhage (sICH) within 24 hours

The MERCI device demonstrates the highest risk of sICH (OR=2.21; 95% CrI: 1.17–4.16), indicating that it more than doubles the odds of sICH compared to ST. Aspiration is similarly associated with a significant increase in risk (OR=2.03, 95% CrI: 1.22–3.34), as are the Solitaire device (OR=1.79, 95% CrI: 1.23–2.64), Stent Retriever (OR=1.83, 95% CrI: 1.33–2.54), and the combination of SR+Asp (OR= 1.89, 95% CrI: 1.41–2.54). Additionally, MT+IAT (OR 1.88, 95% CrI: 1.28–2.72) and MT+IVT (OR 1.86, 95% CrI: 1.37–2.54) significantly increased the odds of sICH.

In contrast, the Penumbra device exhibits a non-significant increase in risk (OR 1.19, 95% CrI: 0.65–2.15). The TREVO device (OR 1.77, 95% CrI: 0.954–3.34) and IAT (OR 1.95, 95% CrI: 0.94–3.91) suggest a potential increase in the odds of sICH, but their credible intervals include 1, rendering these findings not statistically significant. Overall, while several endovascular interventions are associated with an increased risk of sICH, the MERCI device presents the highest risk among those analyzed **[Figure-3c]**.

### All-cause Mortality at 90 days

TREVO device demonstrated the greatest reduction in mortality (OR=0.57; 95% CrI: 0.37–0.87), suggesting a substantial protective effect. This was followed closely by MT+IVT, which also showed a marked reduction in mortality (OR=0.58, 95% CrI: 0.47–0.71), and IAT, exhibited the most pronounced effect (OR=0.36, 95% CrI: 0.21–0.62). In contrast, aspiration (OR=0.99, 95% CrI: 0.69–1.4) did not result in a statistically significant reduction in mortality compared to ST, indicating no meaningful difference. Overall, the TREVO device and MT+IVT were the most effective interventions for reducing 90-day mortality, whereas aspiration had minimal impact **[Figure-3d]**.

The league table, presented as **Table eS3 (a-d)**, compares all treatment pairs and provides estimates from the NMA. According to the radial SUCRA plot **[Figure 4(a-d)]** and Litmus Rank-O-Gram **[Figure 5 (a-d)]**, the TREVO device and MT + IVT exhibited the highest SUCRA values, indicating they were the most effective interventions. Compared to ST, TREVO and MT + IVT were associated with significantly higher rates of functional recovery at 90 days. Based on SUCRA values, the TREVO device (92% probability) and MT + IVT (85% probability) were identified as the most effective procedures for AIS. These interventions also significantly increased the odds of favourable outcomes compared to intravenous thrombolysis (IV t-PA). The large node sizes for TREVO and SR+Asp reflect a substantial number of participants, while the thicker connections between nodes, particularly among TREVO, MT + IVT, and SR, indicate these treatments were assessed in a higher number of trials.

**Figure 4.**
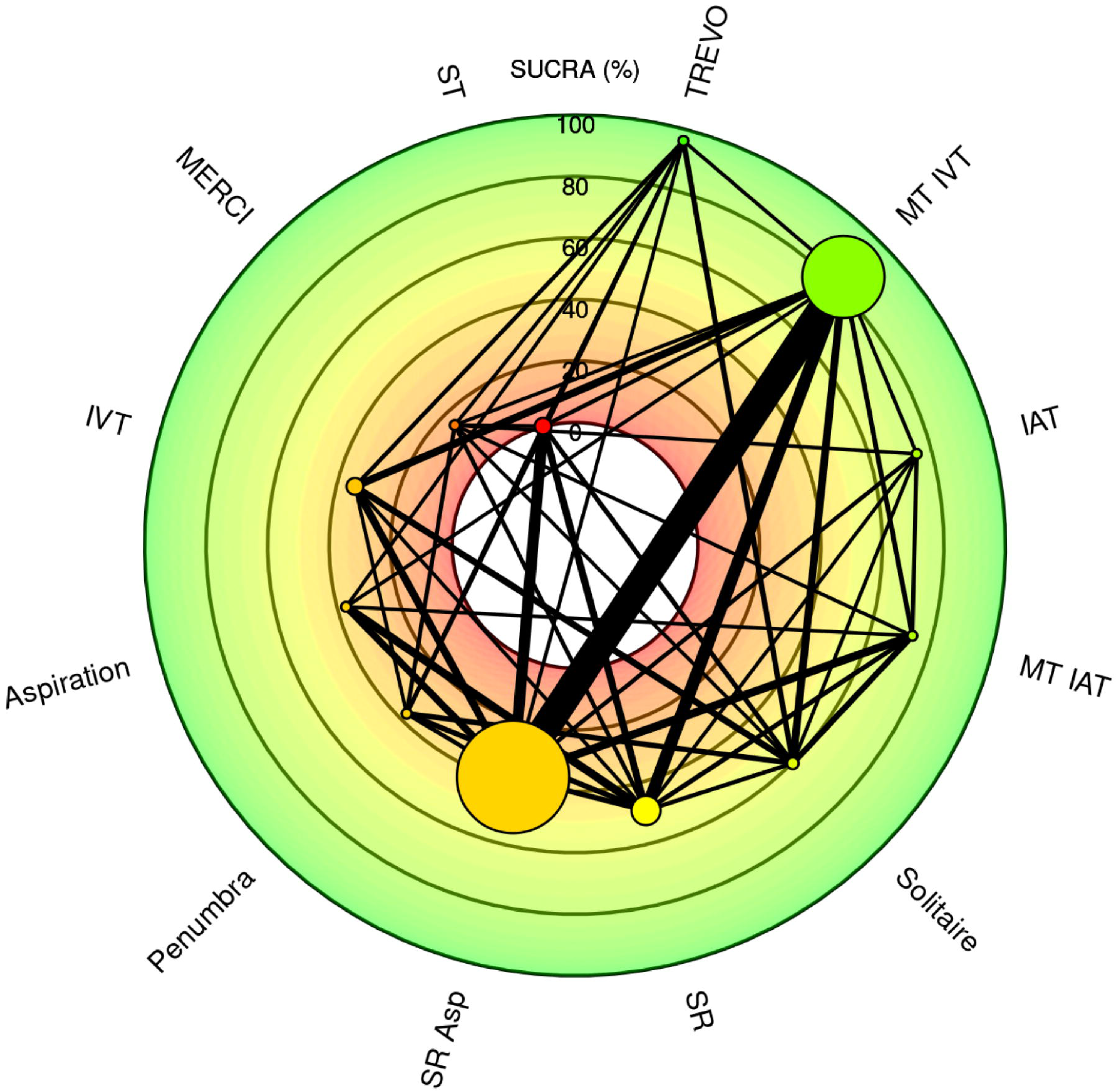
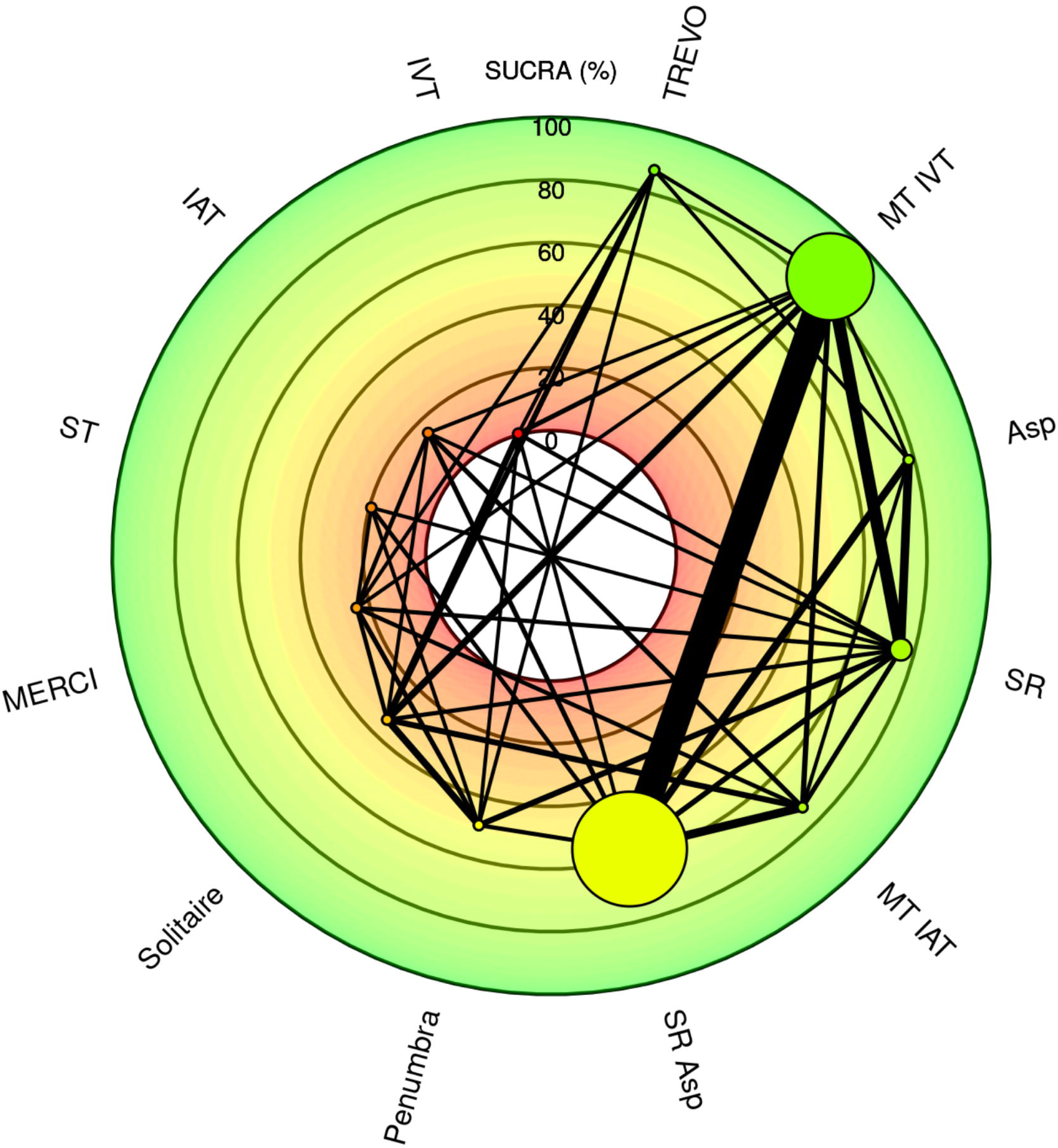
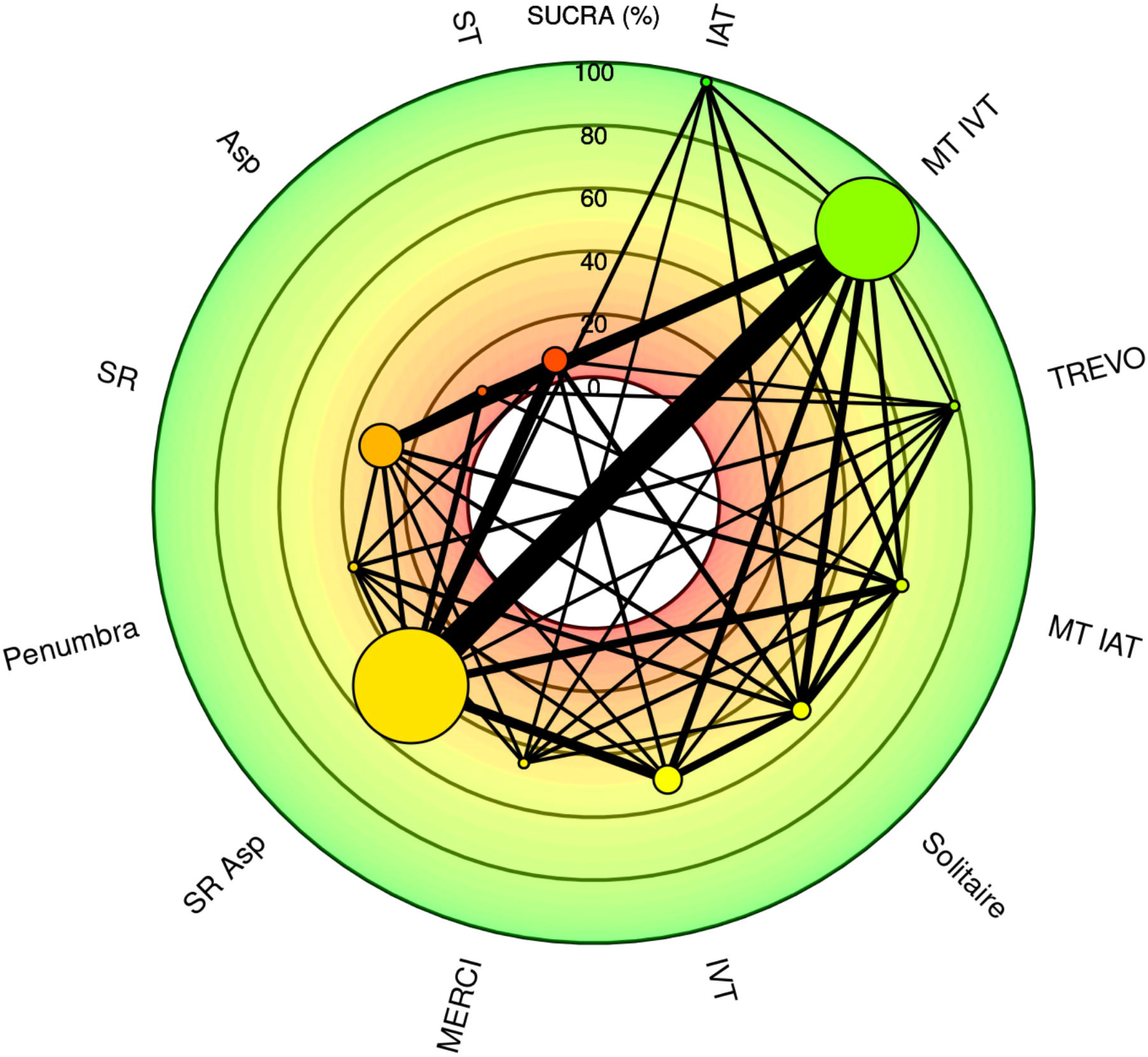
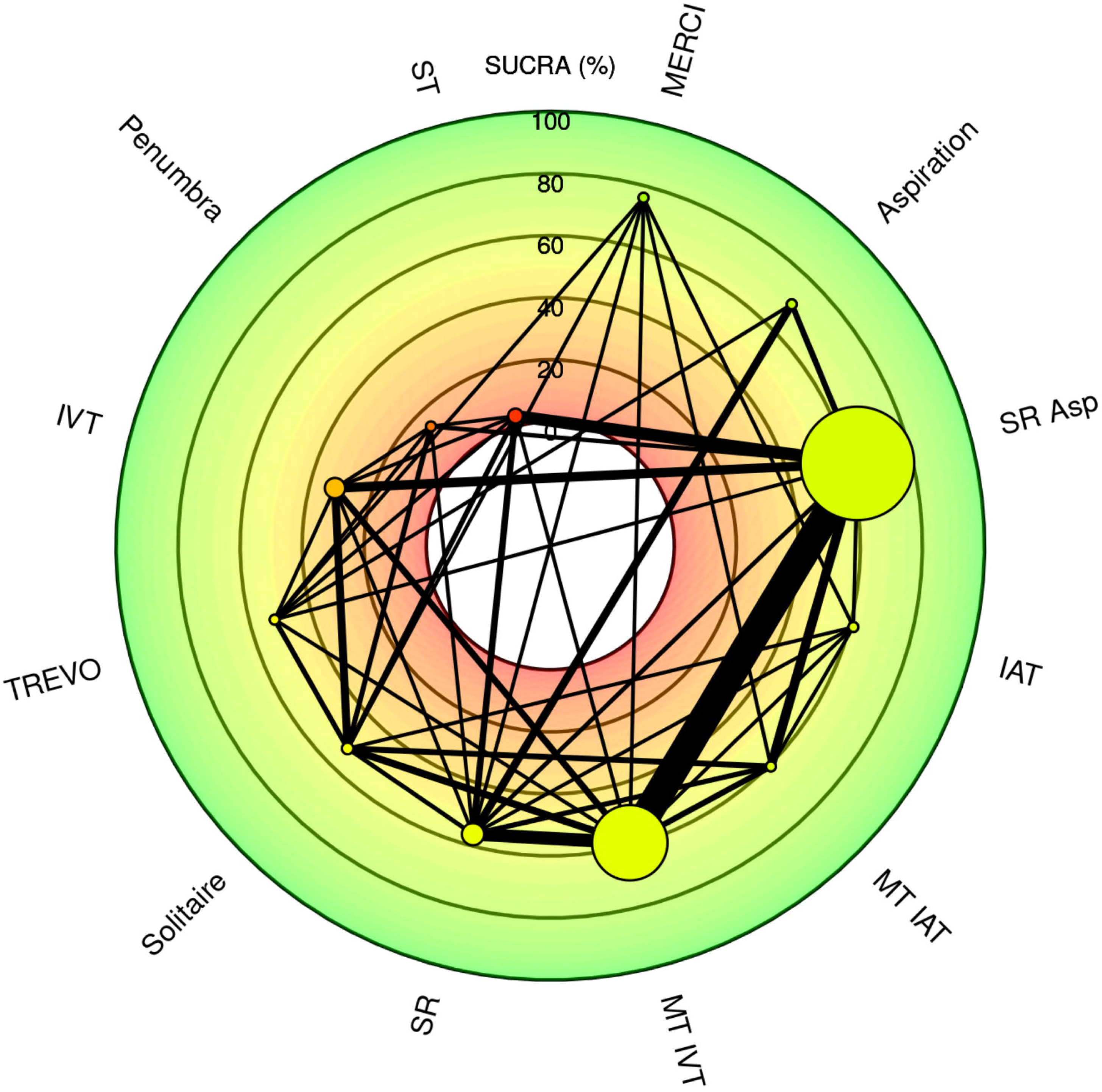
**(a-d)**: Radial SUCRA plot for (a) good functional recovery at 90 days; (b) successful recanalization; (c) all-cause mortality at 90 days and (d) sICH within 24 hours.

**Figure 5.**
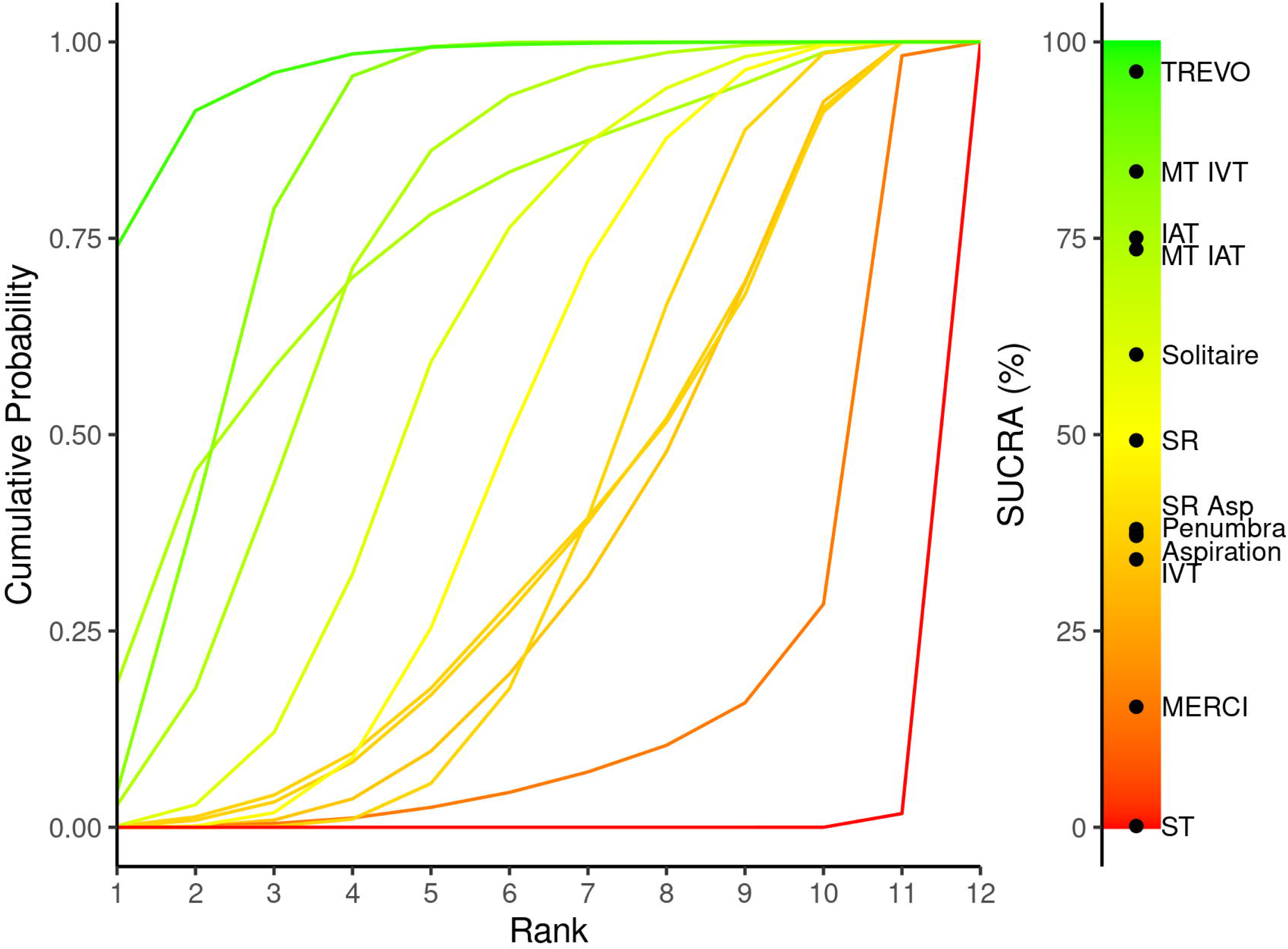
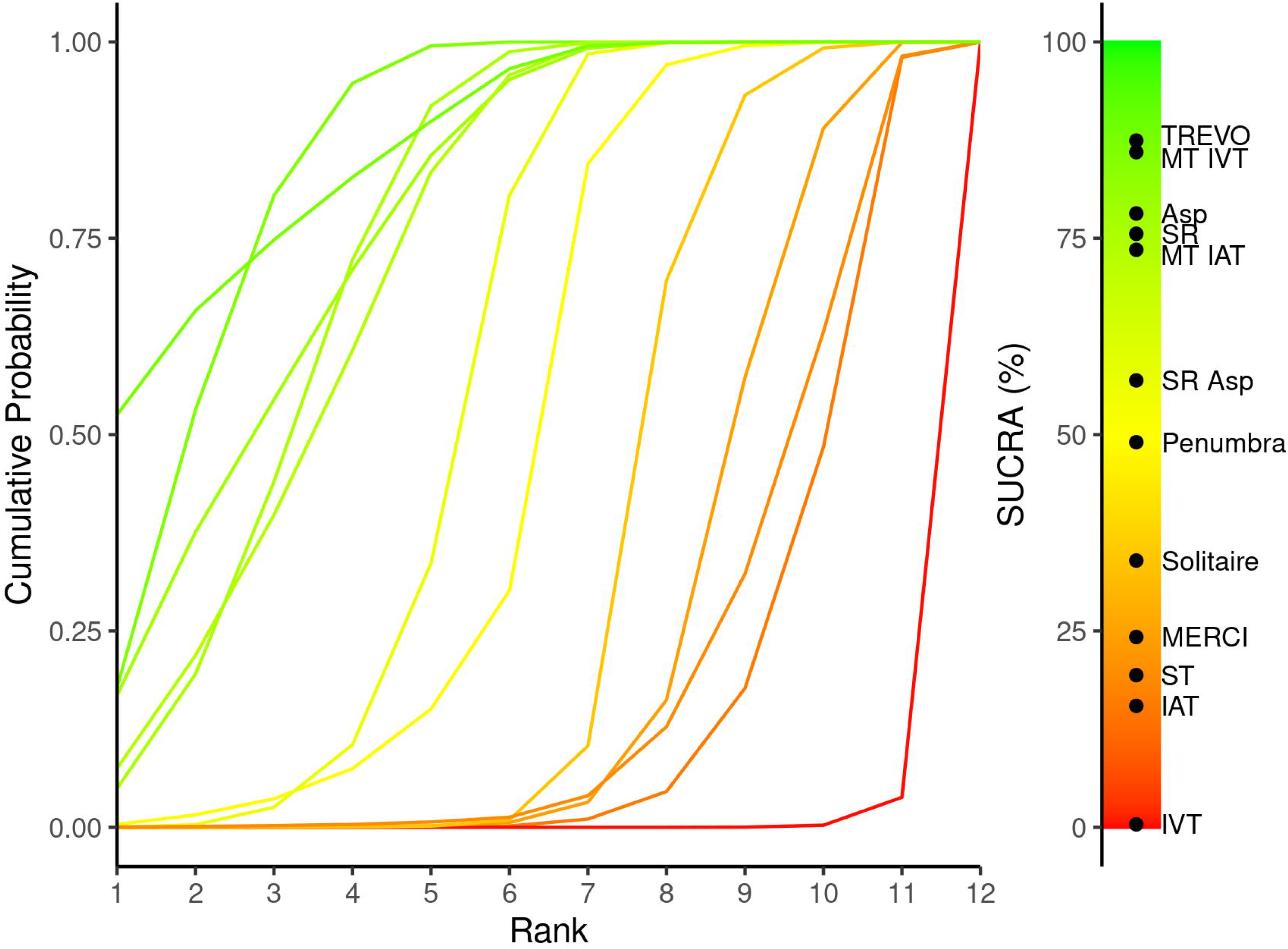
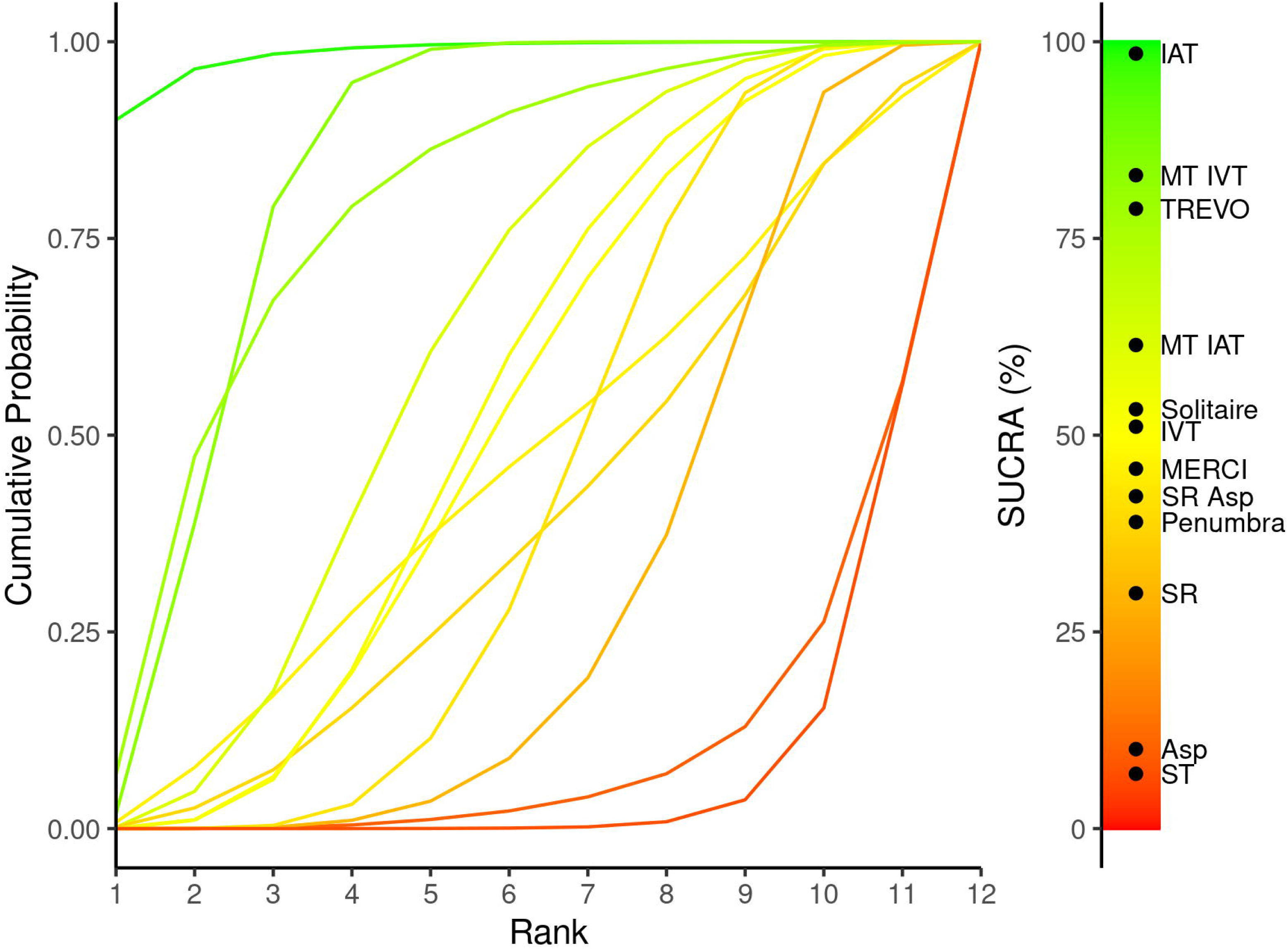
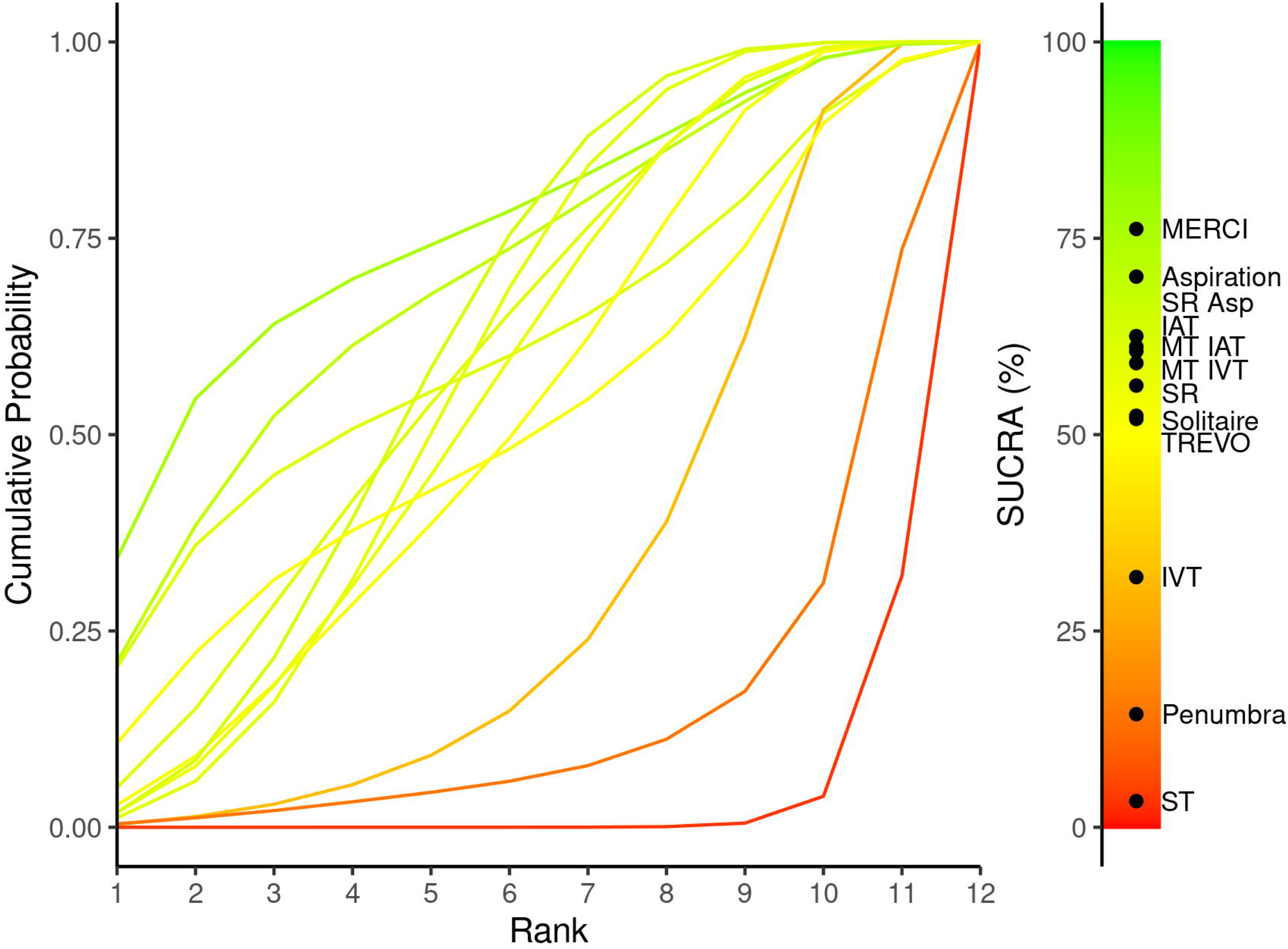
**(a-d)**: Litmus Rank-O-Gram for (a) good functional recovery at 90 days; (b) successful recanalization; (c) all-cause mortality at 90 days and (d) sICH within 24 hours.

SUCRA plot for successful recanalization shows that TREVO has the highest SUCRA value, indicating it as the most effective device. MT IVT also performs well, while Solitaire and Penumbra show lower effectiveness. For all-cause mortality at 90 days, MT IVT leads with the highest SUCRA value, suggesting the greatest reduction in mortality. TREVO and MT IAT also rank high, whereas Penumbra and Solitaire are less effective. The plot for sICH within 24 hours highlights SR+Asp as the safest option with the highest SUCRA value, indicating the lowest risk of sICH. MT IAT also shows low risk, while Solitaire, and IVT are associated with higher sICH risk.

## Discussion

Our NMA demonstrated that the TREVO device emerged as the most effective for improving functional recovery, successful recanalization, and reducing mortality rates in patients with AIS caused by LVO. Conversely, devices such as MERCI and Solitaire were associated with a higher incidence of sICH. This highlights a critical balance between efficacy and safety that must be considered when selecting an MT device. To the best of our knowledge, this is the largest NMA on this topic synthesizing data from 71,154 AIS patients pooled from 201 studies.

For patients with AIS caused by LVO, previous RCTs have indicated that standard medical treatment or thrombolysis is inferior to MT. There is still ongoing debate over which MT devices offer the best outcomes in terms of functional recovery, lower mortality, reduced incidence of sICH, and higher recanalization success rates. A previous NMA by Deng et al.^23^ included five RCTs and five observational studies, totalling only 1,659 AIS subjects. Their findings indicated that both Solitaire and Trevo devices were associated with higher rates of functional independence compared to the Penumbra device. Additionally, Solitaire and Trevo demonstrated superior recanalization success rates compared to the Merci device. However, no significant differences were observed among the devices when assessing sICH and mortality rates.

In some instances, combining stent retrievers and aspiration techniques may be the most effective approach. The COMPASS trial,^24^ which randomized patients to receive either catheter aspiration or stent retriever treatment, found no significant difference in outcomes, suggesting that aspiration-first treatment is noninferior to stent retriever-first treatment. Additionally, the trial reported no significant differences in mortality, sICH, or other safety concerns between the two methods. Although some data suggest that combining stent retrievers with aspiration may offer slight advantages in achieving near-total or total reperfusion, these findings did not reach statistical significance. A recent systematic review and meta-analysis by Zaidat et al.^25^ suggests that the EmboTrap device may lead to significantly better functional outcomes compared to Solitaire and Trevo. Additionally, EmboTrap and Trevo were associated with lower rates of sICH and mortality than Solitaire. They included single-arm study, which can introduce biases based on population and procedural differences. Moreover, their study was lack of multi-arm RCTs which limits the ability to perform a network meta-analysis.

Our findings align with previous analyses, such as the NMA by Deng et al. (2019)^23^, which also highlighted the superiority of the Solitaire and Trevo devices over the Penumbra device in terms of functional independence. In our analysis, Solitaire and Trevo consistently demonstrated higher rates of successful recanalization compared to other devices, including Merci and Penumbra. This supports the growing consensus that stent retrievers like Solitaire and Trevo are among the most effective options for achieving favourable functional outcomes and high recanalization success rates in patients with AIS due to LVO.

Interestingly, despite the advantages in functional recovery and recanalization rates, our study, like previous analyses by Barral et al. (2018)^26^ and Zaidat et al. (2023),^25^ found no significant differences in mortality or the occurrence of sICH across various devices. This suggests that while certain devices may excel in recanalization and functional recovery, the risks associated with sICH and mortality are likely influenced by other factors, such as patient characteristics and procedural variables, rather than the choice of device alone.

Moreover, our analysis corroborates findings from studies by Ahmed et al. (2023)^11^ and Rajkumar et al. (2022),^27^ which emphasize the importance of considering both stent retrievers and aspiration techniques. While stent retrievers such as Solitaire and Trevo remain the gold standard in many clinical scenarios, emerging evidence suggests that aspiration techniques can offer competitive outcomes, particularly in specific patient subgroups. This reinforces the need for personalized treatment approaches, where the choice of device or combination of devices is tailored to the individual patient’s anatomical and clinical characteristics.

It is also worth noting that, similar to findings by Campbell et al. (2016)^28^ and Gory et al. (2018),^29^ the effectiveness of MT devices is highly dependent on operator experience and institutional protocols. Thus, ongoing training and refinement of endovascular techniques are crucial for optimizing patient outcomes across different healthcare settings.

Our study does have some limitations. First, we were unable to compare specific devices directly because many of the included studies used mixed devices and reported outcomes without specifying which device was used, while some studies only mentioned MT without naming the device. Second, the scales used to measure outcomes, such as sICH and successful recanalization, varied across the included studies. Third, the time window for enrolling AIS patients ranged from 3 to 24 hours after symptom onset, which may have influenced the outcomes. Fourth, we excluded a number of studies that only conducted single-arm comparisons, focusing solely on studies that included two or more arms for comparison. Additionally, although our analysis covered global studies from major regions like USA and Europe, studies on Asian populations were relatively rare. Lastly, we included both RCTs and cohort studies to provide a more comprehensive answer, given the limited number of published RCTs on this topic.

## Conclusion

Our findings demonstrate contemporary stent-retriever device technology as the most effective option for improving functional recovery, recanalization success, and reducing mortality in AIS patients. These results highlight the critical need for selecting the most effective and safest thrombectomy device to optimize outcomes in acute stroke care.

## Declaration

### Author contributions

AG & MS: Study screening, Data curation, MN & SM: quality assessment & validation, KKP & ADS: Formal analysis, DV & SD; writing, reviewing & editing. PK: Conceptualization, Formal analysis, Writing – original draft

### Data availability statement

The original contributions presented in the study are included in the article/Supplementary material, further inquiries can be directed to the corresponding author.

### Ethics statement

Ethical approval and informed consent were not required for this systematic review and meta-analysis.

## Supporting information

PRISMA 2020 Chcklist

## Acknowledgement

None

---

**Table-eS1:** Summary of Search Results Across Databases with Search Key Terms and results.

| **Databases** | **Search Key Terms** | **Search Results** |
| --- | --- | --- |
| **PubMed** | ("acute ischemic stroke"[All Fields] OR "acute stroke"[All Fields]) OR "large vessel occlusion") AND ("mechanical thrombectomy"[All Fields] OR "endovascular therapy"[All Fields] OR "endovascular treatment"[All Fields] OR "thrombectomy devices"[All Fields] OR ("retriever"[All Fields] OR "retrievers"[All Fields]) OR ("stent s"[All Fields] OR "stentings"[All Fields] OR "stents"[MeSH Terms] OR "stents"[All Fields] OR "stent"[All Fields] OR "stented"[All Fields] OR "stenting"[All Fields]) OR ("aspirant"[All Fields] OR "aspirants"[All Fields] OR "aspirate"[All Fields] OR "aspirates"[All Fields] OR "aspirating"[All Fields] OR "aspirational"[All Fields] OR "aspirations, psychological"[MeSH Terms] OR ("aspirations"[All Fields] AND "psychological"[All Fields]) OR "psychological aspirations"[All Fields] OR "aspirations"[All Fields] OR "aspirative"[All Fields] OR "aspirator"[All Fields] OR "aspirators"[All Fields] OR "aspire"[All Fields] OR "aspired"[All Fields] OR "aspires"[All Fields] OR "aspiring"[All Fields] OR "direct aspiration"[All Fields] OR "contact aspiration"[All Fields]) OR "Intravenous thrombolysis "OR "IVT"[All Fields] OR "Intra-arterial Therapy "OR "IAT"[All Fields] OR "standard treatment"[All Fields])AND ("recanalization"[All Fields] OR "symptomatic intracranial hemorrhage"[All Fields] OR ("good"[All Fields] AND ("recovery of function"[MeSH Terms] OR ("recovery"[All Fields] AND "function"[All Fields]) OR "recovery of function"[All Fields] OR ("functional"[All Fields] AND "recovery"[All Fields]) OR "functional recovery"[All Fields]) AND ("MERCi"[All Fields] OR "TREVO"[All Fields] OR "Solitaire"[All Fields] OR ("penumbra"[All Fields] | 6,438 |
| **EMBASE** | ("acute ischemic stroke" OR "acute stroke" OR "large vessel occlusion") AND ("mechanical thrombectomy" OR "endovascular therapy" OR "endovascular treatment" OR "thrombectomy devices" OR retriever OR stent OR aspiration OR "direct aspiration" OR "contact aspiration" OR "Intravenous thrombolysis "OR "IVT OR "Intra-arterial Therapy "OR "IAT" OR "standard treatment" AND (MERCi OR TREVO OR Solitaire OR Penumbra) | 739 |
| **Google Scholar** | ("acute ischemic stroke" OR "acute stroke" OR "large vessel occlusion") AND ("mechanical thrombectomy" OR "endovascular therapy" OR "endovascular treatment" OR "thrombectomy devices" OR retriever OR stent OR aspiration OR "direct aspiration" OR "contact aspiration" OR "Intravenous thrombolysis "OR "IVT OR "Intra-arterial Therapy "OR "IAT" OR "standard treatment" AND (MERCi OR TREVO OR Solitaire OR Penumbra) | 895 |
| **Cochrane Library** | ("acute ischemic stroke" OR "acute stroke" OR "large vessel occlusion") AND ("mechanical thrombectomy" OR "endovascular therapy" OR "endovascular treatment" OR "thrombectomy devices" OR retriever OR stent OR aspiration OR "direct aspiration" OR "contact aspiration" OR "Intravenous thrombolysis "OR "IVT OR "Intra-arterial Therapy "OR "IAT" OR "standard treatment" AND (MERCi OR TREVO OR Solitaire OR Penumbra) | 509 |
| **Limits to:** Humans |  |  |

**Table eS2:**
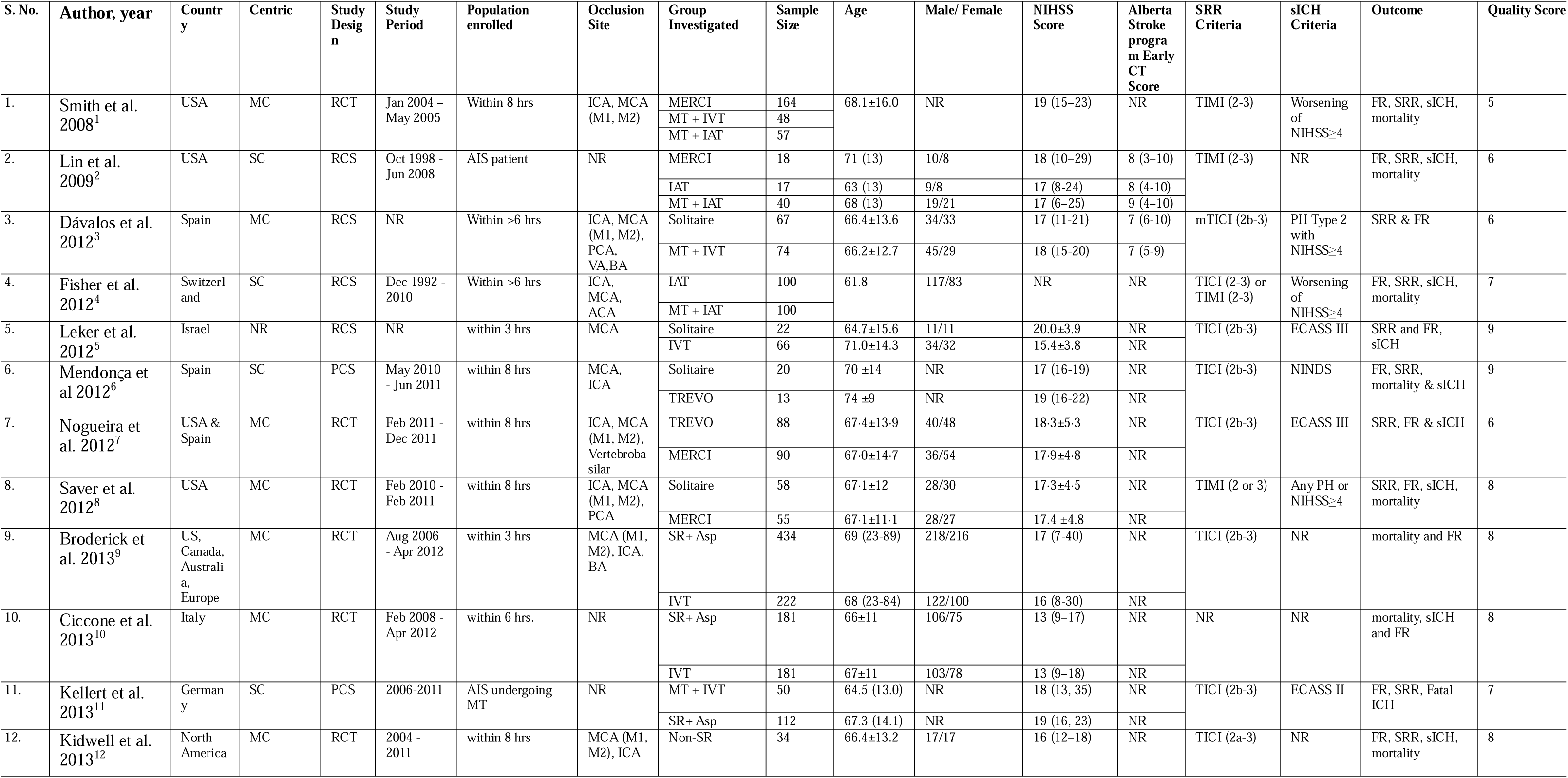
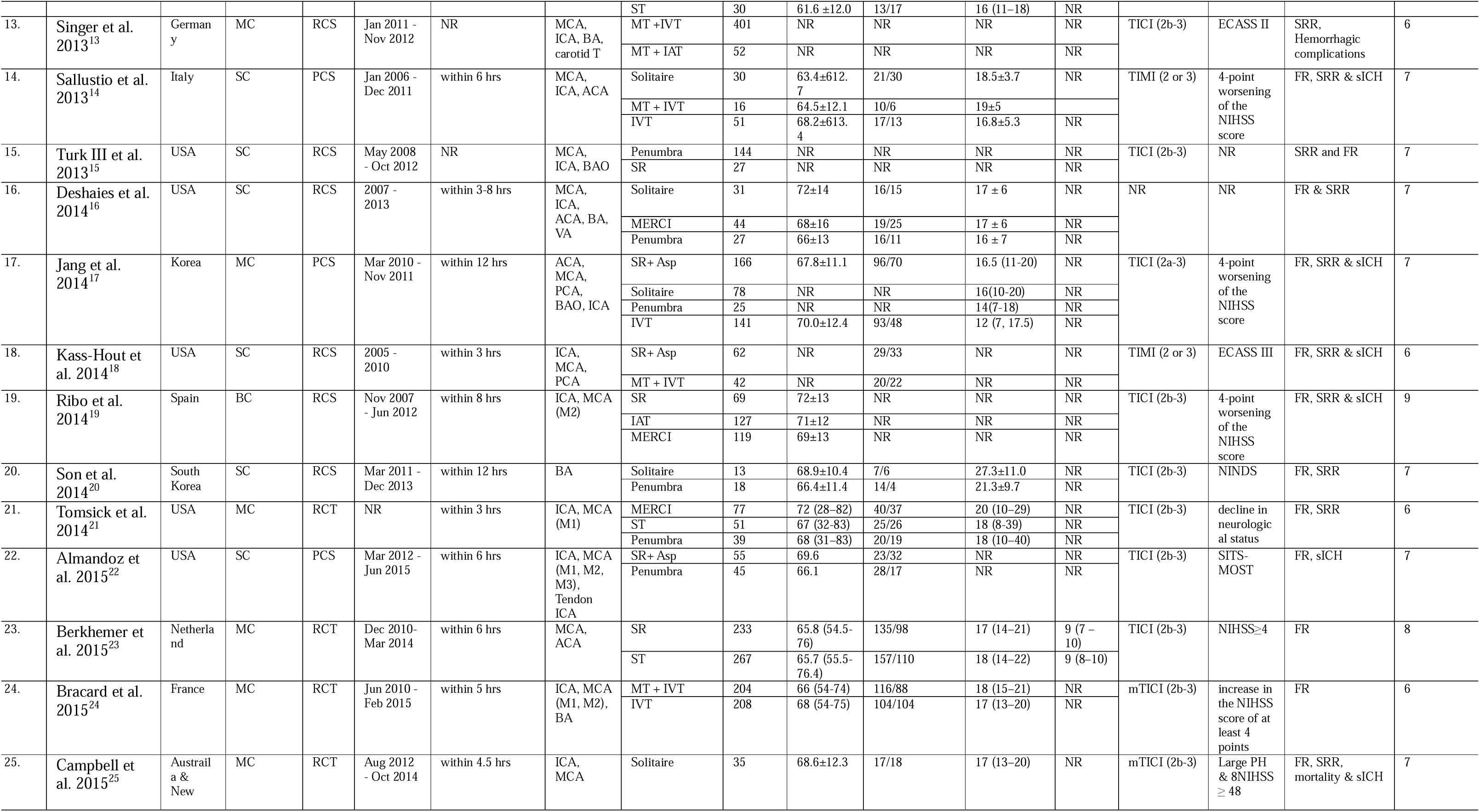
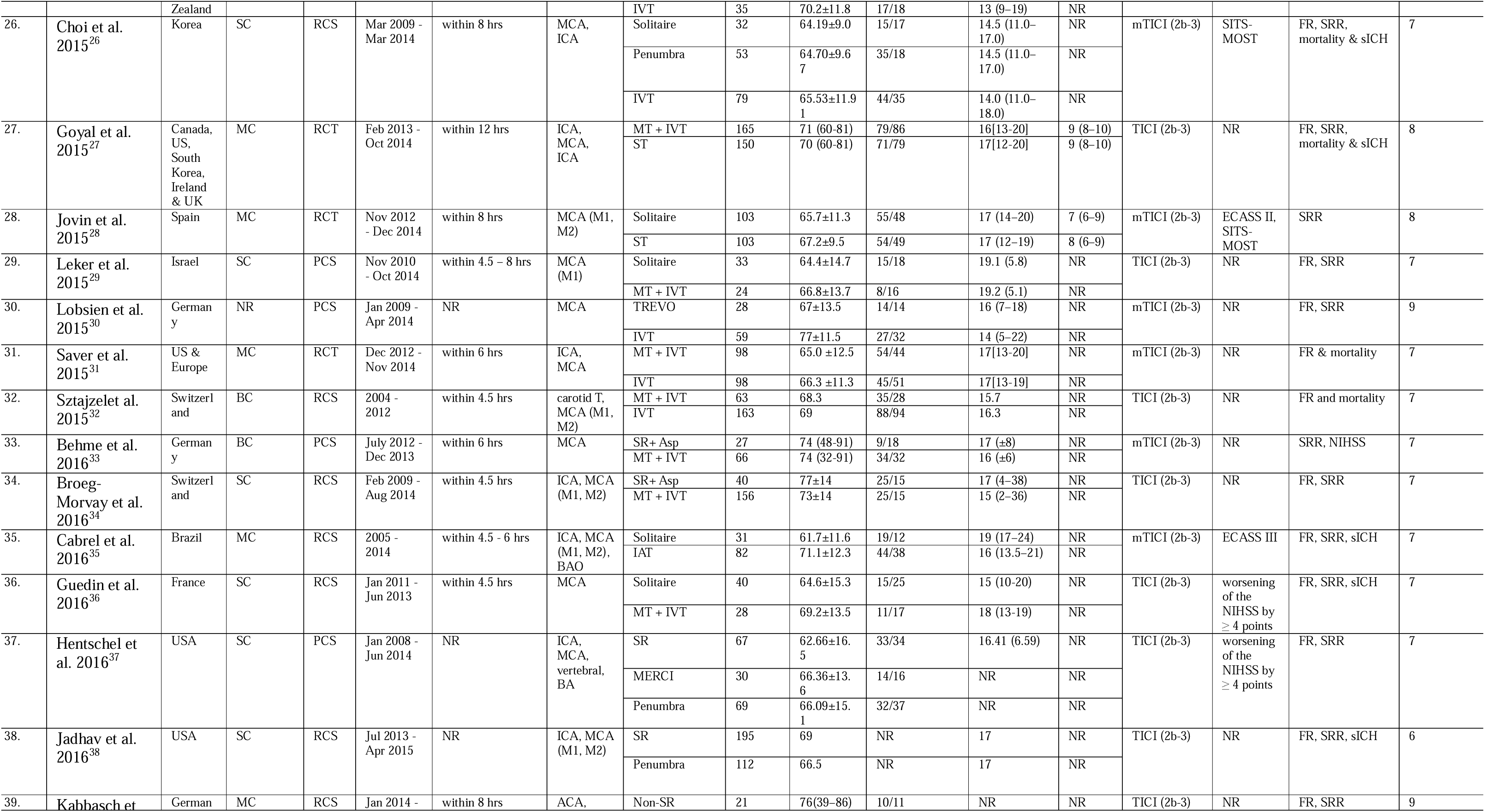
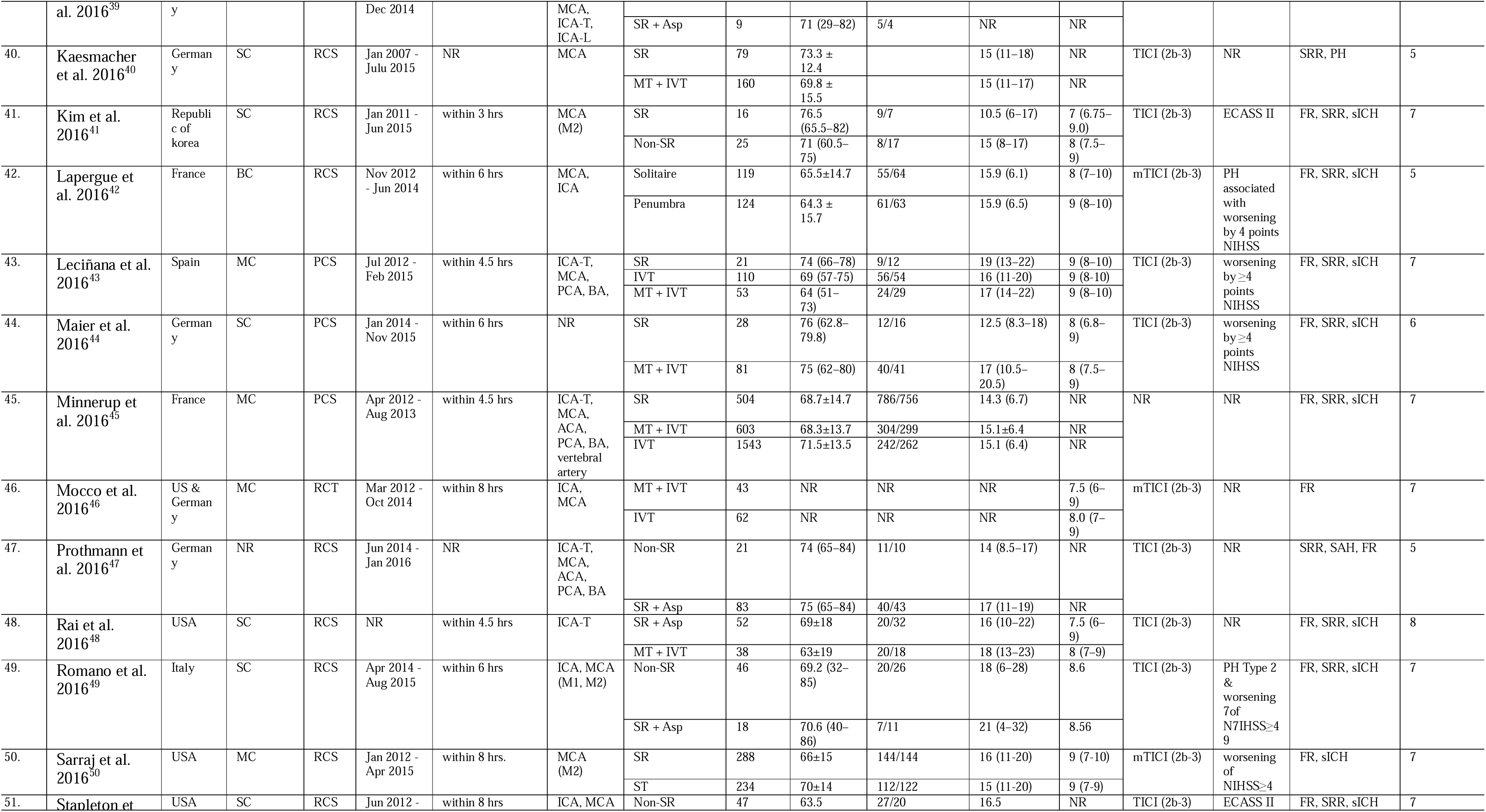
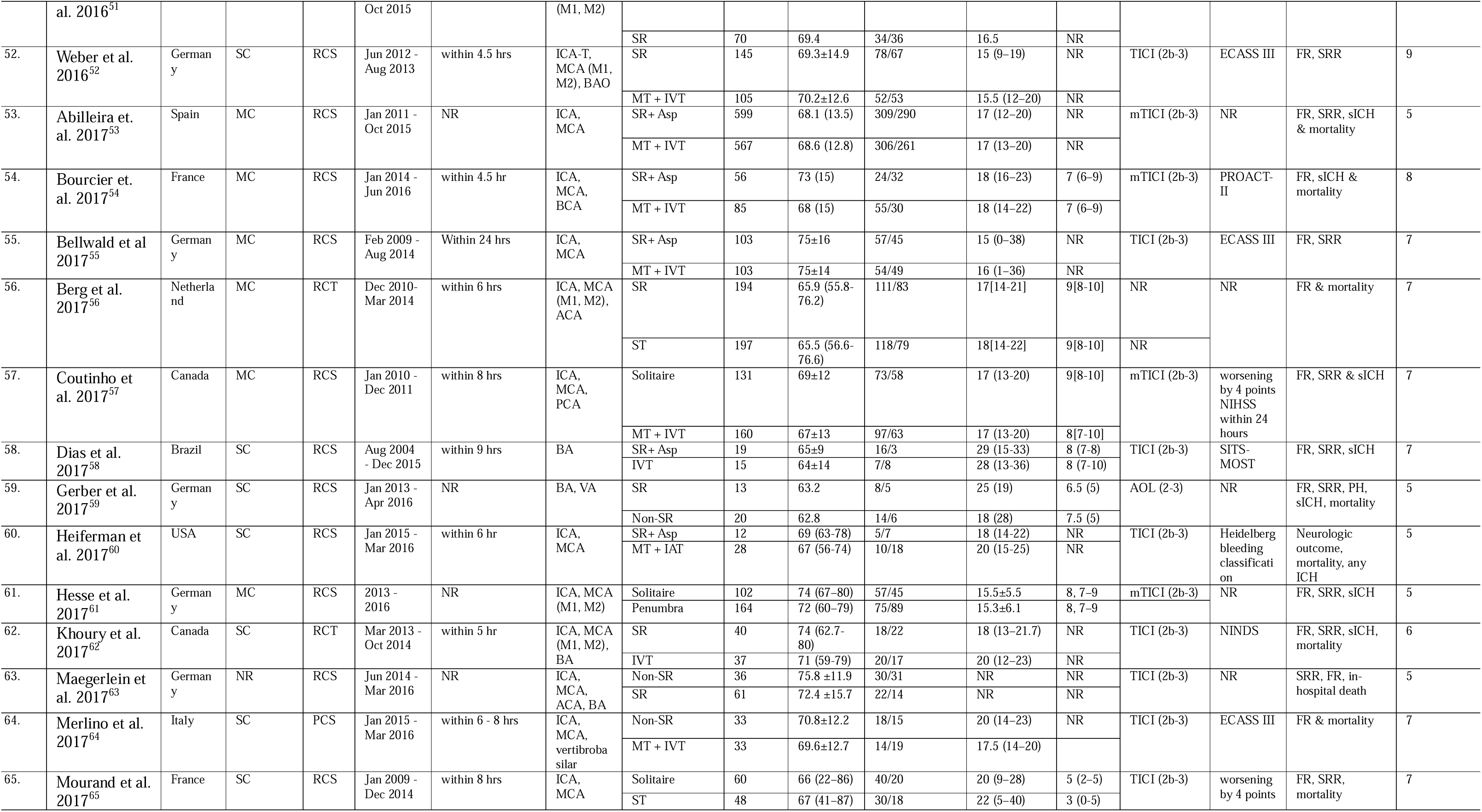
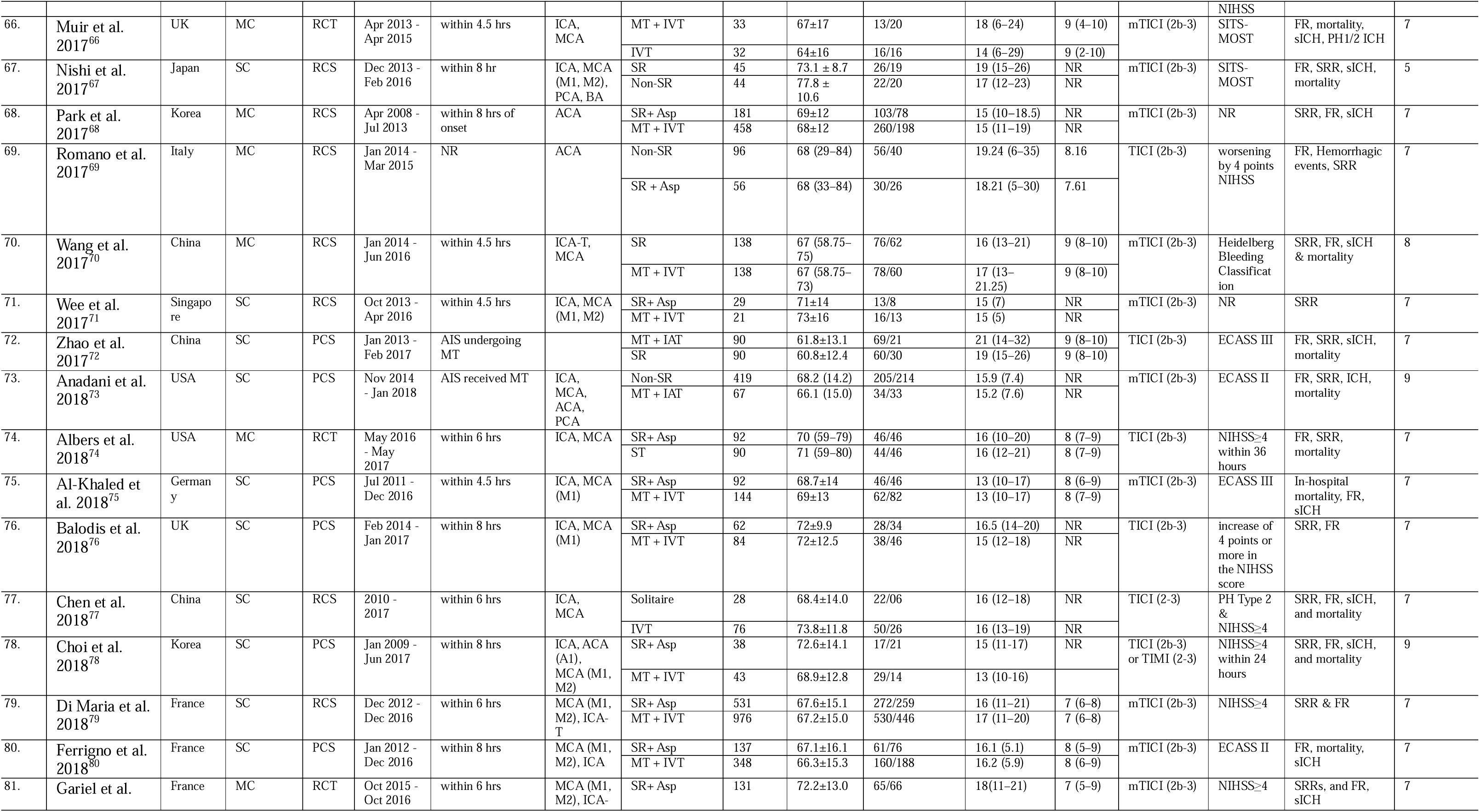
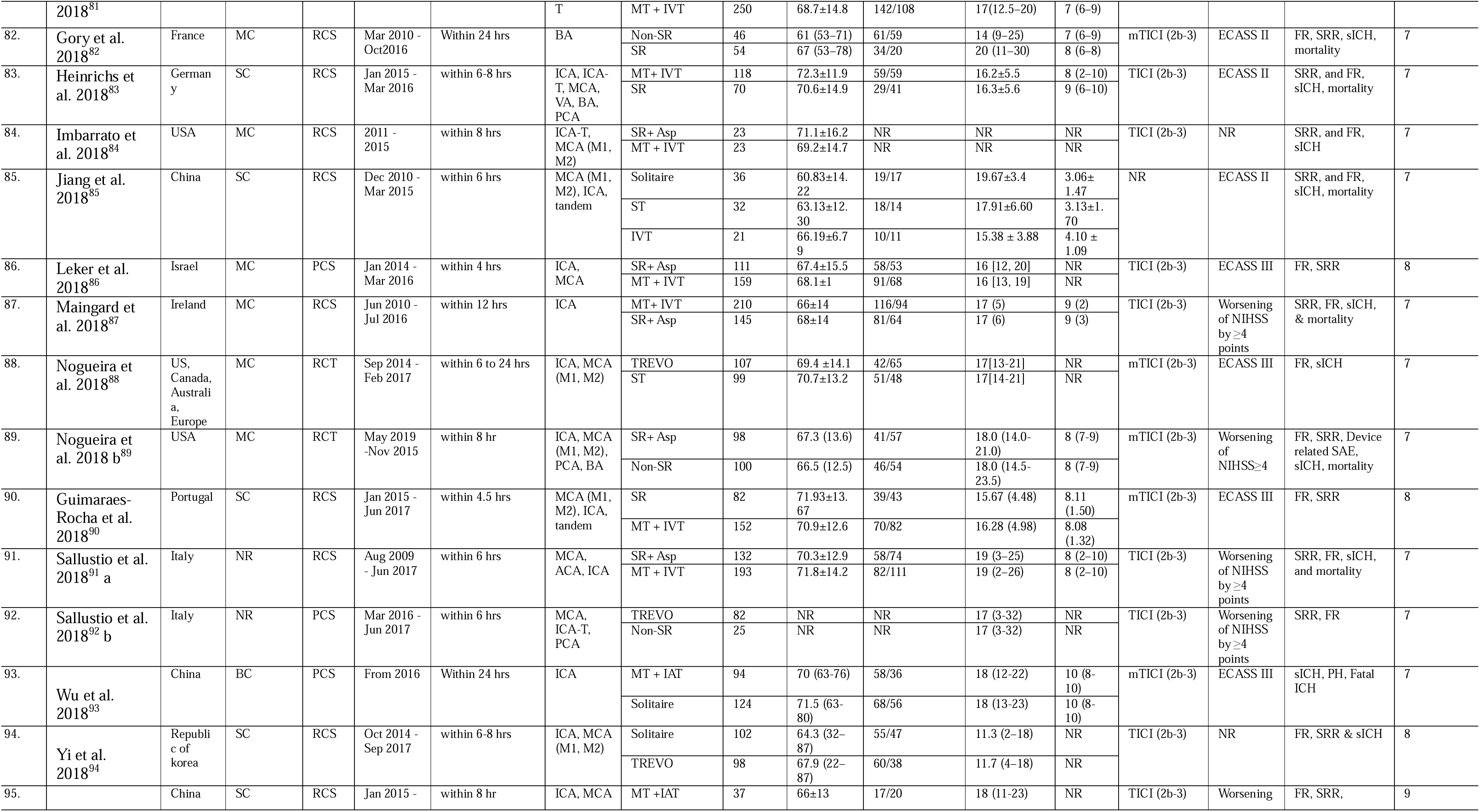
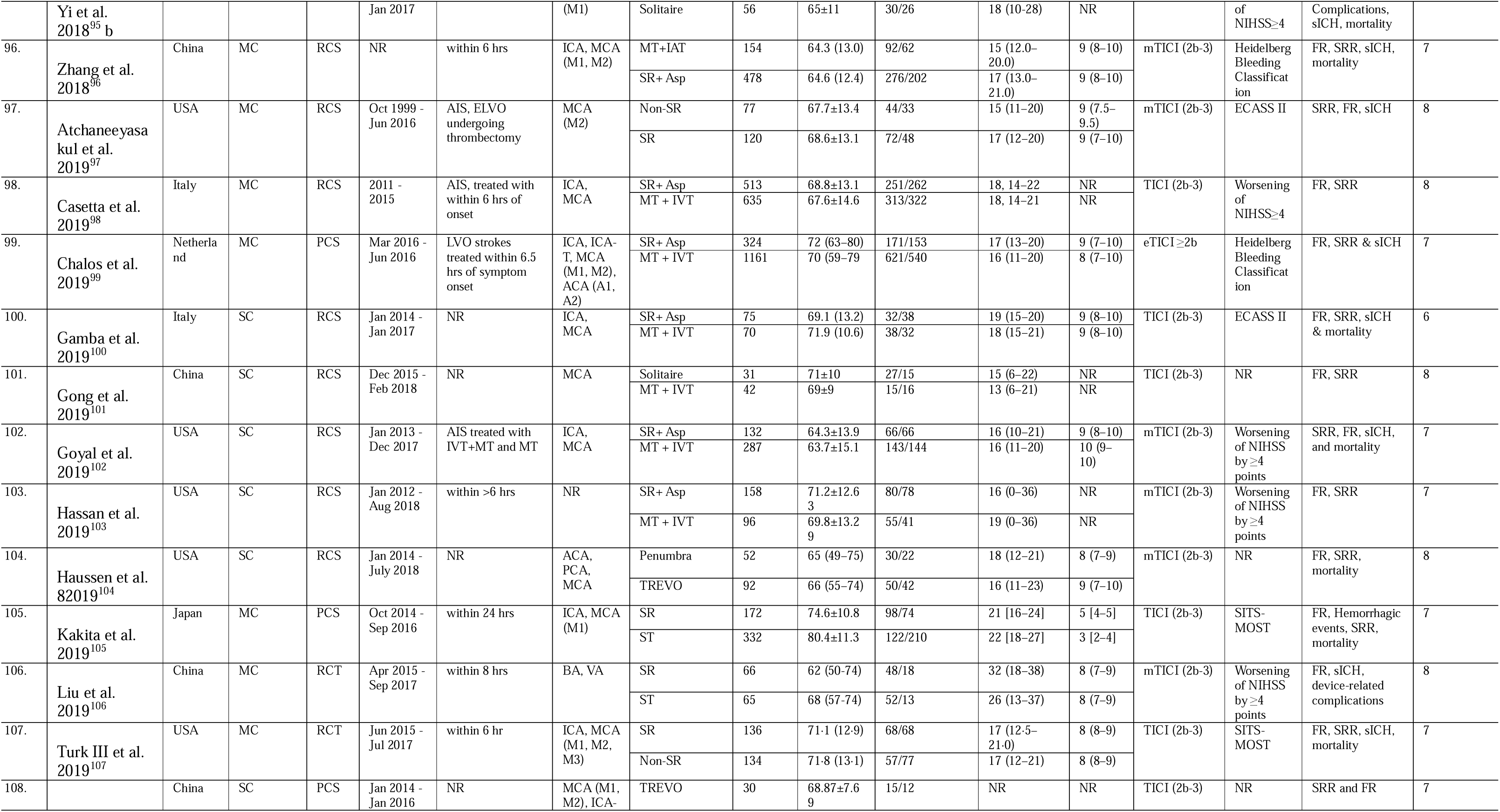
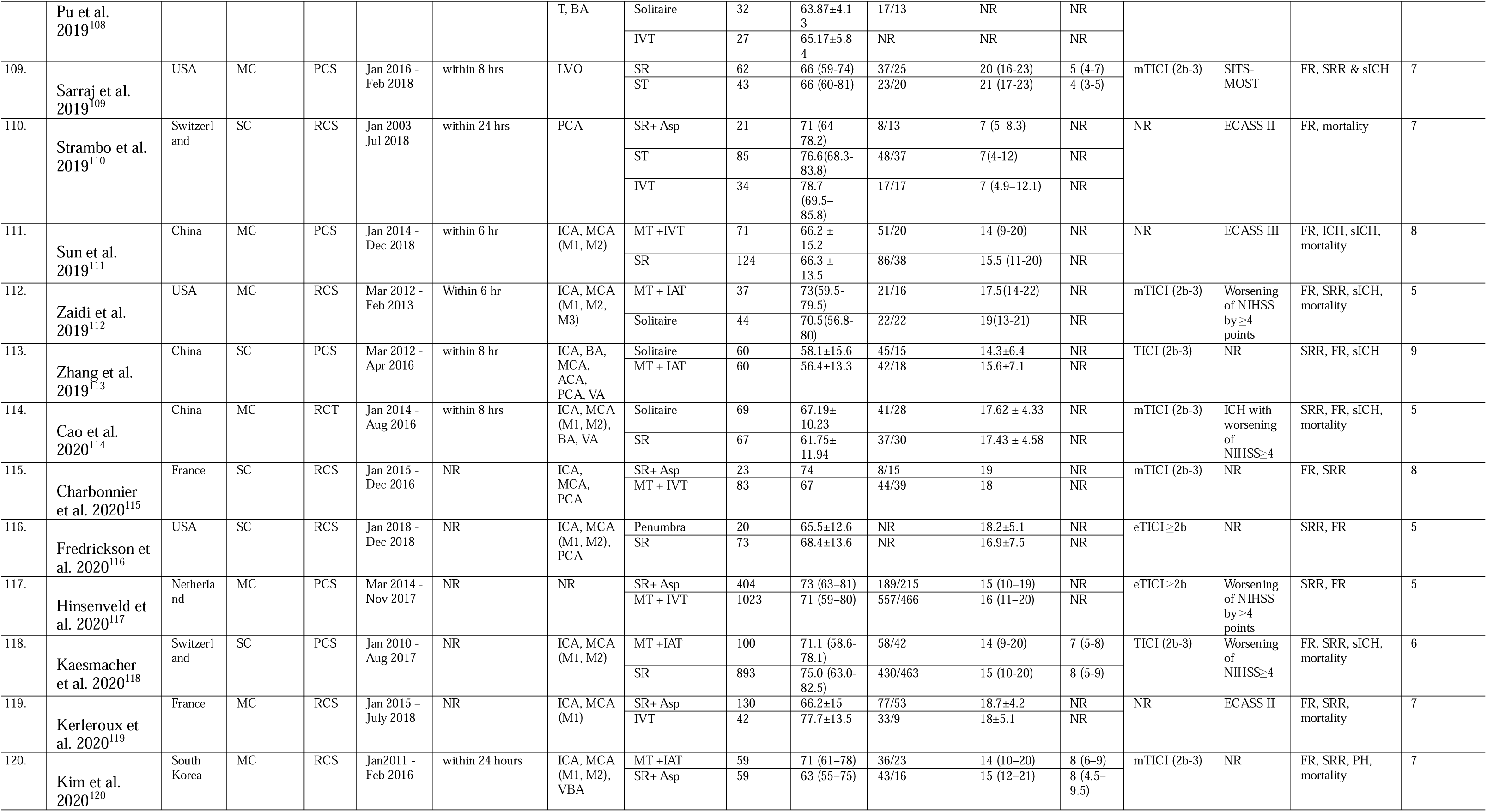
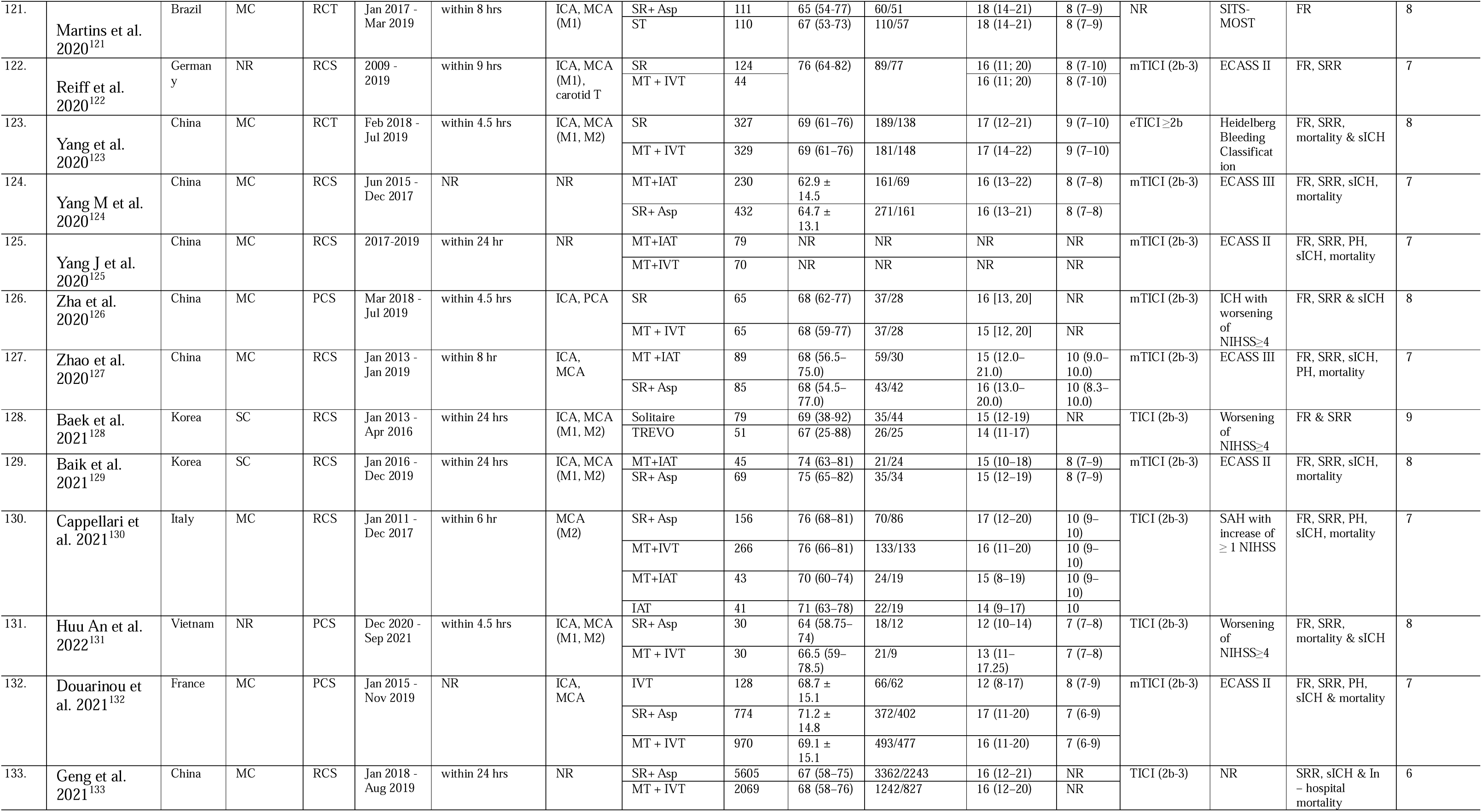
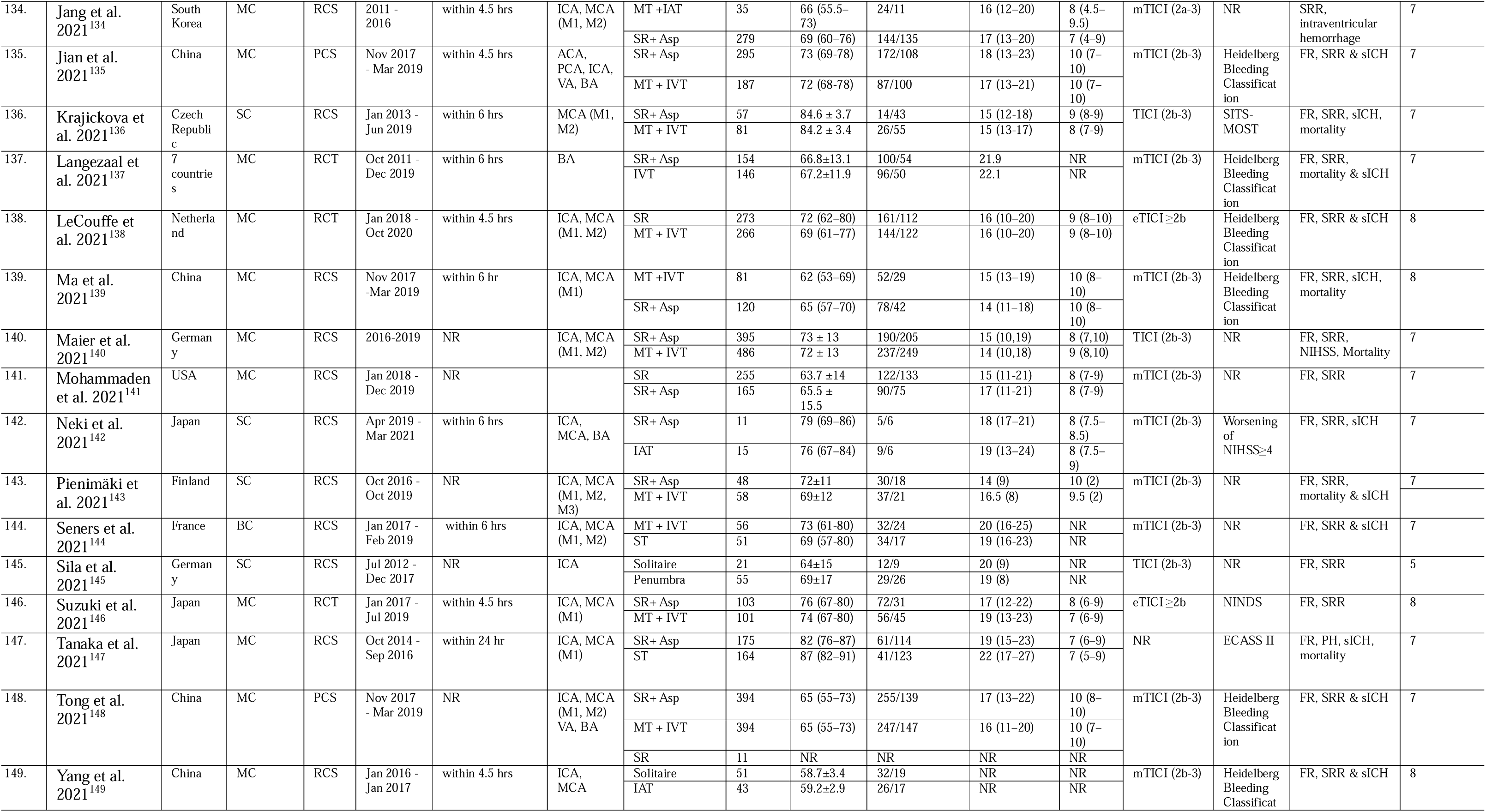
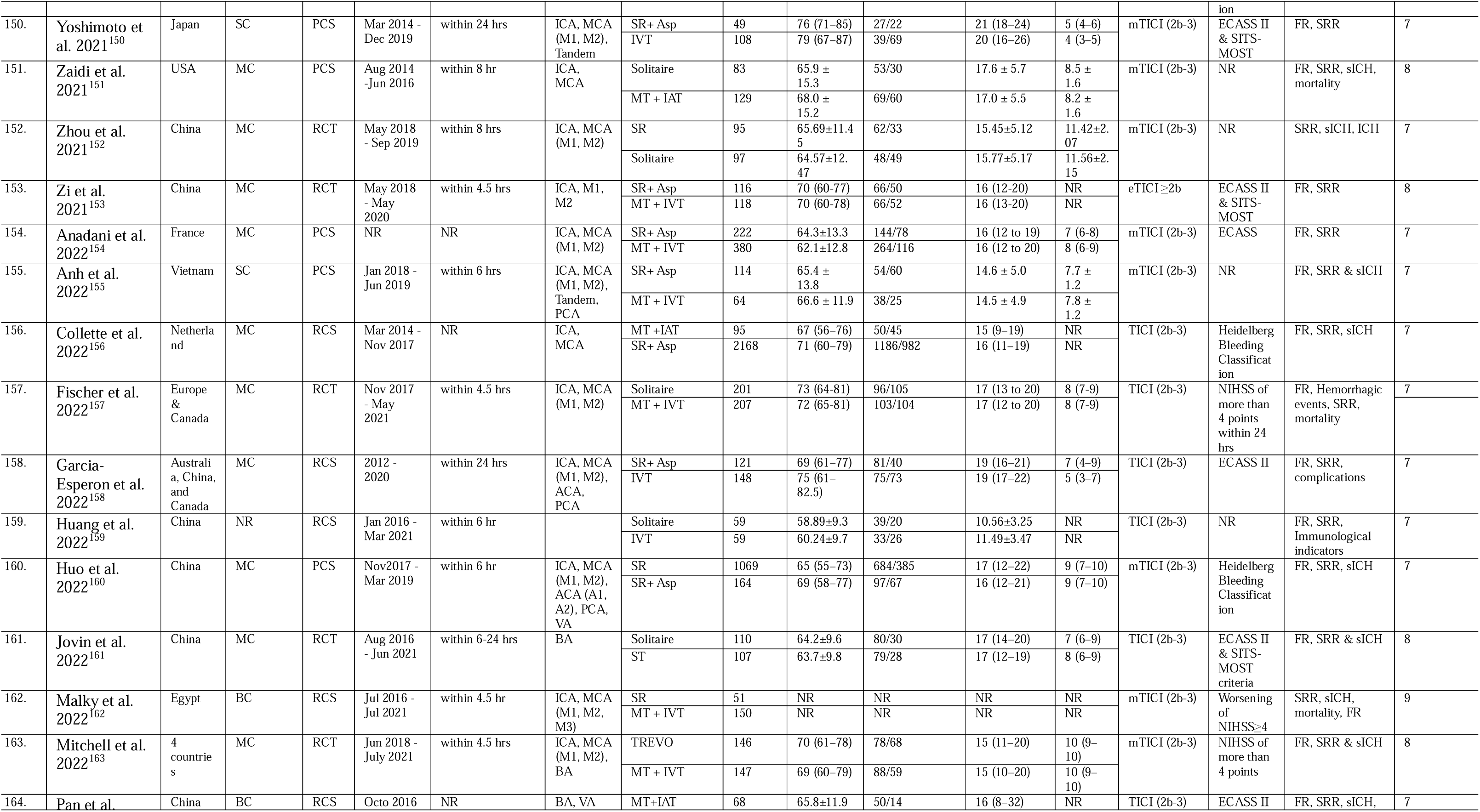
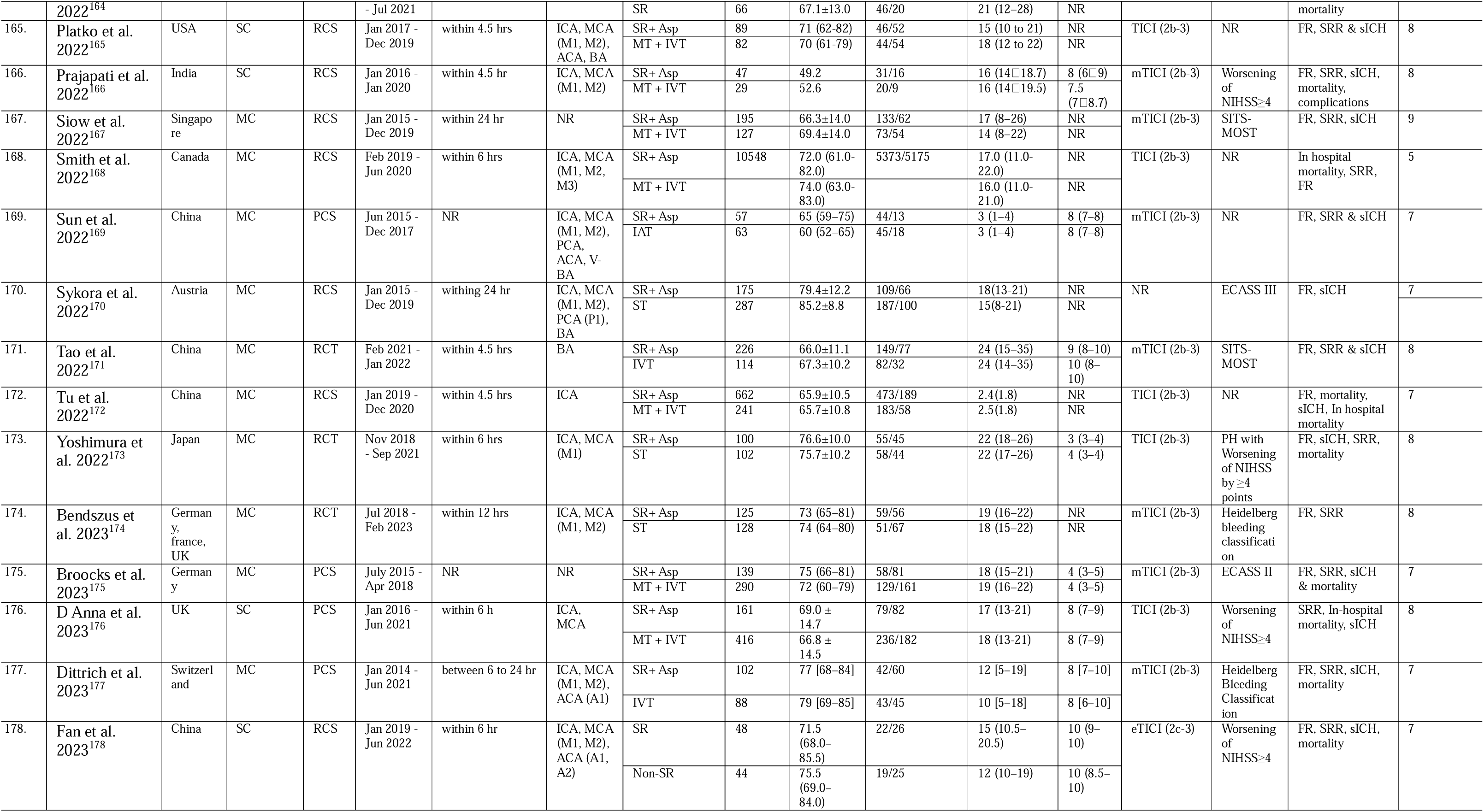
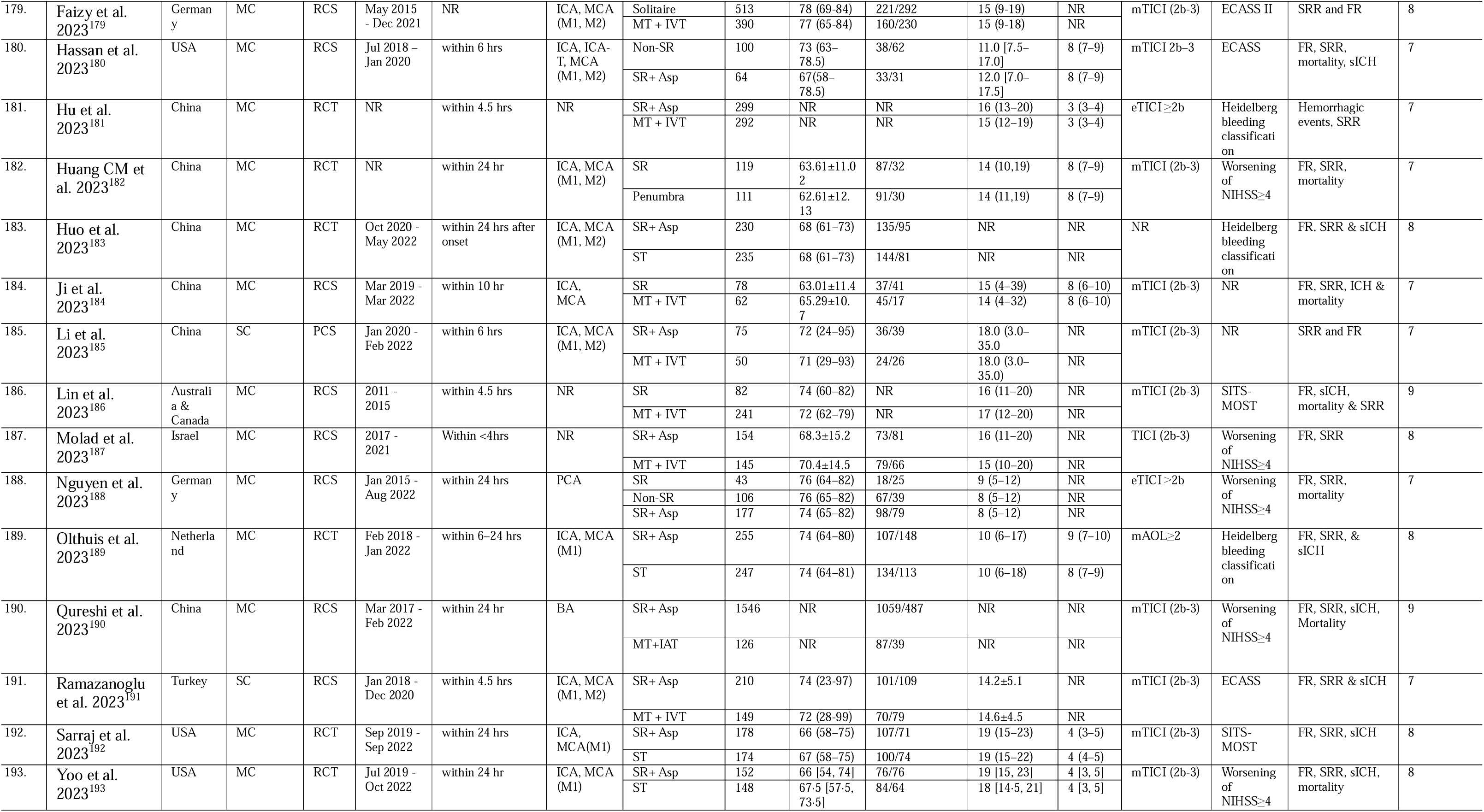
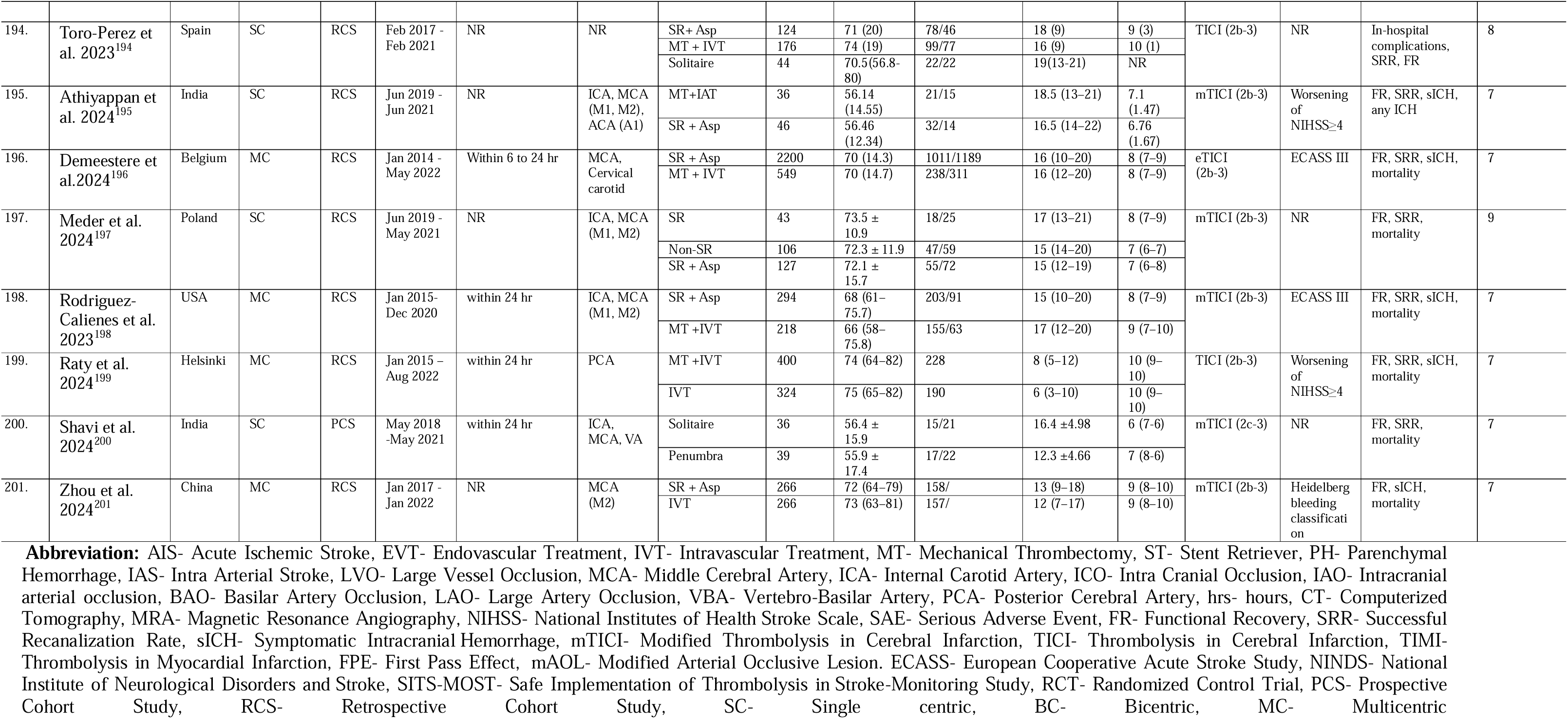
Baseline characteristics for the included studies in the network meta-analysis.

**Table-eS3:**
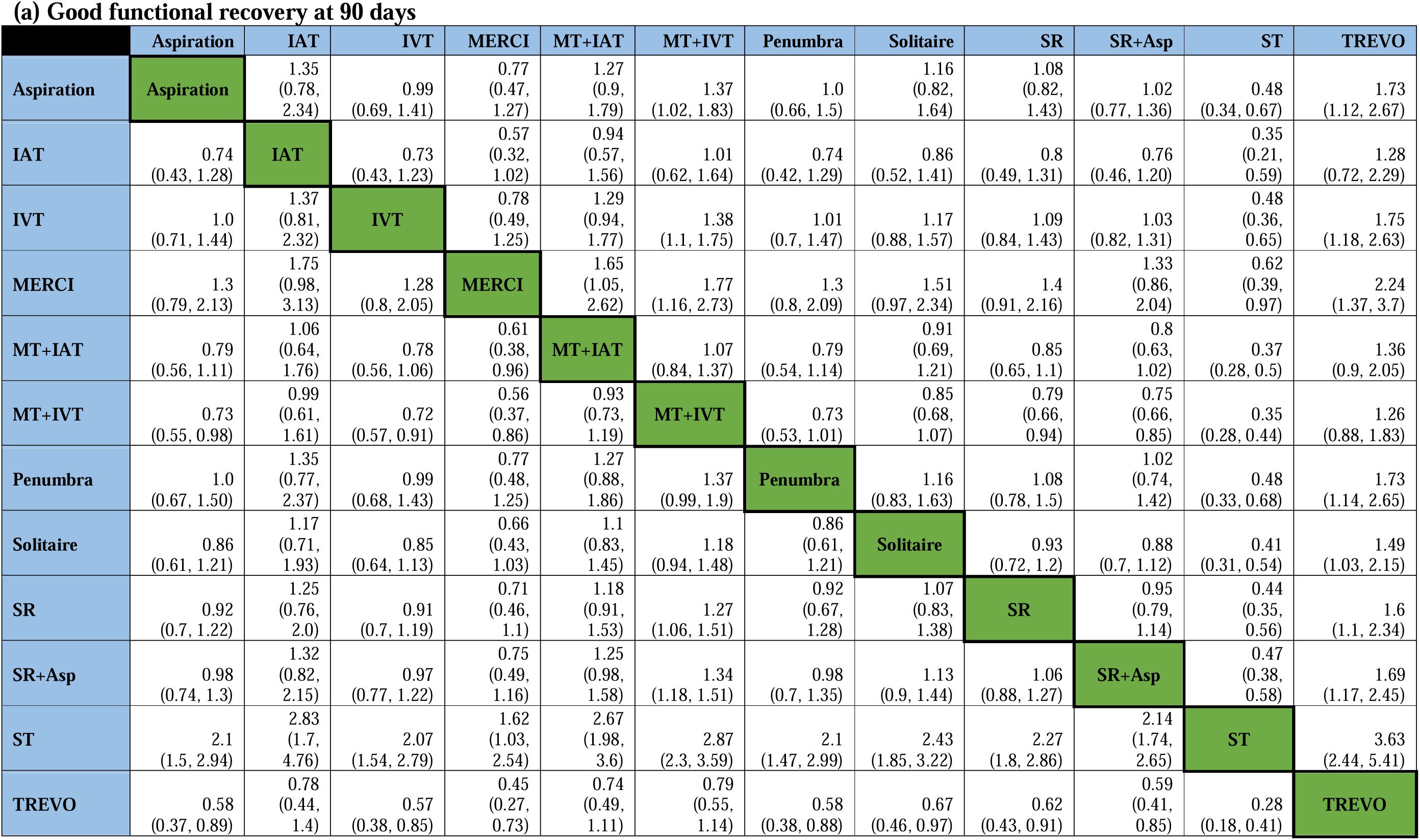
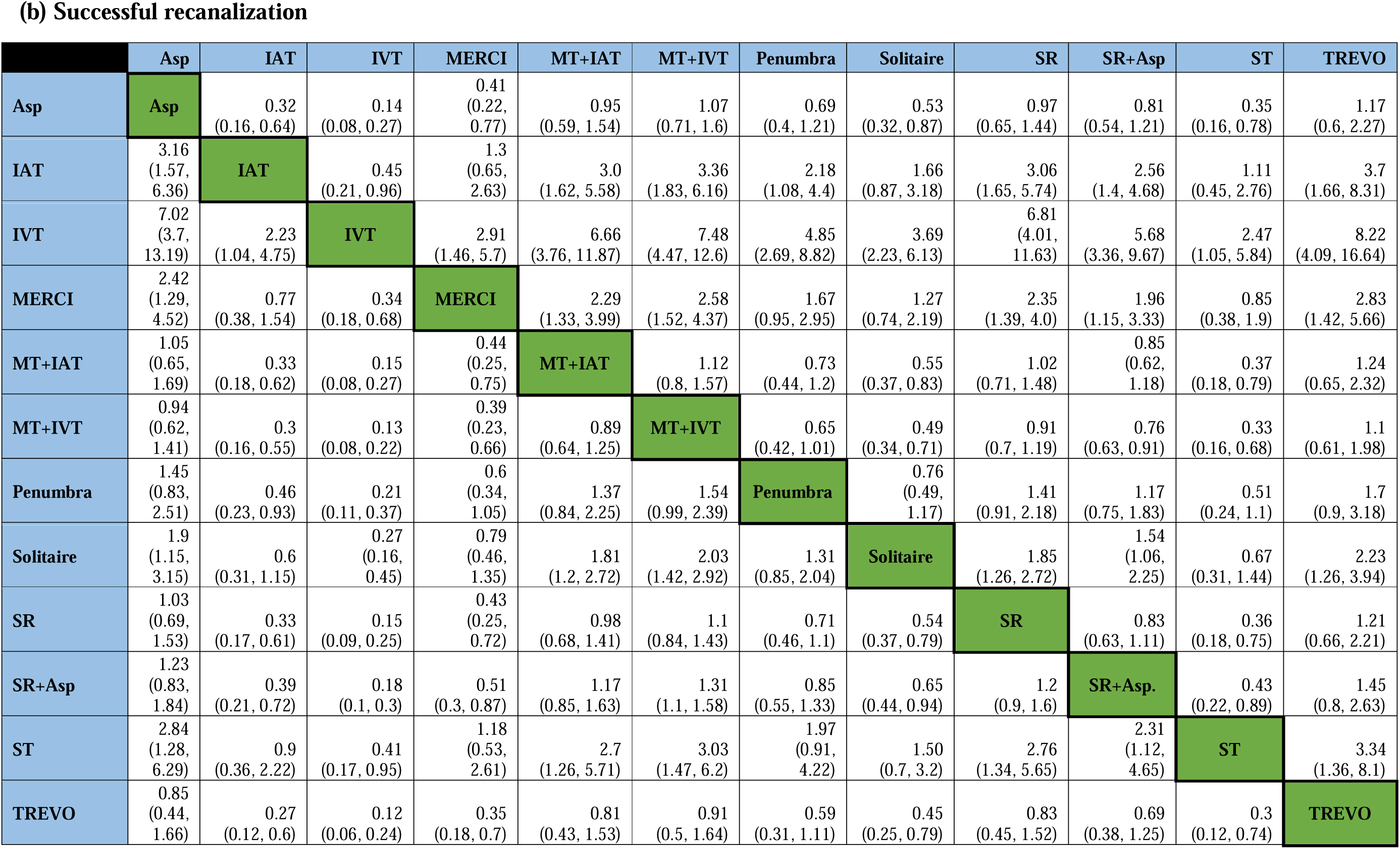
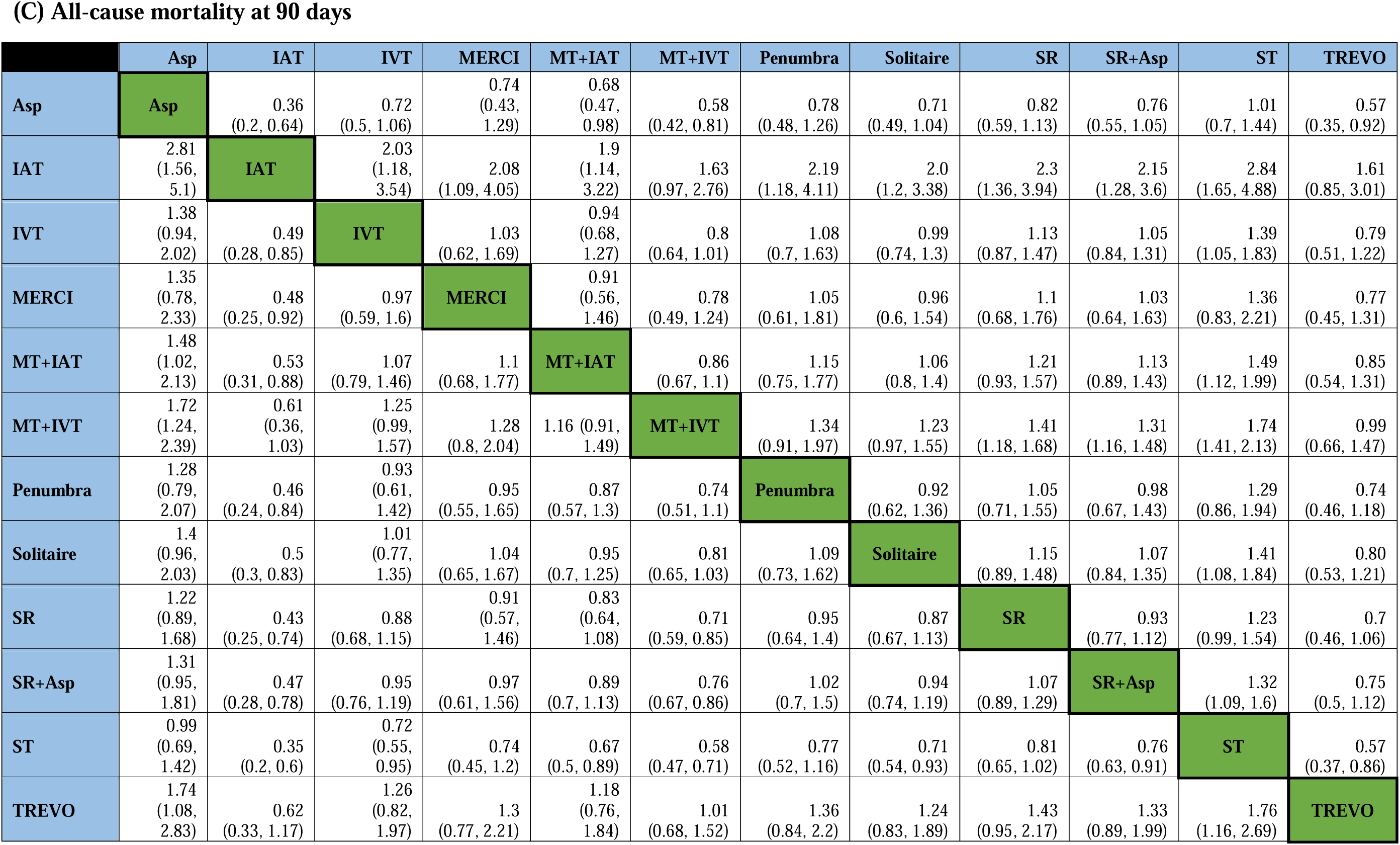
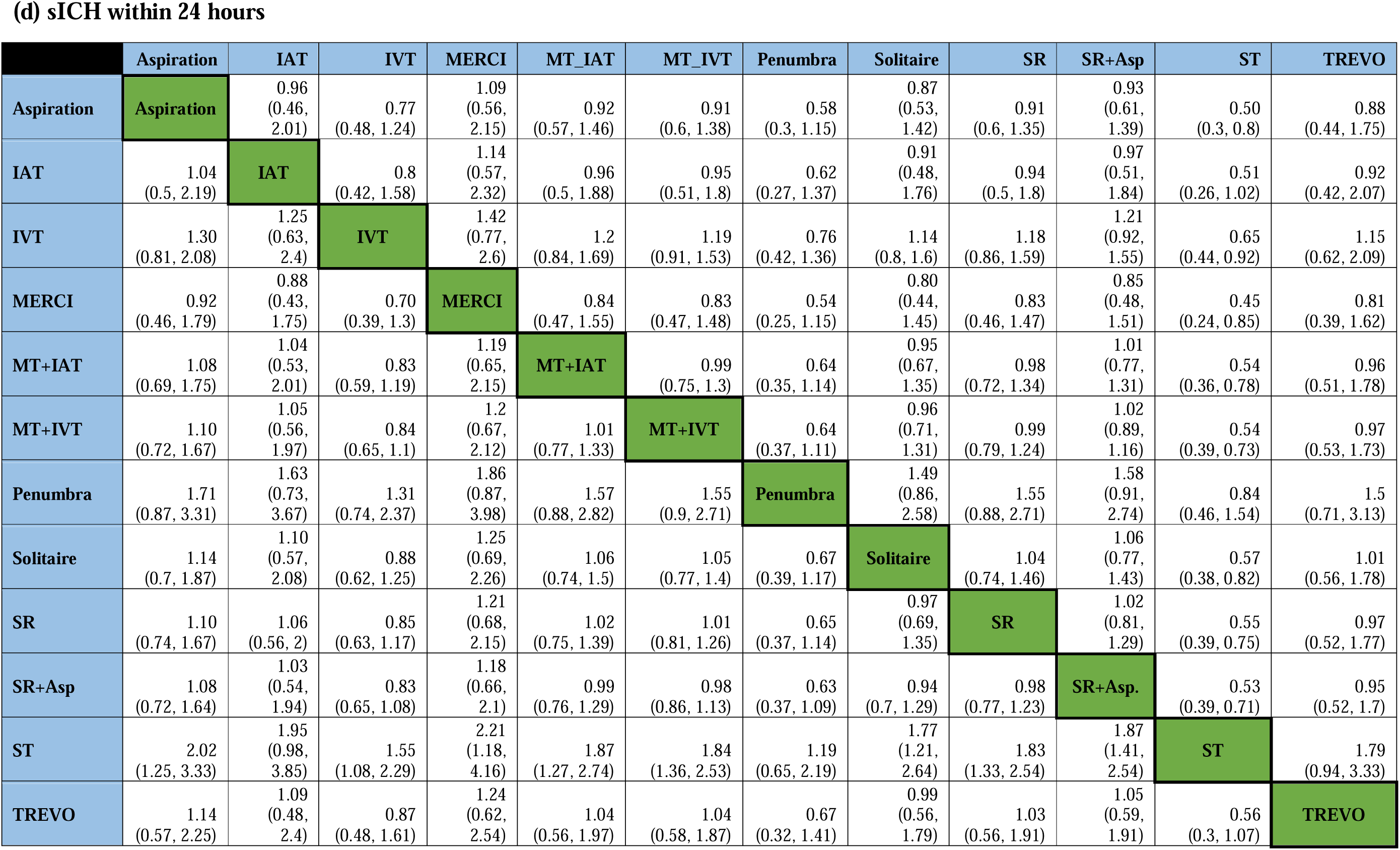
League table representing summary estimates from network meta-analysis for: (a) good functional recovery at 90 days; (b) successful recanalization; (c) all-cause mortality at 90 days and (d) sICH within 24 hours.

**(a) Good functional recovery at 90 days**
|  | Aspiration | IAT | IVT | MERCI | MT+IAT | MT+IVT | Penumbra | Solitaire | SR | SR+Asp | ST | TREVO |
| --- | --- | --- | --- | --- | --- | --- | --- | --- | --- | --- | --- | --- |
| **Aspiration** | **Aspiration** | 1.35<br>(0.78, 2.34) | 0.99<br>(0.69, 1.41) | 0.77<br>(0.47, 1.27) | 1.27<br>(0.9, 1.79) | 1.37<br>(1.02, 1.83) | 1.0<br>(0.66, 1.5) | 1.16<br>(0.82, 1.64) | 1.08<br>(0.82, 1.43) | 1.02<br>(0.77, 1.36) | 0.48<br>(0.34, 0.67) | 1.73<br>(1.12, 2.67) |
| **IAT** | 0.74<br>(0.43, 1.28) | **IAT** | 0.73<br>(0.43, 1.23) | 0.57<br>(0.32, 1.02) | 0.94<br>(0.57, 1.56) | 1.01<br>(0.62, 1.64) | 0.74<br>(0.42, 1.29) | 0.86<br>(0.52, 1.41) | 0.8<br>(0.49, 1.31) | 0.76<br>(0.46, 1.20) | 0.35<br>(0.21, 0.59) | 1.28<br>(0.72, 2.29) |
| **IVT** | 1.0<br>(0.71, 1.44) | 1.37<br>(0.81, 2.32) | **IVT** | 0.78<br>(0.49, 1.25) | 1.29<br>(0.94, 1.77) | 1.38<br>(1.1, 1.75) | 1.01<br>(0.7, 1.47) | 1.17<br>(0.88, 1.57) | 1.09<br>(0.84, 1.43) | 1.03<br>(0.82, 1.31) | 0.48<br>(0.36, 0.65) | 1.75<br>(1.18, 2.63) |
| **MERCI** | 1.3<br>(0.79, 2.13) | 1.75<br>(0.98, 3.13) | 1.28<br>(0.8, 2.05) | **MERCI** | 1.65<br>(1.05, 2.62) | 1.77<br>(1.16, 2.73) | 1.3<br>(0.8, 2.09) | 1.51<br>(0.97, 2.34) | 1.4<br>(0.91, 2.16) | 1.33<br>(0.86, 2.04) | 0.62<br>(0.39, 0.97) | 2.24<br>(1.37, 3.7) |
| **MT+IAT** | 0.79<br>(0.56, 1.11) | 1.06<br>(0.64, 1.76) | 0.78<br>(0.56, 1.06) | 0.61<br>(0.38, 0.96) | **MT+IAT** | 1.07<br>(0.84, 1.37) | 0.79<br>(0.54, 1.14) | 0.91<br>(0.69, 1.21) | 0.85<br>(0.65, 1.1) | 0.8<br>(0.63, 1.02) | 0.37<br>(0.28, 0.5) | 1.36<br>(0.9, 2.05) |
| **MT+IVT** | 0.73<br>(0.55, 0.98) | 0.99<br>(0.61, 1.61) | 0.72<br>(0.57, 0.91) | 0.56<br>(0.37, 0.86) | 0.93<br>(0.73, 1.19) | **MT+IVT** | 0.73<br>(0.53, 1.01) | 0.85<br>(0.68, 1.07) | 0.79<br>(0.66, 0.94) | 0.75<br>(0.66, 0.85) | 0.35<br>(0.28, 0.44) | 1.26<br>(0.88, 1.83) |
| **Penumbra** | 1.0<br>(0.67, 1.50) | 1.35<br>(0.77, 2.37) | 0.99<br>(0.68, 1.43) | 0.77<br>(0.48, 1.25) | 1.27<br>(0.88, 1.86) | 1.37<br>(0.99, 1.9) | **Penumbra** | 1.16<br>(0.83, 1.63) | 1.08<br>(0.78, 1.5) | 1.02<br>(0.74, 1.42) | 0.48<br>(0.33, 0.68) | 1.73<br>(1.14, 2.65) |
| **Solitaire** | 0.86<br>(0.61, 1.21) | 1.17<br>(0.71, 1.93) | 0.85<br>(0.64, 1.13) | 0.66<br>(0.43, 1.03) | 1.1<br>(0.83, 1.45) | 1.18<br>(0.94, 1.48) | 0.86<br>(0.61, 1.21) | **Solitaire** | 0.93<br>(0.72, 1.2) | 0.88<br>(0.7, 1.12) | 0.41<br>(0.31, 0.54) | 1.49<br>(1.03, 2.15) |
| **SR** | 0.92<br>(0.7, 1.22) | 1.25<br>(0.76, 2.0) | 0.91<br>(0.7, 1.19) | 0.71<br>(0.46, 1.1) | 1.18<br>(0.91, 1.53) | 1.27<br>(1.06, 1.51) | 0.92<br>(0.67, 1.28) | 1.07<br>(0.83, 1.38) | **SR** | 0.95<br>(0.79, 1.14) | 0.44<br>(0.35, 0.56) | 1.6<br>(1.1, 2.34) |
| **SR+Asp** | 0.98<br>(0.74, 1.3) | 1.32<br>(0.82, 2.15) | 0.97<br>(0.77, 1.22) | 0.75<br>(0.49, 1.16) | 1.25<br>(0.98, 1.58) | 1.34<br>(1.18, 1.51) | 0.98<br>(0.7, 1.35) | 1.13<br>(0.9, 1.44) | 1.06<br>(0.88, 1.27) | **SR+Asp** | 0.47<br>(0.38, 0.58) | 1.69<br>(1.17, 2.45) |
| **ST** | 2.1<br>(1.5, 2.94) | 2.83<br>(1.7, 4.76) | 2.07<br>(1.54, 2.79) | 1.62<br>(1.03, 2.54) | 2.67<br>(1.98, 3.6) | 2.87<br>(2.3, 3.59) | 2.1<br>(1.47, 2.99) | 2.43<br>(1.85, 3.22) | 2.27<br>(1.8, 2.86) | 2.14<br>(1.74, 2.65) | **ST** | 3.63<br>(2.44, 5.41) |
| **TREVO** | 0.58<br>(0.37, 0.89) | 0.78<br>(0.44, 1.4) | 0.57<br>(0.38, 0.85) | 0.45<br>(0.27, 0.73) | 0.74<br>(0.49, 1.11) | 0.79<br>(0.55, 1.14) | 0.58<br>(0.38, 0.88) | 0.67<br>(0.46, 0.97) | 0.62<br>(0.43, 0.91) | 0.59<br>(0.41, 0.85) | 0.28<br>(0.18, 0.41) | **TREVO** |

**(b) Successful recanalization**
|  | Asp | IAT | IVT | MERCI | MT+IAT | MT+IVT | Penumbra | Solitaire | SR | SR+Asp | ST | TREVO |
| --- | --- | --- | --- | --- | --- | --- | --- | --- | --- | --- | --- | --- |
| Asp | Asp | 0.32<br>(0.16, 0.64) | 0.14<br>(0.08, 0.27) | 0.41<br>(0.22, 0.77) | 0.95<br>(0.59, 1.54) | 1.07<br>(0.71, 1.6) | 0.69<br>(0.4, 1.21) | 0.53<br>(0.32, 0.87) | 0.97<br>(0.65, 1.44) | 0.81<br>(0.54, 1.21) | 0.35<br>(0.16, 0.78) | 1.17<br>(0.6, 2.27) |
| IAT | 3.16<br>(1.57, 6.36) | IAT | 0.45<br>(0.21, 0.96) | 1.3<br>(0.65, 2.63) | 3.0<br>(1.62, 5.58) | 3.36<br>(1.83, 6.16) | 2.18<br>(1.08, 4.4) | 1.66<br>(0.87, 3.18) | 3.06<br>(1.65, 5.74) | 2.56<br>(1.4, 4.68) | 1.11<br>(0.45, 2.76) | 3.7<br>(1.66, 8.31) |
| IVT | 7.02<br>(3.7, 13.19) | 2.23<br>(1.04, 4.75) | IVT | 2.91<br>(1.46, 5.7) | 6.66<br>(3.76, 11.87) | 7.48<br>(4.47, 12.6) | 4.85<br>(2.69, 8.82) | 3.69<br>(2.23, 6.13) | 6.81<br>(4.01, 11.63) | 5.68<br>(3.36, 9.67) | 2.47<br>(1.05, 5.84) | 8.22<br>(4.09, 16.64) |
| MERCI | 2.42<br>(1.29, 4.52) | 0.77<br>(0.38, 1.54) | 0.34<br>(0.18, 0.68) | MERCI | 2.29<br>(1.33, 3.99) | 2.58<br>(1.52, 4.37) | 1.67<br>(0.95, 2.95) | 1.27<br>(0.74, 2.19) | 2.35<br>(1.39, 4.0) | 1.96<br>(1.15, 3.33) | 0.85<br>(0.38, 1.9) | 2.83<br>(1.42, 5.66) |
| MT+IAT | 1.05<br>(0.65, 1.69) | 0.33<br>(0.18, 0.62) | 0.15<br>(0.08, 0.27) | 0.44<br>(0.25, 0.75) | MT+IAT | 1.12<br>(0.8, 1.57) | 0.73<br>(0.44, 1.2) | 0.55<br>(0.37, 0.83) | 1.02<br>(0.71, 1.48) | 0.85<br>(0.62, 1.18) | 0.37<br>(0.18, 0.79) | 1.24<br>(0.65, 2.32) |
| MT+IVT | 0.94<br>(0.62, 1.41) | 0.3<br>(0.16, 0.55) | 0.13<br>(0.08, 0.22) | 0.39<br>(0.23, 0.66) | 0.89<br>(0.64, 1.25) | MT+IVT | 0.65<br>(0.42, 1.01) | 0.49<br>(0.34, 0.71) | 0.91<br>(0.7, 1.19) | 0.76<br>(0.63, 0.91) | 0.33<br>(0.16, 0.68) | 1.1<br>(0.61, 1.98) |
| Penumbra | 1.45<br>(0.83, 2.51) | 0.46<br>(0.23, 0.93) | 0.21<br>(0.11, 0.37) | 0.6<br>(0.34, 1.05) | 1.37<br>(0.84, 2.25) | 1.54<br>(0.99, 2.39) | Penumbra | 0.76<br>(0.49, 1.17) | 1.41<br>(0.91, 2.18) | 1.17<br>(0.75, 1.83) | 0.51<br>(0.24, 1.1) | 1.7<br>(0.9, 3.18) |
| Solitaire | 1.9<br>(1.15, 3.15) | 0.6<br>(0.31, 1.15) | 0.27<br>(0.16, 0.45) | 0.79<br>(0.46, 1.35) | 1.81<br>(1.2, 2.72) | 2.03<br>(1.42, 2.92) | 1.31<br>(0.85, 2.04) | Solitaire | 1.85<br>(1.26, 2.72) | 1.54<br>(1.06, 2.25) | 0.67<br>(0.31, 1.44) | 2.23<br>(1.26, 3.94) |
| SR | 1.03<br>(0.69, 1.53) | 0.33<br>(0.17, 0.61) | 0.15<br>(0.09, 0.25) | 0.43<br>(0.25, 0.72) | 0.98<br>(0.68, 1.41) | 1.1<br>(0.84, 1.43) | 0.71<br>(0.46, 1.1) | 0.54<br>(0.37, 0.79) | SR | 0.83<br>(0.63, 1.11) | 0.36<br>(0.18, 0.75) | 1.21<br>(0.66, 2.21) |
| SR+Asp | 1.23<br>(0.83, 1.84) | 0.39<br>(0.21, 0.72) | 0.18<br>(0.1, 0.3) | 0.51<br>(0.3, 0.87) | 1.17<br>(0.85, 1.63) | 1.31<br>(1.1, 1.58) | 0.85<br>(0.55, 1.33) | 0.65<br>(0.44, 0.94) | 1.2<br>(0.9, 1.6) | SR+Asp. | 0.43<br>(0.22, 0.89) | 1.45<br>(0.8, 2.63) |
| ST | 2.84<br>(1.28, 6.29) | 0.9<br>(0.36, 2.22) | 0.41<br>(0.17, 0.95) | 1.18<br>(0.53, 2.61) | 2.7<br>(1.26, 5.71) | 3.03<br>(1.47, 6.2) | 1.97<br>(0.91, 4.22) | 1.50<br>(0.7, 3.2) | 2.76<br>(1.34, 5.65) | 2.31<br>(1.12, 4.65) | ST | 3.34<br>(1.36, 8.1) |
| TREVO | 0.85<br>(0.44, 1.66) | 0.27<br>(0.12, 0.6) | 0.12<br>(0.06, 0.24) | 0.35<br>(0.18, 0.7) | 0.81<br>(0.43, 1.53) | 0.91<br>(0.5, 1.64) | 0.59<br>(0.31, 1.11) | 0.45<br>(0.25, 0.79) | 0.83<br>(0.45, 1.52) | 0.69<br>(0.38, 1.25) | 0.3<br>(0.12, 0.74) | TREVO |

(C) All-cause mortality at 90 days
|  | Asp | IAT | IVT | MERCI | MT+IAT | MT+IVT | Penumbra | Solitaire | SR | SR+Asp | ST | TREVO |
| --- | --- | --- | --- | --- | --- | --- | --- | --- | --- | --- | --- | --- |
| Asp | Asp | 0.36<br>(0.2, 0.64) | 0.72<br>(0.5, 1.06) | 0.74<br>(0.43, 1.29) | 0.68<br>(0.47, 0.98) | 0.58<br>(0.42, 0.81) | 0.78<br>(0.48, 1.26) | 0.71<br>(0.49, 1.04) | 0.82<br>(0.59, 1.13) | 0.76<br>(0.55, 1.05) | 1.01<br>(0.7, 1.44) | 0.57<br>(0.35, 0.92) |
| IAT | 2.81<br>(1.56, 5.1) | IAT | 2.03<br>(1.18, 3.54) | 2.08<br>(1.09, 4.05) | 1.9<br>(1.14, 3.22) | 1.63<br>(0.97, 2.76) | 2.19<br>(1.18, 4.11) | 2.0<br>(1.2, 3.38) | 2.3<br>(1.36, 3.94) | 2.15<br>(1.28, 3.6) | 2.84<br>(1.65, 4.88) | 1.61<br>(0.85, 3.01) |
| IVT | 1.38<br>(0.94, 2.02) | 0.49<br>(0.28, 0.85) | IVT | 1.03<br>(0.62, 1.69) | 0.94<br>(0.68, 1.27) | 0.8<br>(0.64, 1.01) | 1.08<br>(0.7, 1.63) | 0.99<br>(0.74, 1.3) | 1.13<br>(0.87, 1.47) | 1.05<br>(0.84, 1.31) | 1.39<br>(1.05, 1.83) | 0.79<br>(0.51, 1.22) |
| MERCI | 1.35<br>(0.78, 2.33) | 0.48<br>(0.25, 0.92) | 0.97<br>(0.59, 1.6) | MERCI | 0.91<br>(0.56, 1.46) | 0.78<br>(0.49, 1.24) | 1.05<br>(0.61, 1.81) | 0.96<br>(0.6, 1.54) | 1.1<br>(0.68, 1.76) | 1.03<br>(0.64, 1.63) | 1.36<br>(0.83, 2.21) | 0.77<br>(0.45, 1.31) |
| MT+IAT | 1.48<br>(1.02, 2.13) | 0.53<br>(0.31, 0.88) | 1.07<br>(0.79, 1.46) | 1.1<br>(0.68, 1.77) | MT+IAT | 0.86<br>(0.67, 1.1) | 1.15<br>(0.75, 1.77) | 1.06<br>(0.8, 1.4) | 1.21<br>(0.93, 1.57) | 1.13<br>(0.89, 1.43) | 1.49<br>(1.12, 1.99) | 0.85<br>(0.54, 1.31) |
| MT+IVT | 1.72<br>(1.24, 2.39) | 0.61<br>(0.36, 1.03) | 1.25<br>(0.99, 1.57) | 1.28<br>(0.8, 2.04) | 1.16 (0.91, 1.49) | MT+IVT | 1.34<br>(0.91, 1.97) | 1.23<br>(0.97, 1.55) | 1.41<br>(1.18, 1.68) | 1.31<br>(1.16, 1.48) | 1.74<br>(1.41, 2.13) | 0.99<br>(0.66, 1.47) |
| Penumbra | 1.28<br>(0.79, 2.07) | 0.46<br>(0.24, 0.84) | 0.93<br>(0.61, 1.42) | 0.95<br>(0.55, 1.65) | 0.87<br>(0.57, 1.3) | 0.74<br>(0.51, 1.1) | Penumbra | 0.92<br>(0.62, 1.36) | 1.05<br>(0.71, 1.55) | 0.98<br>(0.67, 1.43) | 1.29<br>(0.86, 1.94) | 0.74<br>(0.46, 1.18) |
| Solitaire | 1.4<br>(0.96, 2.03) | 0.5<br>(0.3, 0.83) | 1.01<br>(0.77, 1.35) | 1.04<br>(0.65, 1.67) | 0.95<br>(0.7, 1.25) | 0.81<br>(0.65, 1.03) | 1.09<br>(0.73, 1.62) | Solitaire | 1.15<br>(0.89, 1.48) | 1.07<br>(0.84, 1.35) | 1.41<br>(1.08, 1.84) | 0.80<br>(0.53, 1.21) |
| SR | 1.22<br>(0.89, 1.68) | 0.43<br>(0.25, 0.74) | 0.88<br>(0.68, 1.15) | 0.91<br>(0.57, 1.46) | 0.83<br>(0.64, 1.08) | 0.71<br>(0.59, 0.85) | 0.95<br>(0.64, 1.4) | 0.87<br>(0.67, 1.13) | SR | 0.93<br>(0.77, 1.12) | 1.23<br>(0.99, 1.54) | 0.7<br>(0.46, 1.06) |
| SR+Asp | 1.31<br>(0.95, 1.81) | 0.47<br>(0.28, 0.78) | 0.95<br>(0.76, 1.19) | 0.97<br>(0.61, 1.56) | 0.89<br>(0.7, 1.13) | 0.76<br>(0.67, 0.86) | 1.02<br>(0.7, 1.5) | 0.94<br>(0.74, 1.19) | 1.07<br>(0.89, 1.29) | SR+Asp | 1.32<br>(1.09, 1.6) | 0.75<br>(0.5, 1.12) |
| ST | 0.99<br>(0.69, 1.42) | 0.35<br>(0.2, 0.6) | 0.72<br>(0.55, 0.95) | 0.74<br>(0.45, 1.2) | 0.67<br>(0.5, 0.89) | 0.58<br>(0.47, 0.71) | 0.77<br>(0.52, 1.16) | 0.71<br>(0.54, 0.93) | 0.81<br>(0.65, 1.02) | 0.76<br>(0.63, 0.91) | ST | 0.57<br>(0.37, 0.86) |
| TREVO | 1.74<br>(1.08, 2.83) | 0.62<br>(0.33, 1.17) | 1.26<br>(0.82, 1.97) | 1.3<br>(0.77, 2.21) | 1.18<br>(0.76, 1.84) | 1.01<br>(0.68, 1.52) | 1.36<br>(0.84, 2.2) | 1.24<br>(0.83, 1.89) | 1.43<br>(0.95, 2.17) | 1.33<br>(0.89, 1.99) | 1.76<br>(1.16, 2.69) | TREVO |

(d) sICH within 24 hours
|  | Aspiration | IAT | IVT | MERCI | MT_IAT | MT_IVT | Penumbra | Solitaire | SR | SR+Asp | ST | TREVO |
| --- | --- | --- | --- | --- | --- | --- | --- | --- | --- | --- | --- | --- |
| Aspiration | Aspiration | 0.96<br>(0.46, 2.01) | 0.77<br>(0.48, 1.24) | 1.09<br>(0.56, 2.15) | 0.92<br>(0.57, 1.46) | 0.91<br>(0.6, 1.38) | 0.58<br>(0.3, 1.15) | 0.87<br>(0.53, 1.42) | 0.91<br>(0.6, 1.35) | 0.93<br>(0.61, 1.39) | 0.50<br>(0.3, 0.8) | 0.88<br>(0.44, 1.75) |
| IAT | 1.04<br>(0.5, 2.19) | IAT | 0.8<br>(0.42, 1.58) | 1.14<br>(0.57, 2.32) | 0.96<br>(0.5, 1.88) | 0.95<br>(0.51, 1.8) | 0.62<br>(0.27, 1.37) | 0.91<br>(0.48, 1.76) | 0.94<br>(0.5, 1.8) | 0.97<br>(0.51, 1.84) | 0.51<br>(0.26, 1.02) | 0.92<br>(0.42, 2.07) |
| IVT | 1.30<br>(0.81, 2.08) | 1.25<br>(0.63, 2.4) | IVT | 1.42<br>(0.77, 2.6) | 1.2<br>(0.84, 1.69) | 1.19<br>(0.91, 1.53) | 0.76<br>(0.42, 1.36) | 1.14<br>(0.8, 1.6) | 1.18<br>(0.86, 1.59) | 1.21<br>(0.92, 1.55) | 0.65<br>(0.44, 0.92) | 1.15<br>(0.62, 2.09) |
| MERCI | 0.92<br>(0.46, 1.79) | 0.88<br>(0.43, 1.75) | 0.70<br>(0.39, 1.3) | MERCI | 0.84<br>(0.47, 1.55) | 0.83<br>(0.47, 1.48) | 0.54<br>(0.25, 1.15) | 0.80<br>(0.44, 1.45) | 0.83<br>(0.46, 1.47) | 0.85<br>(0.48, 1.51) | 0.45<br>(0.24, 0.85) | 0.81<br>(0.39, 1.62) |
| MT+IAT | 1.08<br>(0.69, 1.75) | 1.04<br>(0.53, 2.01) | 0.83<br>(0.59, 1.19) | 1.19<br>(0.65, 2.15) | MT+IAT | 0.99<br>(0.75, 1.3) | 0.64<br>(0.35, 1.14) | 0.95<br>(0.67, 1.35) | 0.98<br>(0.72, 1.34) | 1.01<br>(0.77, 1.31) | 0.54<br>(0.36, 0.78) | 0.96<br>(0.51, 1.78) |
| MT+IVT | 1.10<br>(0.72, 1.67) | 1.05<br>(0.56, 1.97) | 0.84<br>(0.65, 1.1) | 1.2<br>(0.67, 2.12) | 1.01<br>(0.77, 1.33) | MT+IVT | 0.64<br>(0.37, 1.11) | 0.96<br>(0.71, 1.31) | 0.99<br>(0.79, 1.24) | 1.02<br>(0.89, 1.16) | 0.54<br>(0.39, 0.73) | 0.97<br>(0.53, 1.73) |
| Penumbra | 1.71<br>(0.87, 3.31) | 1.63<br>(0.73, 3.67) | 1.31<br>(0.74, 2.37) | 1.86<br>(0.87, 3.98) | 1.57<br>(0.88, 2.82) | 1.55<br>(0.9, 2.71) | Penumbra | 1.49<br>(0.86, 2.58) | 1.55<br>(0.88, 2.71) | 1.58<br>(0.91, 2.74) | 0.84<br>(0.46, 1.54) | 1.5<br>(0.71, 3.13) |
| Solitaire | 1.14<br>(0.7, 1.87) | 1.10<br>(0.57, 2.08) | 0.88<br>(0.62, 1.25) | 1.25<br>(0.69, 2.26) | 1.06<br>(0.74, 1.5) | 1.05<br>(0.77, 1.4) | 0.67<br>(0.39, 1.17) | Solitaire | 1.04<br>(0.74, 1.46) | 1.06<br>(0.77, 1.43) | 0.57<br>(0.38, 0.82) | 1.01<br>(0.56, 1.78) |
| SR | 1.10<br>(0.74, 1.67) | 1.06<br>(0.56, 2) | 0.85<br>(0.63, 1.17) | 1.21<br>(0.68, 2.15) | 1.02<br>(0.75, 1.39) | 1.01<br>(0.81, 1.26) | 0.65<br>(0.37, 1.14) | 0.97<br>(0.69, 1.35) | SR | 1.02<br>(0.81, 1.29) | 0.55<br>(0.39, 0.75) | 0.97<br>(0.52, 1.77) |
| SR+Asp | 1.08<br>(0.72, 1.64) | 1.03<br>(0.54, 1.94) | 0.83<br>(0.65, 1.08) | 1.18<br>(0.66, 2.1) | 0.99<br>(0.76, 1.29) | 0.98<br>(0.86, 1.13) | 0.63<br>(0.37, 1.09) | 0.94<br>(0.7, 1.29) | 0.98<br>(0.77, 1.23) | SR+Asp. | 0.53<br>(0.39, 0.71) | 0.95<br>(0.52, 1.7) |
| ST | 2.02<br>(1.25, 3.33) | 1.95<br>(0.98, 3.85) | 1.55<br>(1.08, 2.29) | 2.21<br>(1.18, 4.16) | 1.87<br>(1.27, 2.74) | 1.84<br>(1.36, 2.53) | 1.19<br>(0.65, 2.19) | 1.77<br>(1.21, 2.64) | 1.83<br>(1.33, 2.54) | 1.87<br>(1.41, 2.54) | ST | 1.79<br>(0.94, 3.33) |
| TREVO | 1.14<br>(0.57, 2.25) | 1.09<br>(0.48, 2.4) | 0.87<br>(0.48, 1.61) | 1.24<br>(0.62, 2.54) | 1.04<br>(0.56, 1.97) | 1.04<br>(0.58, 1.87) | 0.67<br>(0.32, 1.41) | 0.99<br>(0.56, 1.79) | 1.03<br>(0.56, 1.91) | 1.05<br>(0.59, 1.91) | 0.56<br>(0.3, 1.07) | TREVO |

**Table eS4.**
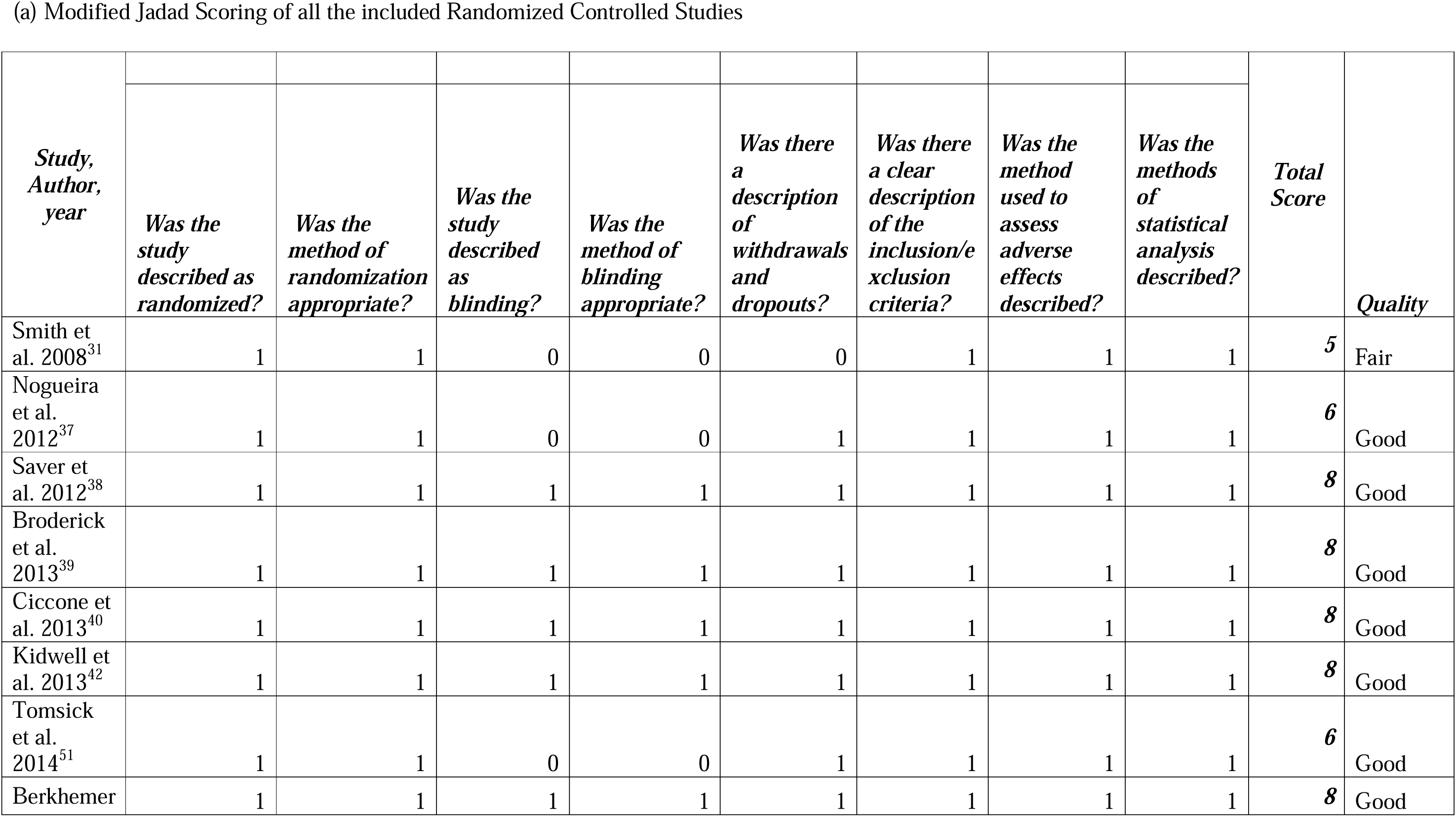
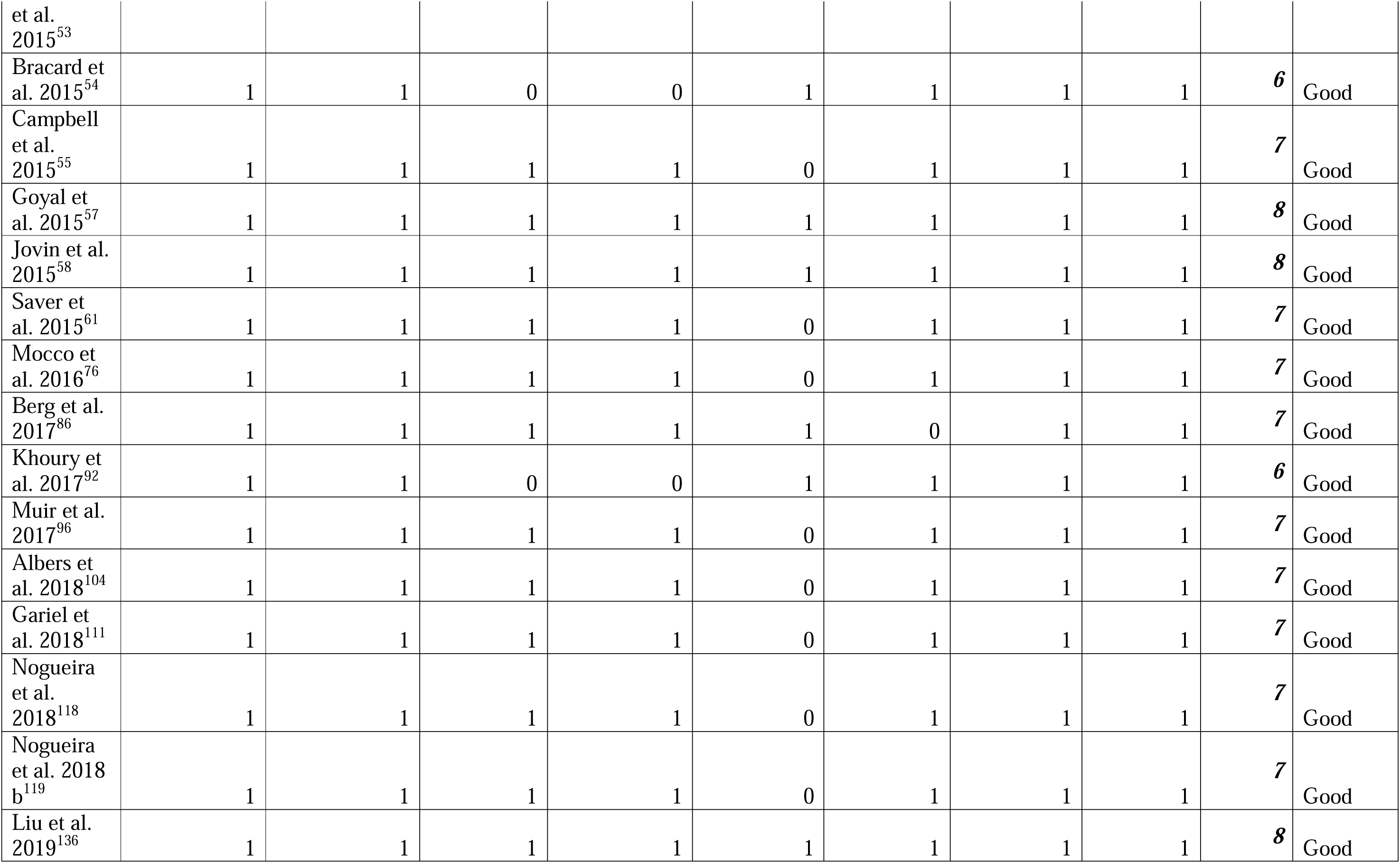
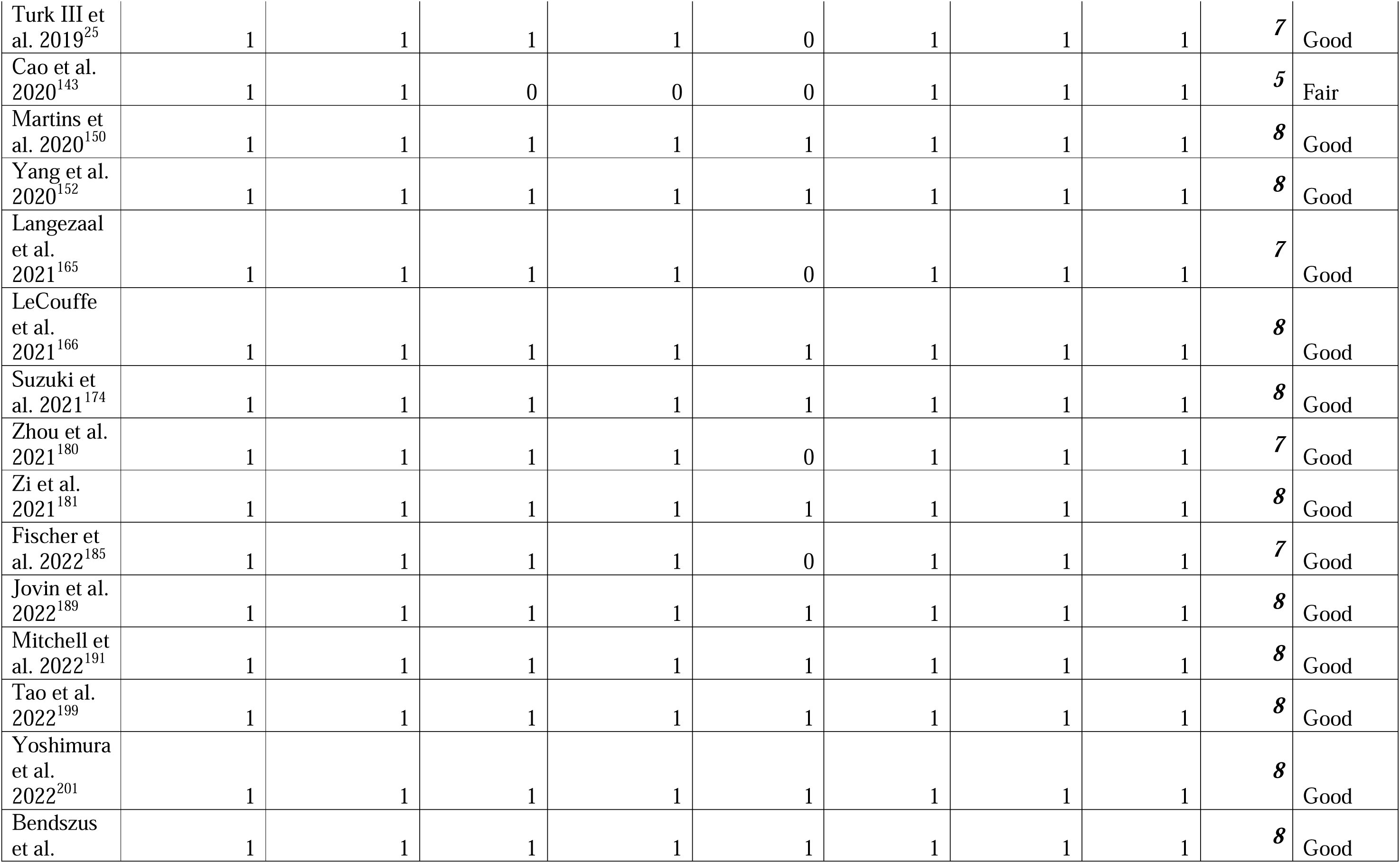
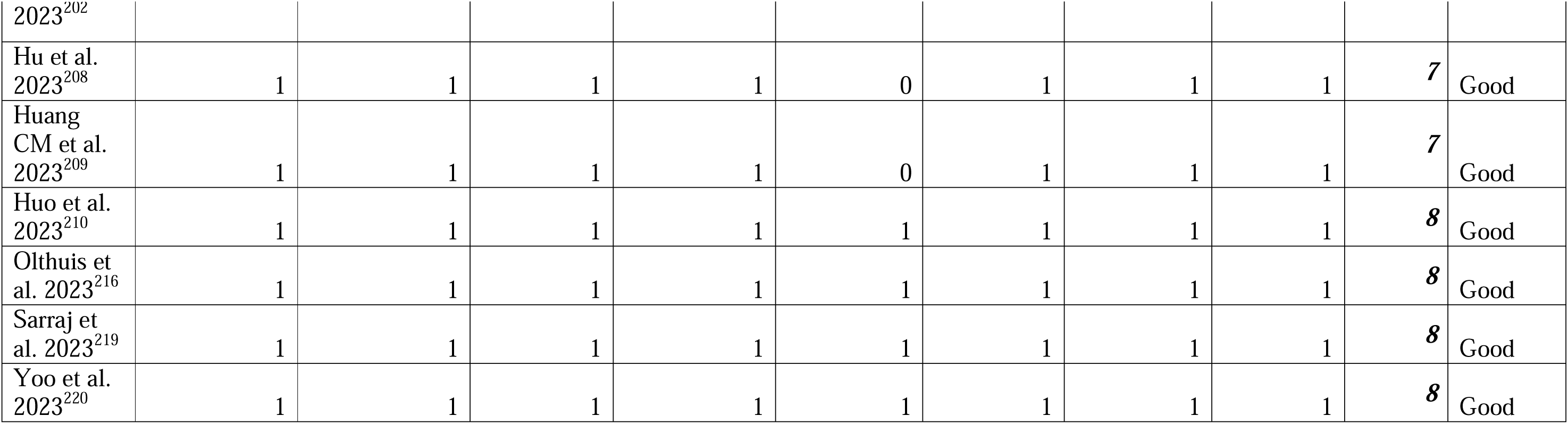
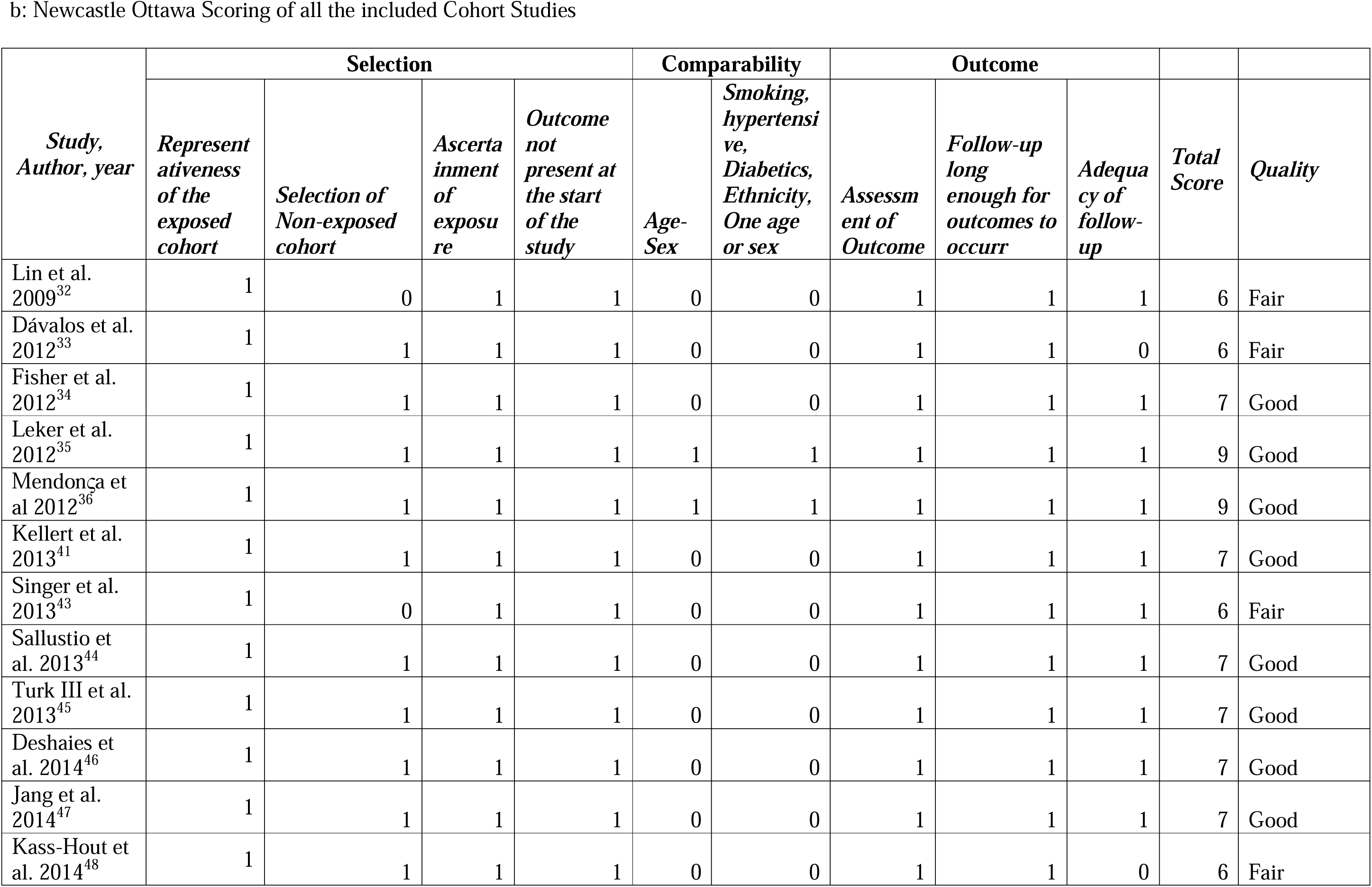
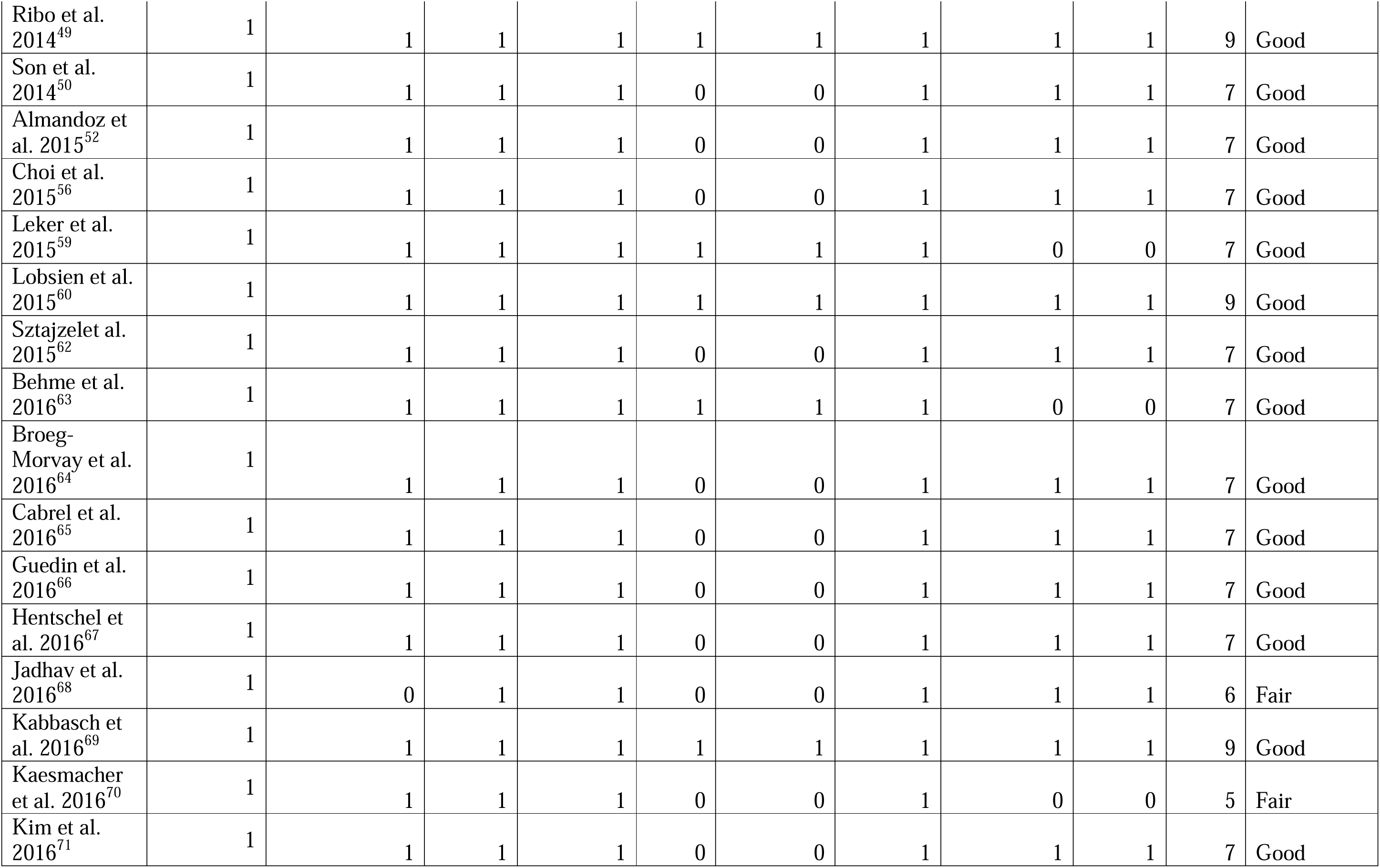
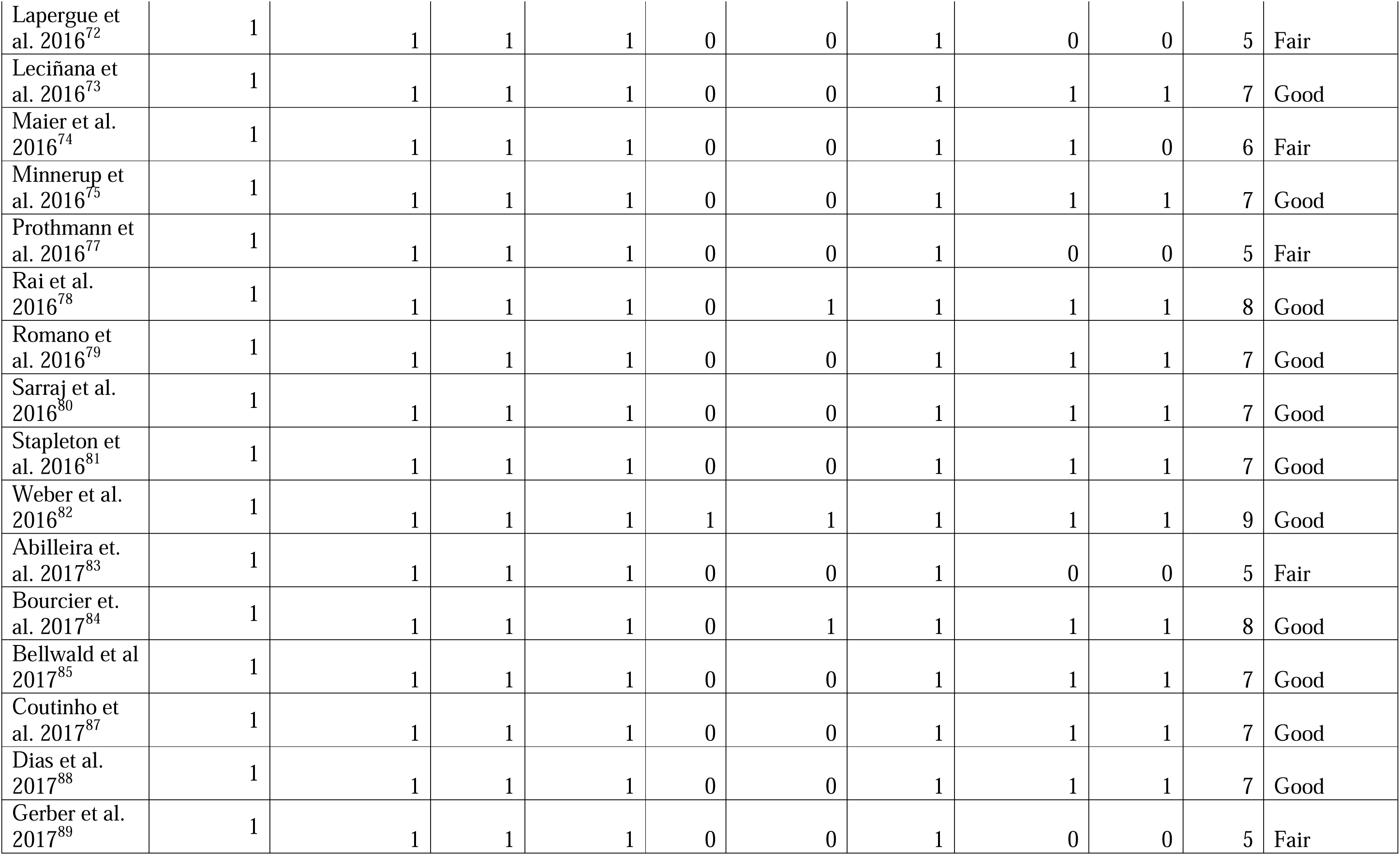
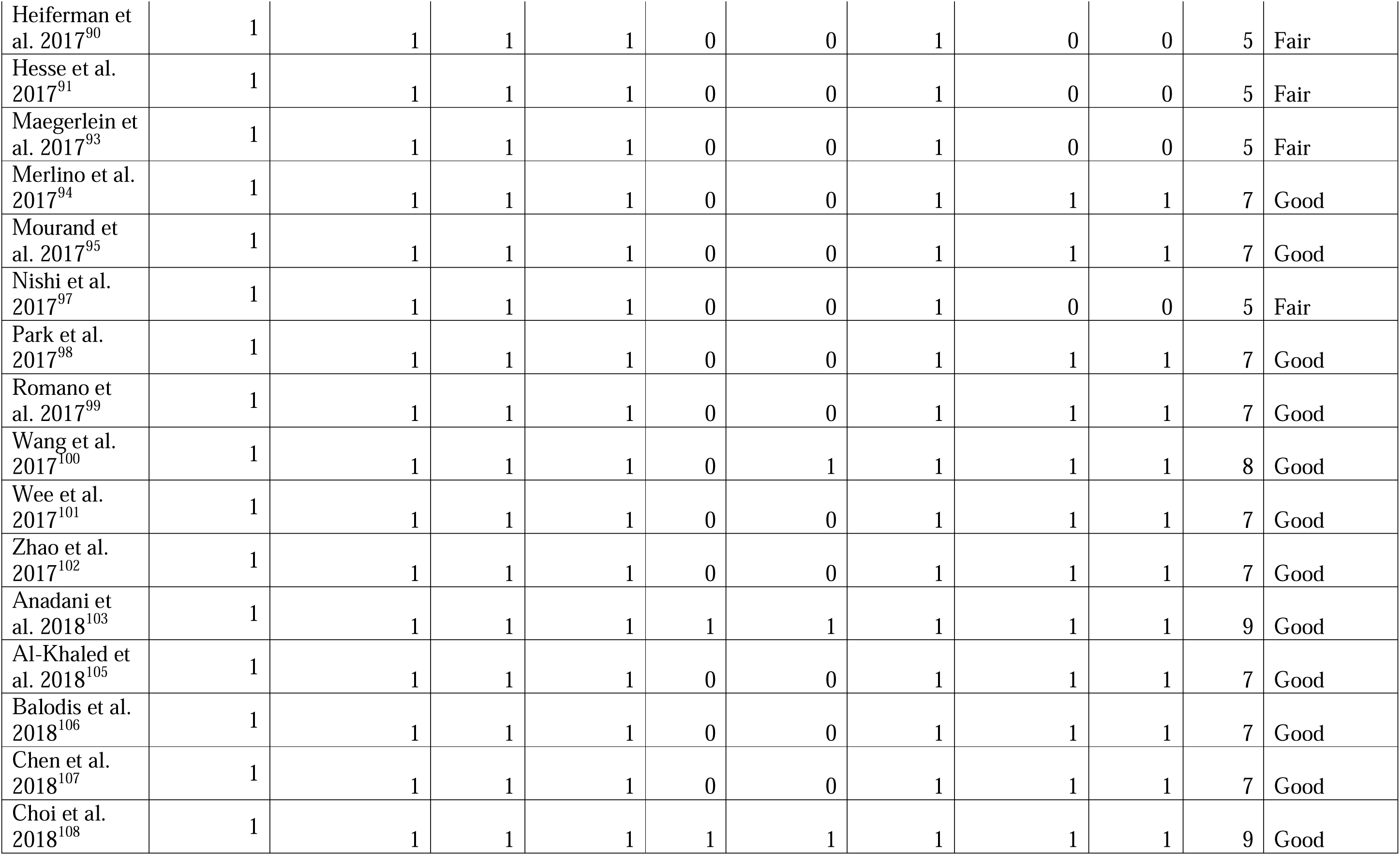
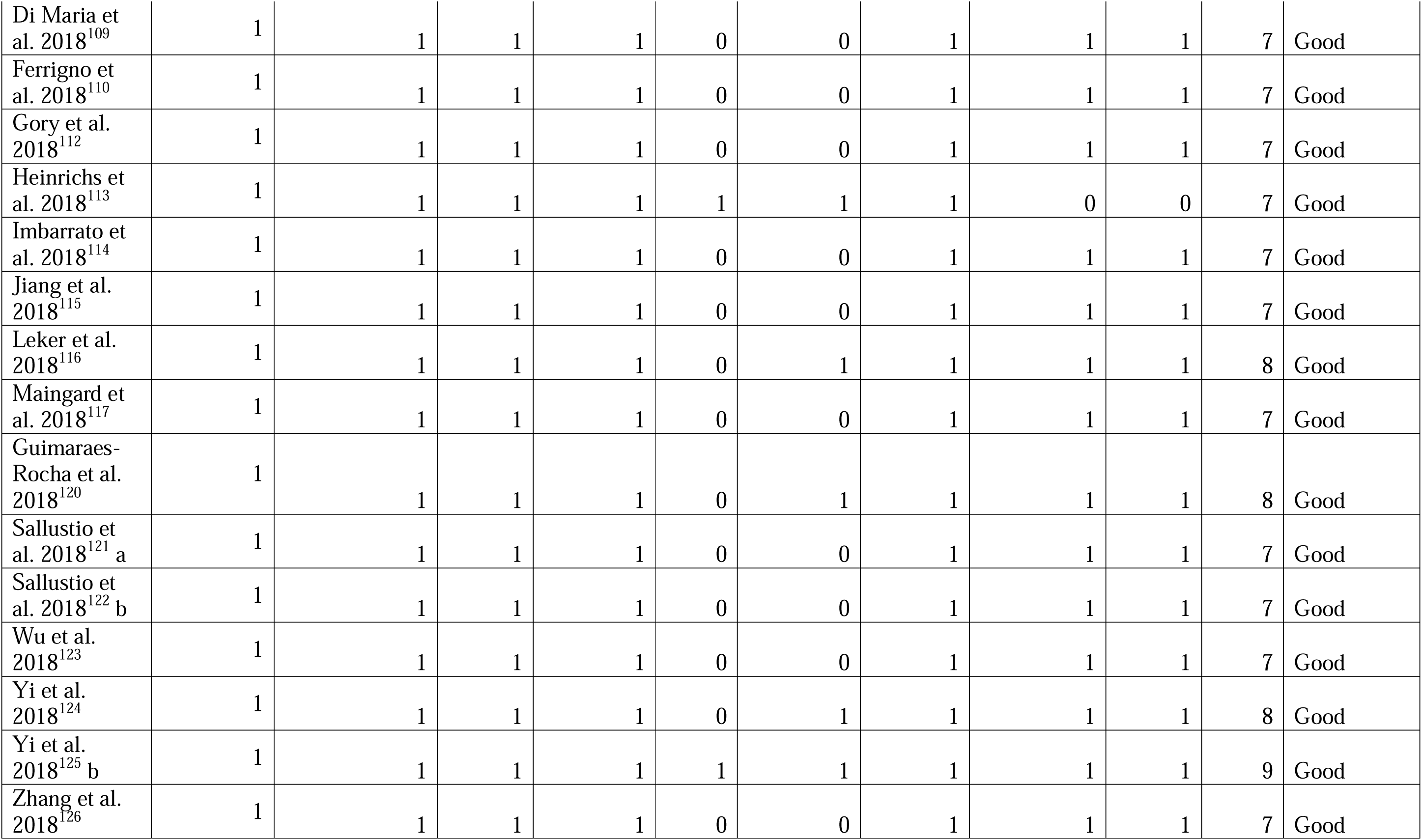
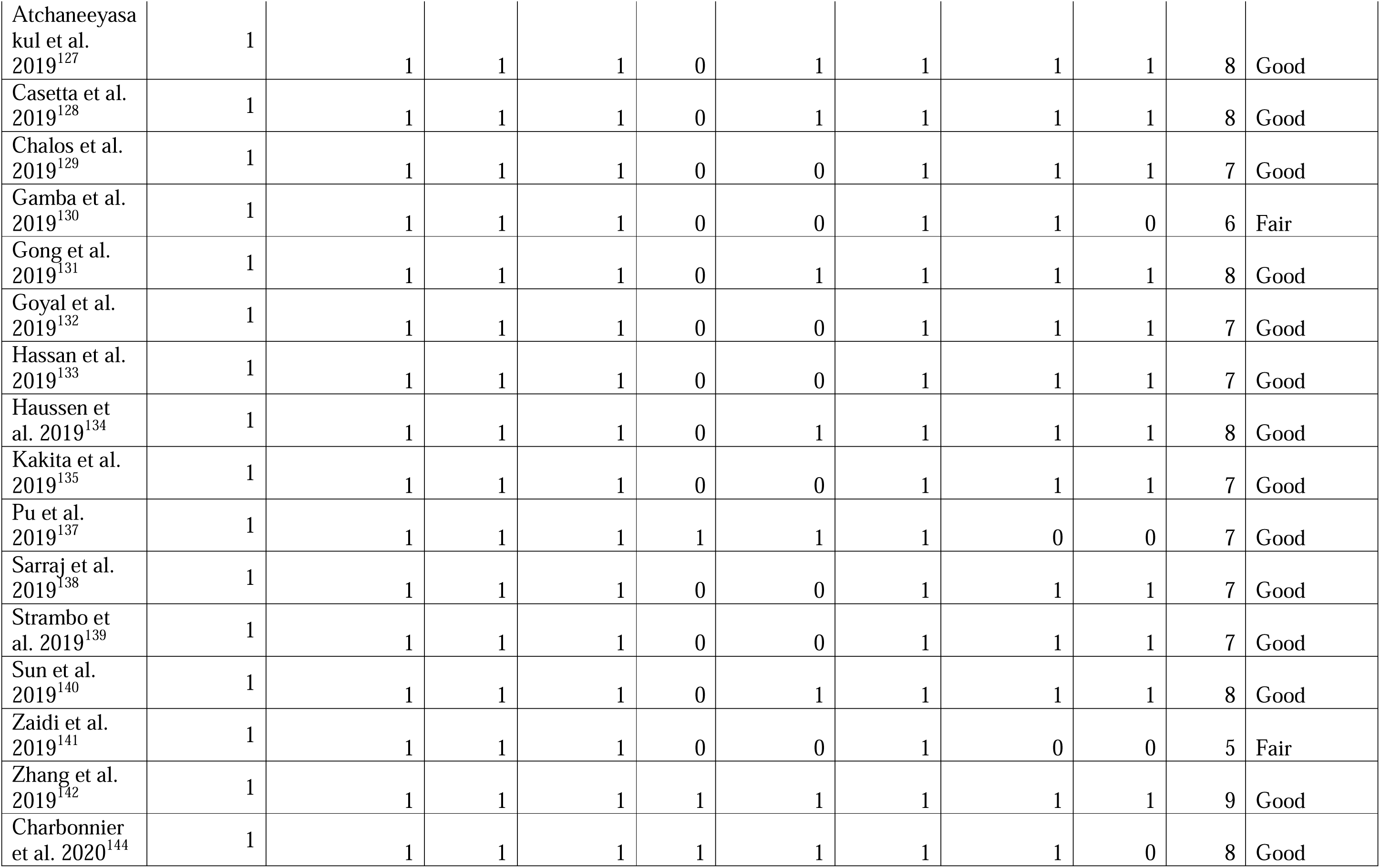
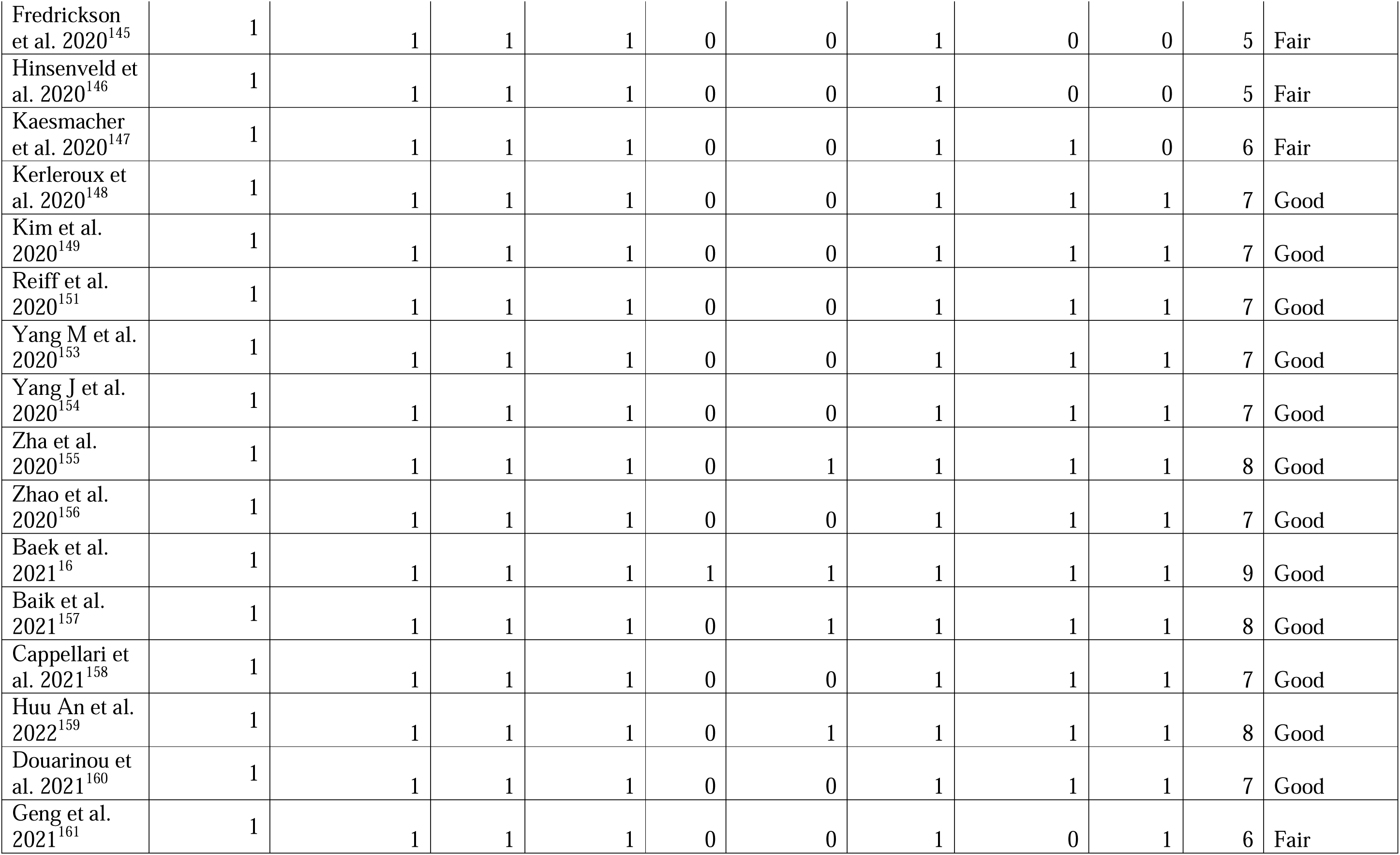
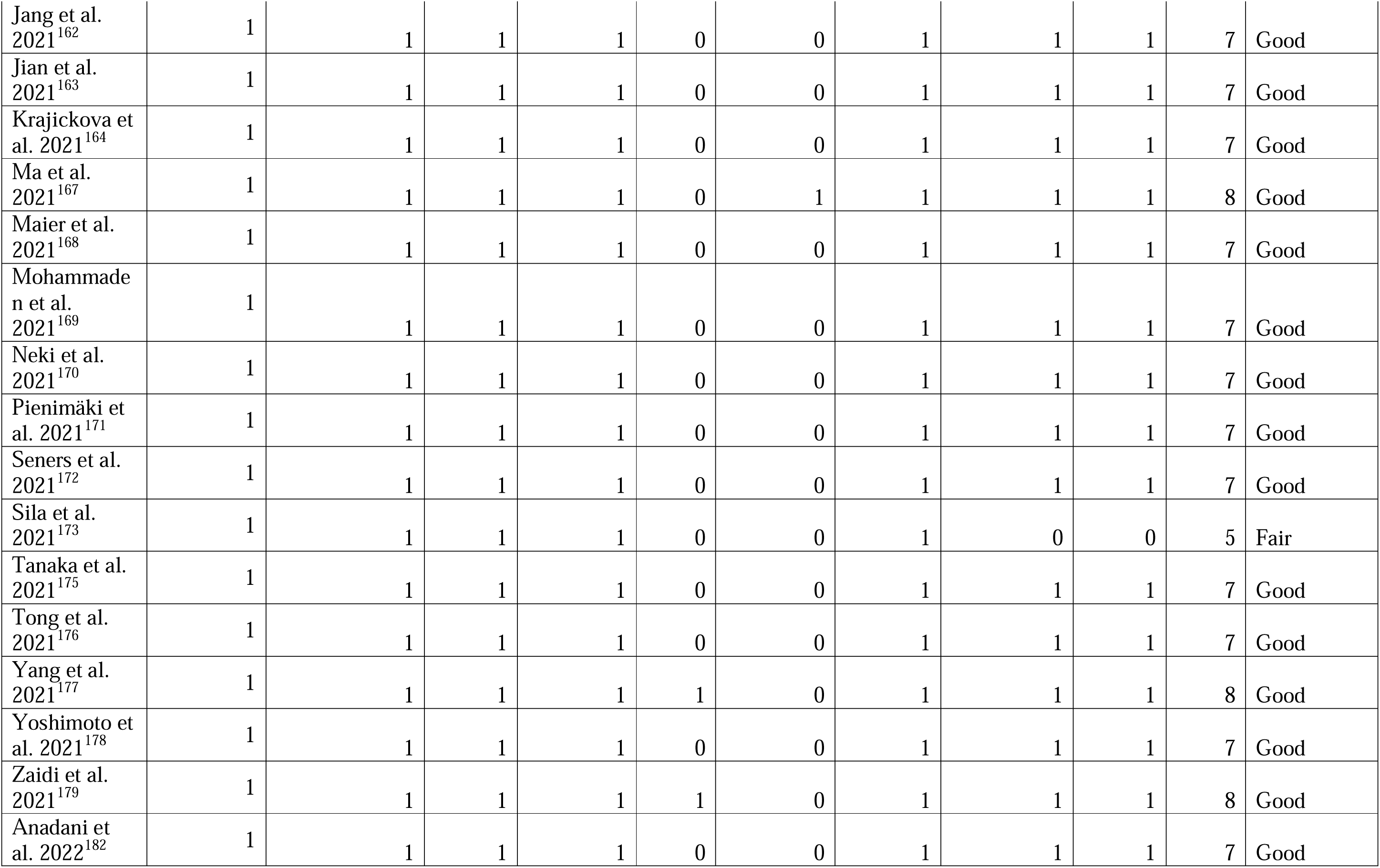
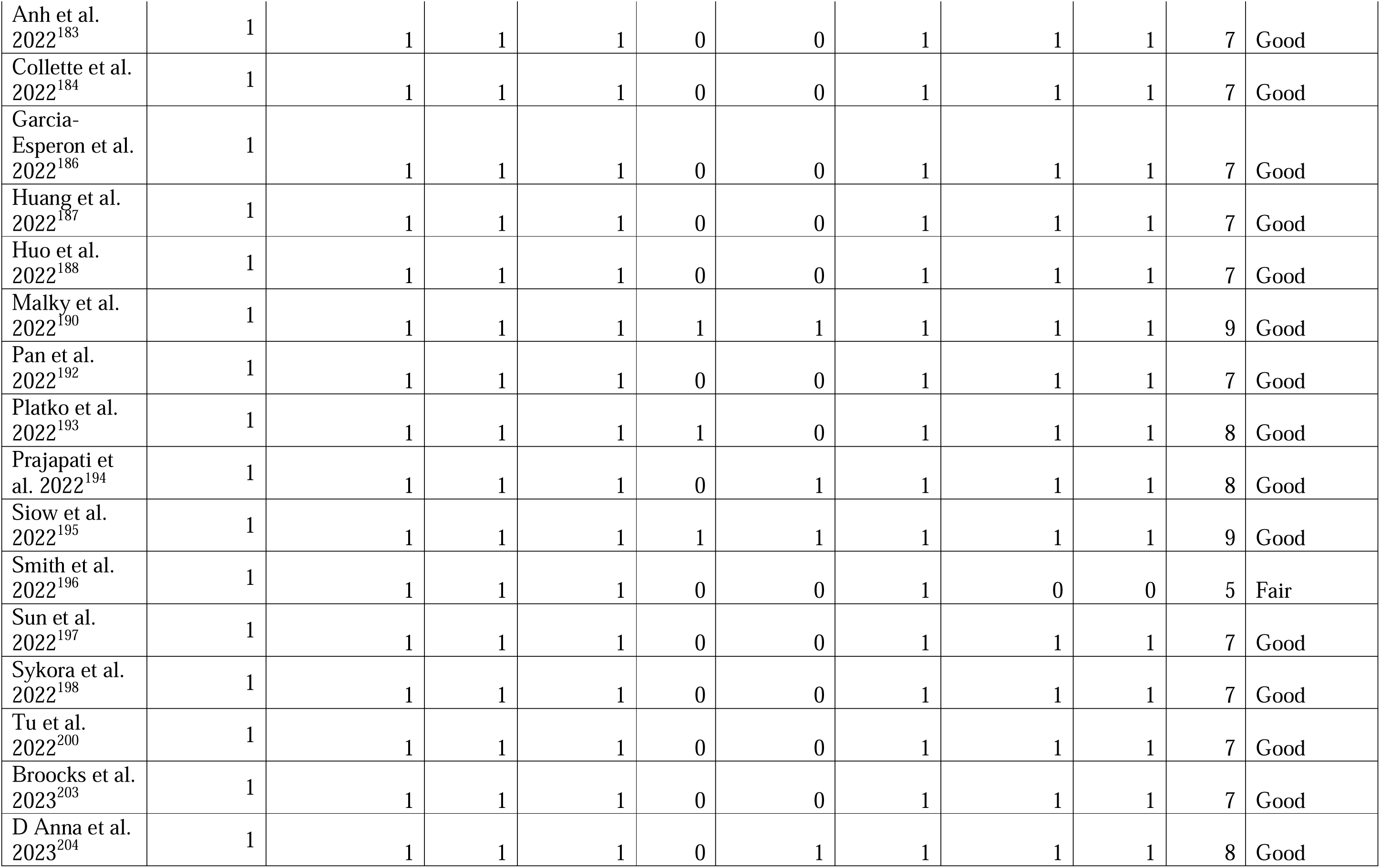
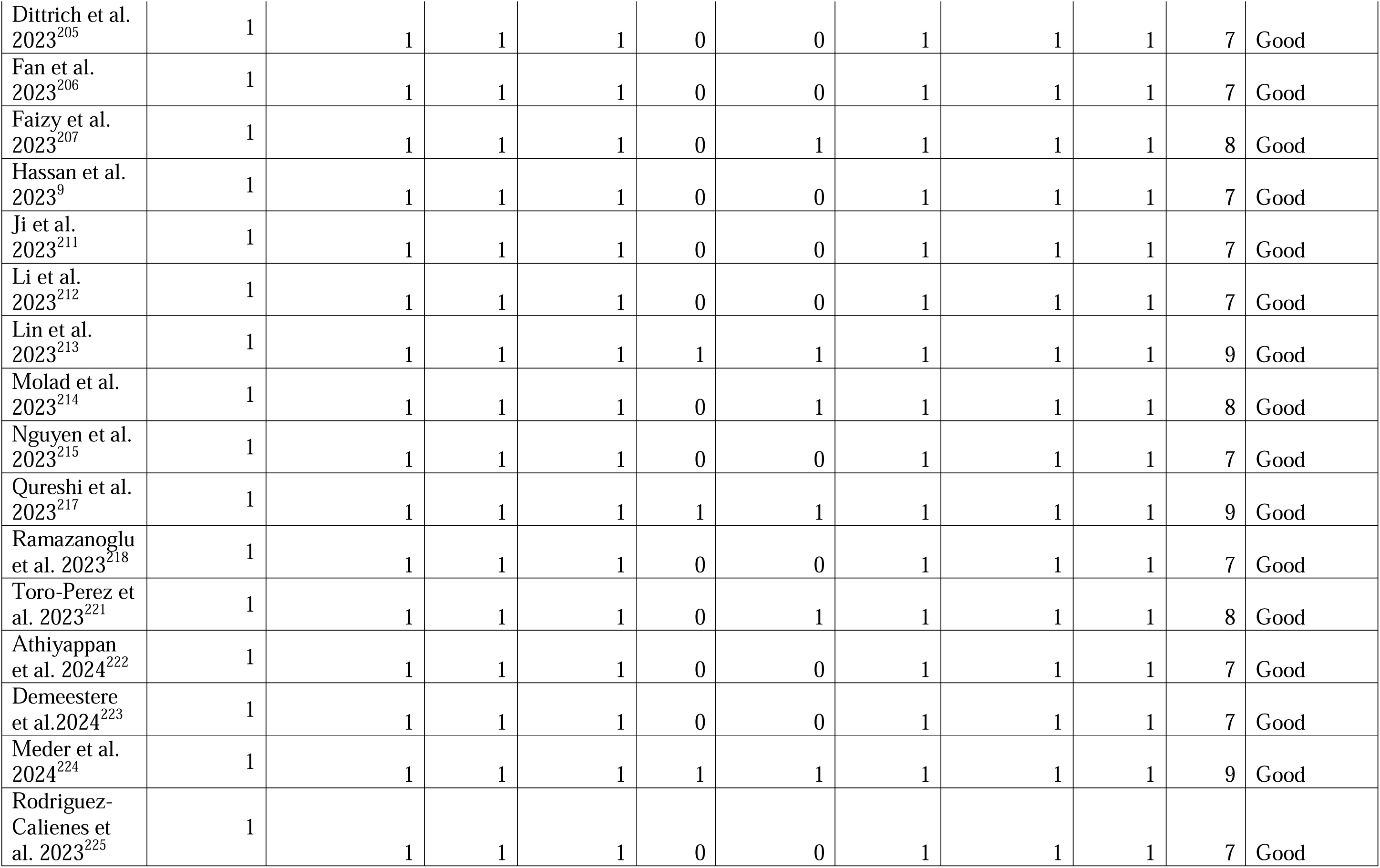
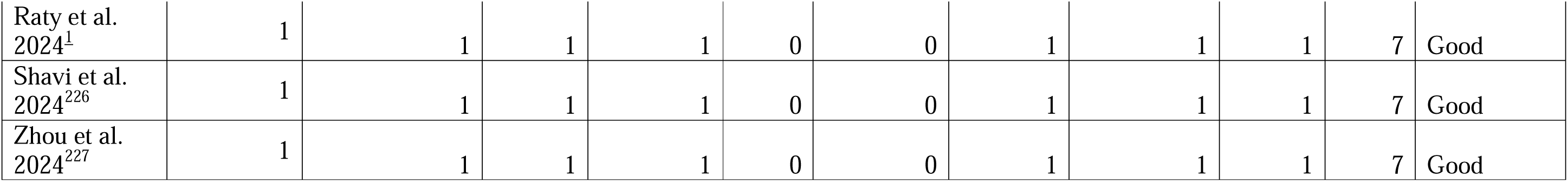
**(a-b):** Quality assessment using (a) Modified Jadad Scoring of all the included Randomized Controlled Studies (b) Newcastle Ottawa Scoring of all the included Cohort Studies **Figure eS1:** Distribution of Thrombectomy Devices across safety and efficacy outcomes.

