## Supplementary File for "Evaluating Thrombectomy Devices and Combination Therapies in Acute ischemic Stroke: A systematic review & Network Meta-Analysis of 201 studies"

**eFigure S1:** Distribution of Thrombectomy Devices for included studies for both safety and efficacy outcomes

Distribution of Thrombectomy devices

|  | mRS | Mortality | sICH | Recanalization |
| --- | --- | --- | --- | --- |
| Aspiration | 17 | 13 | 10 | 19 |
| IAT | 5 | 6 | 3 | 7 |
| IVT | 21 | 22 | 20 | 9 |
| MERCI | 6 | 5 | 4 | 8 |
| MT + IAT | 21 | 21 | 19 | 24 |
| MT + IVT | 80 | 70 | 65 | 77 |
| Penumbra | 12 | 8 | 6 | 16 |
| ST | 27 | 25 | 22 | 4 |
| Solitaire | 31 | 32 | 27 | 30 |
| SR | 44 | 36 | 34 | 40 |
| SR + Asp | 85 | 76 | 70 | 74 |
| TREVO | 10 | 9 | 7 | 8 |
